# Diffusion kurtosis imaging (gen)omics unravels mechanisms of cerebral small vessel disease

**DOI:** 10.64898/2026.08.12.26360241

**Authors:** Quentin Le Grand, Alexandra Koch, Mohammed-Aslam Imtiaz, Greta Maier, Valentina Talevi, Dan Liu, N. Ahmad Aziz, Monique M.B. Breteler

**Affiliations:** Population Health Sciences, German Center for Neurodegenerative Diseases (DZNE), Bonn, Germany; University of Bonn, University Hospital Bonn, Centre of Neurology, Bonn, Germany; University of Bonn, University Hospital Bonn, Institute for Medical Biometry, Informatics and Epidemiology (IMBIE), Bonn, Germany

## Abstract

Cerebral small vessel disease (cSVD) is a leading cause of stroke and dementia. Traditional MRI-markers of cSVD are mainly detectable in older adults, but diffusion MRI (dMRI) measures of white matter microstructure can capture changes predisposing to cSVD earlier in life. In this study, we conducted large genomics and omics explorations of diffusion kurtosis imaging (DKI) dMRI markers, to better characterize the underlying biological mechanisms and explore their clinical relevance in relation to cognition, established cSVD MRI-markers and dementia. We conducted a genome-wide association study (GWAS) of DKI markers in the population-based Rhineland Study (N=5 930). We identified four genome-wide significant loci associated with DKI markers at chr3p25.1 (*LINC00620*-*WNT7A*), chr5q14.2 (*VCAN*), chr5q14.3 (*VCAN-AS1*) and chr8q24.21 (*CCDC26*), and 11 additional suggestive loci. Lead SNPs at chr5q14.3 and chr17q25.1 were associated with white matter hyperintensity volume, chr3p25.1 with white matter perivascular spaces, and chr7p11.2 with Alzheimer disease. Using a transcriptome-wide association study, we identified 17 genes with genetically determined expression associated with DKI markers, including 14 at the chr17q21.31 suggestive GWAS locus. Finally, we identified eight proteins associated with DKI markers in GWAS suggestive loci. Of these, MAD1L1, EGFR and GFAP were also associated with cognitive decline, and MAD1L1 with white matter hyperintensity volume. In conclusion, leveraging omics data, our study identified novel molecular determinants of DKI markers, providing important novel insights into life course determinants of cSVD, a leading cause of stroke and dementia.

## Introduction

Cerebral small vessel disease (cSVD) is a leading cause of stroke and vascular cognitive impairment. Moreover, it is also likely the main pathological substrate underlying the vascular contribution to dementia [1, 2]. cSVD is most often covert, i.e. detectable on brain imaging in asymptomatic individuals, but covert cSVD has also been associated with an increased risk of stroke and dementia [3, 4]. MRI-markers of cSVD, including white matter hyperintensities (WMH) and perivascular spaces (PVS), are key markers of vascular brain aging and highly prevalent in old age [5–7]. Thus, covert cSVD represents a promising target for the prevention of stroke and dementia in the general population. Despite recent advances, the biological mechanisms underlying cSVD are still largely unknown [4, 8, 9]. Improving our understanding of these mechanisms is crucial to identify novel therapeutic targets.

Genome-wide association studies (GWAS) are a powerful tool to unravel molecular mechanisms underlying complex diseases. Such analyses allowed to identify numerous genetic variants associated with MRI-markers of cSVD [4, 8, 9]. These traditional MRI markers of cSVD are predominantly detectable in late middle to older age. However, cSVD-related lesions are unlikely to appear suddenly in healthy brain tissue. Diffusion MRI (dMRI) markers of white matter microstructure, which can be automatically measured in large population-based cohorts, have been shown to better characterize clinical deficits and disease progression in cSVD, and earlier, than traditional MRI markers [4, 10–14]. Genetic association studies leveraging diffusion tensor imaging (DTI), the most commonly used type of dMRI, provided evidence that these markers are genetically correlated with WMH, and several additional neurological disorders (including stroke and Alzheimer disease) [15–17]. Beyond genetics, other omics data, such as transcriptomics and proteomics, could provide further insight into the mechanisms underlying the variability of these markers. Thus, exploring the molecular determinants of these markers could enable unravelling of the biological mechanisms underlying cSVD across the lifespan, with potential therapeutic implications at an earlier stage of the disease process.

One of the limitations of DTI is that it simplifies modelling of diffusion processes by assuming a Gaussian distribution of diffusivity [18]. Therefore, more advanced dMRI models have been developed to provide more accurate insights into the complex brain microstructure [19]. Among these models, diffusion kurtosis imaging (DKI), derived from multi-b-value dMRI acquisitions, also allows characterization of the non-Gaussian behavior of diffusion in complex biological tissues, thereby providing more sensitive measures of altered tissue diffusion properties [20]. Specifically, DKI allows estimation of separate indices: axial kurtosis (AK; tissue complexity parallel to the direction of water diffusion), radial kurtosis (RK; tissue complexity perpendicular to the direction of water diffusion) and mean kurtosis (MK; measure of the overall microstructural complexity of the brain) [18]. To date, DKI markers have not only been shown to be associated with cognitive deficits in cSVD, but also with Alzheimer and Parkinson disease, demonstrating improved sensitivity and a better characterization of white matter alterations in patients [11, 18, 21, 22].

Thus, exploring the molecular determinants of DKI markers may provide novel insights into the biological mechanisms underlying alterations of white matter microstructure across the lifespan. However, to our knowledge, only one genetic-association study has been conducted on DKI markers, in a Chinese population (n=4 183) and their genetic component is unknown in European cohorts [23]. In this study, we therefore aimed to explore the genomic, transcriptomic and proteomic determinants of DKI markers and assess their clinical relevance by evaluating their relation to cognition, established cSVD MRI-markers and dementia.

## Materials and methods

### Study Population

We used data from the Rhineland Study, an ongoing community-based prospective cohort study that enrolls inhabitants aged 30 years and above living in the city of Bonn, Germany [24]. We used baseline data of 5 930 participants in whom both genetic array and brain MRI data were available (mean age ± standard deviation (SD): 55.6 ± 13.4years; 58% women) (of whom 2 233 also had proteomics data available). Only participants without a known diagnosis of dementia, multiple sclerosis, stroke and intracranial hemorrhage were included. In addition, we performed age-stratified analyses using age strata defined by age-tertiles: ≤50 (N = 1 995), [50-62[(N = 2 008), and >62 (N = 1 927) years. Detailed information on this cohort is presented in the **Supplementary methods.**

All human research presented in this manuscript was approved by the ethics committee of the Medical Faculty of the University of Bonn and was conducted according to the Declaration of Helsinki. All participants provided written informed consent.

### MRI Acquisition and Phenotyping

MRI acquisition in the Rhineland Study was performed on a 3-Tesla MRI scanner (MAGNETOM Prisma, Siemens Healthineers), MRI protocols are described in the **Supplementary methods** and in detail elsewhere [25, 26]. We generated one global measure across the full white matter for each of the DKI (AK, MK, RK) and DTI (fractional anisotropy [FA] and mean diffusivity [MD]) metrics (**Supplementary methods**). To normalize distributions, we applied a rank-based inverse normal transformation to all DKI and DTI variables. Using the Li & Ji method as implemented in the *poolr* R package to account for correlation between the 3 DKI markers, we estimated the effective number of tests to be two [27].

### Cognitive assessment

The cognitive test battery of the Rhineland Study has been described in detail elsewhere [28]. We computed cognitive domain performance scores for the following domains at baseline (N=8 454) and first follow-up (4 years [mean: 5.18], N=3 481): episodic verbal memory, working memory, executive function and processing speed. These scores were created by averaging the z scores of the tests contributing to each domain (**Supplementary methods**). Global cognition score was computed by averaging the z scores for episodic verbal memory, working memory, executive function and processing speed. Working memory and episodic verbal memory scores were further averaged to produce a total memory composite score. To normalize distributions, we applied a rank-based inverse normal transformation to cognitive variables.

### Genotyping, quality control, and imputation

In the Rhineland Study, genome-wide genotyping was performed using Illumina Infinium Omni2.5Exome-8 BeadChip. Genotypes were imputed to the 1000 Genomes p3v5 reference panel. Quality control procedures are described in the **Supplementary methods** and have been reported in detail elsewhere [29].

### Plasma proteomics data

Blood samples were collected from each participant in EDTA tubes and centrifuged immediately to obtain plasma. The resulting supernatant was aliquoted and stored at −80 °C until further processing. Plasma proteomics were assessed using the Olink Explore Proximity Extension Assay platform, which quantified 2 923 unique proteins. Investigators conducting the experiments were blinded to all sample characteristics and clinical data. After rigorous quality control (described in the **Supplementary methods**), protein abundances were expressed as Normalized Protein eXpression (NPX) values and used for downstream analyses.

### Statistical analyses

Analytical steps are summarized in **Figure 1**.

**Figure 1:**
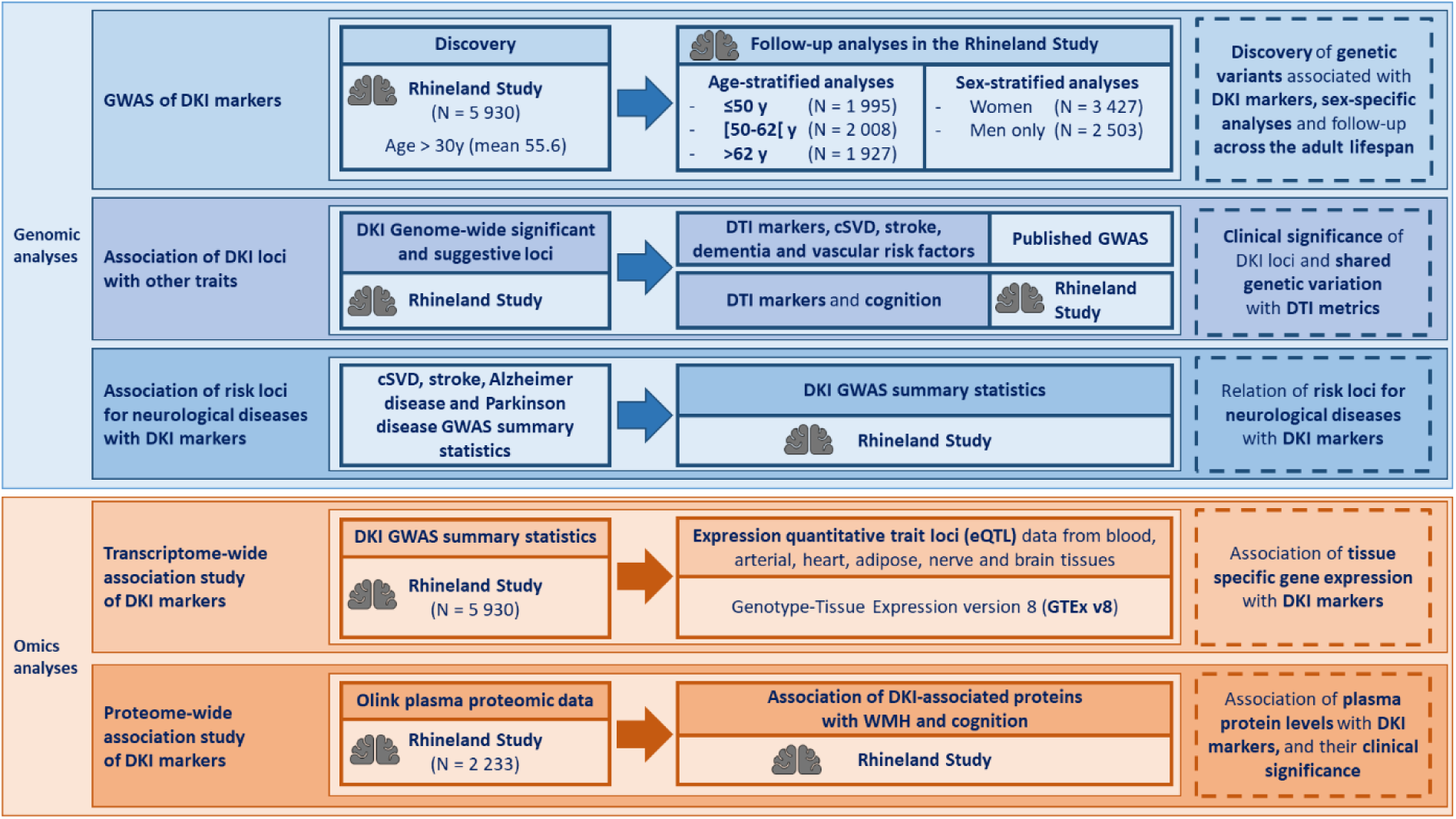
Study workflow. cSVD: Cerebral small vessel disease; DKI: Diffusion kurtosis imaging; DTI: Diffusion tensor imaging; GWAS: Genome-wide association study.

#### Genomic analyses

##### GWAS

We performed GWAS of DKI markers using genome-wide linear mixed models implemented in REGENIE (v3.2.8) [30]. Analyses were restricted to Single Nucleotide Polymorphisms (SNPs) with an imputation score >0.5 and a minor allele frequency (MAF) >0.01 and adjusted for age at MRI, sex, total intracranial volume and the first ten principal components of population stratification (details in **Supplementary methods**). In addition to SNPs reaching genome-wide significance at p<5×10^-8^, we also considered “suggestive” SNPs reaching p<1×10^-6^.

In order to explore the association of the genome-wide significant (and suggestive) DKI loci across the lifespan, we performed GWAS analyses with age-interaction (continuous variable), as implemented in REGENIE. For significant SNP-age interactions, we then extracted association results for the corresponding SNP in the different age strata defined by age-tertiles in the Rhineland Study (≤50, [50-62[, and >62 years). For genome-wide significant DKI-associated SNPs, we also performed sex-specific association analyses.

##### Association of DKI-associated loci in Chinese with DKI markers in the Rhineland Study

We explored associations with DKI markers in the Rhineland Study of loci identified in a recent GWAS in a Chinese population (N=4 183) [23]. For the 46 lead SNPs, if they were present in the Rhineland Study, we extracted their association results from the DKI GWAS results, using p<1.09×10^−3^ (0.05/46) as the significance threshold.

##### Association of DKI loci with DTI markers, cSVD, stroke, dementia and vascular risk factors

To assess the possible added value of DKI markers compared with classical DTI metrics in genetic analyses, we tested the association of suggestive (p<1×10^-6^) DKI loci with the two most studied DTI markers, FA and MD, in the Rhineland Study. In addition, to better characterize the identified DKI loci in relation to cSVD, we extracted results from these loci in the latest and much larger GWAS of DTI (FA and MD) markers (N = 33 292)[16], and we explored their association with cSVD, stroke, dementia and vascular risk factors, using the largest available European ancestry GWAS summary statistics of WMH (N = 50 970)[4], PVS (extensive PVS burden in white matter [PVS-WM], N = 9 607/39 822; in basal ganglia [PVS-BG] N = 9 189/40 000; and in hippocampus [PVS-HIP] N = 9 339/40 095)[8], ischemic stroke (N = 86 668/1 503 898)[31], Alzheimer disease (N = 111 326/677 663)[32], Parkinson disease (N = 33 674/449 056)[33], blood pressure (systolic [SBP], diastolic blood pressure [DBP] and pulse pressure [PP], N = 757 601)[34], lipids (LDL-, HDL-cholesterol, triglycerides [TG], N = 1 320 016)[35], body mass index (BMI) (N = 806 834)[36], and waist-to-hip ratio adjusted for BMI (WHR) (N = 694 649)[36]. Genetic associations at p<1.75x10^-4^ (Bonferroni-corrected threshold for 15 independent loci and 19 traits) were considered statistically significant.

##### Association of DKI loci with baseline and longitudinal cognitive performance in the Rhineland Study

We examined the association of suggestive DKI loci with six cognitive scores in the Rhineland Study. Associations were tested using similar regression models as for the DKI GWAS, adjusting for age, sex, and the first 10 principal components of population stratification. We then examined the association of suggestive DKI loci with the evolution of the six cognitive scores between baseline and the follow-up visit in the full sample. Associations were tested using a linear mixed model (in R, function lmer, lme4 package version 1.1-31) to model cognitive scores as a function of SNP*time interaction, time since baseline, age, age², sex, and the first ten principal components of population stratification, with a participant-specific random intercept to account for repeated measurements. Genetic associations reaching p<5.56×10^-4^ (correcting for 15 loci and 6 cognitive scores) were considered significant. However, considering the relatively limited sample size for these analyses, we only corrected for 15 independent loci (p<0.0033) and also considered nominally significant associations (p<0.05).

##### Association of cSVD MRI-markers (WMH, PVS), stroke, Alzheimer disease and Parkinson disease known risk loci with DKI and DTI markers

We explored associations with DKI markers in the Rhineland Study of known risk loci previously identified in European-ancestry populations for cSVD (WMH[4], PVS[8]), stroke[31], Alzheimer disease[32] and Parkinson disease[33]. We extracted association results of these 267 variants (232 independent loci at LD-r^2^>0.01) from DKI and DTI markers GWAS, using p<2.16×10^−4^ as the significance threshold.

##### Transcriptomic analyses

We performed transcriptome-wide association studies (TWAS) using TWAS-Fusion to identify genes whose expression is significantly associated with DKI markers [37]. We used precomputed functional weights from publicly available gene expression reference panels (expression quantitative trait loci [eQTL]) from tissues considered relevant for cerebrovascular disease (blood, arterial, heart, adipose, nerve and brain tissues) [38]. Transcriptome-wide significance at p<8.9x10^-6^ was based on the average number of features (5,608 genes) tested across tissues. These significant genes were then tested in conditional analyses in TWAS-Fusion [37]. Next, we performed a colocalization analysis (COLOC) on the conditionally significant genes (p<0.05) to estimate the posterior probability of a shared causal variant between the gene expression and trait association (PP4)[39], considering genes with PP4≥0.75 as colocalized. Colocalized genes with eQTLs reaching p<1×10^-5^ in association with the corresponding DKI marker (or in moderate-high LD, r²>0.5, with the lead SNP at p<1×10^-6^) were considered as being in a GWAS locus.

#### Proteomic analyses

##### Proteome-wide association study on individual-level data in Olink plasma proteomic data from the Rhineland Study

First, we studied the association of protein levels with DKI markers in the Rhineland Study. We used linear regression models (in R), adjusting for age at MRI, sex, total intracranial volume. We specifically extracted the results for the 62 proteins from GWAS significant and suggestive loci (Bonferroni corrected significant threshold of p<8.06x10^-4^ (0.05/62)).

Finally, to assess whether the suggestive DKI loci (lead SNPs) are associated with the level of the proteins located in GWAS loci and showing significant association with DKI markers in the Rhineland Study, we extracted their association results from the largest GWAS on protein levels in UK Biobank (Olink Explore 3072) [40]. SNPs with p<4.17x10^-4^ (correcting for 15 loci and 8 proteins) were considered significant.

##### Association of significant proteins with WMH, baseline and longitudinal cognitive performance in the Rhineland Study

To explore the clinical relevance of proteins associated with DKI markers also in GWAS loci, we tested their association in the Rhineland Study with the six cognitive domain scores and WMH. We used linear regression models (in R), adjusting for age at MRI and sex (and total intracranial volume for WMH). We then tested the association of these proteins with the evolution of the six cognitive domain scores between baseline and the follow-up visit. For cognitive decline, we used a linear mixed model (in R, function lmer, lme4 package version 1.1-31) to model cognitive scores as function of protein*time interaction, while adjusting for time since baseline, age, age² and sex, with a participant-specific random intercept to account for repeated measurements. Association at p<8.93x10^-4^ for analyses on GWAS loci (correcting for 8 proteins and 7 traits) were considered significant. For the latter, we also considered proteins reaching nominal significance (p<0.05).

## Results

### Genetic susceptibility to variations in DKI markers across the lifespan

Using GWAS for DKI markers in 5 930 participants from the Rhineland Study, we identified four genome-wide significant loci (p<5×10^−8^) associated with the three markers (AK, MK, RK): chr3p25.1 (intergenic variant between *LINC00620* and *WNT7A*), chr5q14.2 (in *VCAN*), chr5q14.3 (in *VCAN-AS1*) and chr8q24.21 (in *CCDC26*) (**Table 1**, **Figures S1-S2**). In addition, considering the relatively limited sample size for a GWAS, we also considered suggestive variants at p<1×10^−6^ (**Table S1**), thus identifying four additional loci for AK (chr7p22.3, chr7p11.2, chr16q12.2, chr17q21.31), five for MK (chr5q14.2, chr9q31.1, chr10q25.1, chr17q21.31, chr22q13.31), and six for RK (chr5q14.2, chr5q14.3, chr10q22.3, chr10q25.1, chr17q25.1, chr22q13.31). To explore the association of the genome-wide significant (and suggestive) DKI loci across the lifespan, we then tested the SNP-age interaction of these loci (**Table S1**). Only rs3852188 (chr5q14.3) showed significant interaction with age for all DKI markers. We performed age-stratified analyses for this SNP in the Rhineland Study (≤50, [50-62[, and >62 years) and observed weaker and non-significant associations in the older age strata than in the two others (**Table S2**). In addition, for genome-wide significant DKI-associated SNPs, we performed sex-specific association analyses. Sex-stratified results were comparable in men and women for the four loci, with the same directions of effect and overlapping confidence intervals, except for a weaker (and non-significant for RK) association of rs3852188 (chr5q14.3) in men than women (**Figure S3**).

**Table 1:** GWAS results - DKI genome-wide significant results.

| SNP | Locus | POS | Function | Nearest genes | EA | NEA | EAF | Beta | SE | p |
| --- | --- | --- | --- | --- | --- | --- | --- | --- | --- | --- |
| **Axial kurtosis** |  |  |  |  |  |  |  |  |  |  |
| rs6442411 | 3p25.1 | 13836296 | intergenic | *LINC00620;WNT7A* | T | C | 0.39 | 0.11 | 0.02 | 8.01E-12 |
| rs12332199 | 5q14.2 | 82786194 | exonic | *VCAN* | T | C | 0.63 | 0.13 | 0.02 | 1.08E-14 |
| rs3852188 | 5q14.3 | 82858014 | ncRNA exonic | *VCAN-AS1* | C | G | 0.80 | 0.21 | 0.02 | 1.02E-23 |
| rs147958197 | 8q24.21 | 130631395 | ncRNA intronic | *CCDC26* | T | C | 0.96 | 0.25 | 0.04 | 5.05E-09 |
| **Mean kurtosis** |  |  |  |  |  |  |  |  |  |  |
| rs6442411 | 3p25.1 | 13836296 | intergenic | *LINC00620;WNT7A* | T | C | 0.39 | 0.12 | 0.02 | 1.94E-13 |
| rs11749904 | 5q14.2 | 82782530 | intronic | *VCAN* | T | C | 0.64 | 0.13 | 0.02 | 6.21E-14 |
| rs3852188 | 5q14.3 | 82858014 | ncRNA exonic | *VCAN-AS1* | C | G | 0.80 | 0.21 | 0.02 | 6.57E-23 |
| rs147958197 | 8q24.21 | 130631395 | ncRNA intronic | *CCDC26* | T | C | 0.96 | 0.25 | 0.04 | 1.14E-08 |
| **Radial kurtosis** |  |  |  |  |  |  |  |  |  |  |
| rs6442411 | 3p25.1 | 13836296 | intergenic | *LINC00620;WNT7A* | T | C | 0.39 | 0.12 | 0.02 | 3.74E-12 |
| rs11749904 | 5q14.2 | 82782530 | intronic | *VCAN* | T | C | 0.64 | 0.12 | 0.02 | 3.36E-12 |
| rs3852188 | 5q14.3 | 82858014 | ncRNA exonic | *VCAN-AS1* | C | G | 0.80 | 0.19 | 0.02 | 9.33E-20 |
| rs147958197 | 8q24.21 | 130631395 | ncRNA intronic | *CCDC26* | T | C | 0.96 | 0.24 | 0.04 | 3.21E-08 |

POS: Position (build 37); EA: Effect allele; NEA: Non-effect allele; EAF: Effect allele frequency; Beta: Regression coefficient for EA; SE: Standard error for Beta; p: p-value.

We looked-up the lead SNPs identified in a recent GWAS of DKI markers from a Chinese population in the GWAS results from the Rhineland Study. Of the 46 SNPs tested, 19 were found in the Rhineland Study. Of these, only SNPs at chr5q14.2 and chr5q14.3 reached (genome-wide) significance (**Table S3**).

### Genetic relationship of DKI and DTI markers

We then tested the association of genome-wide significant and suggestive (p<1×10^-6^) DKI loci with the two most studied DTI markers, FA and MD, in the Rhineland Study and in the latest GWAS of DTI markers [16]. The four genome-wide significant DKI loci were genome-wide significantly associated with at least one DTI marker in both the Rhineland Study and the published GWAS, except chr8q24.21 which was not genome-wide significant in the latter. Among the 11 additional suggestive DKI loci, two were genome-wide significant in the published GWAS (chr7p22.3 and chr7p11.2 with MD), and most of them were associated with DTI markers in the Rhineland Study at p<1.75×10^-4^ (**Figure S4, Table S4**).

To assess the relation between cSVD, stroke and dementia risk loci and DKI (compared to DTI) markers, we extracted the association results of these risk loci in the GWAS of DKI and DTI in the Rhineland Study (**Table 2**, **Table S5**). After correction for multiple testing, three Alzheimer disease loci (chr6p21.32, chr7p11.2 and chr17q21.31), and one Parkinson disease locus (chr17q21.31) were associated with DKI markers. PVS-WM loci at chr3p25.1 and chr5q14.3 WMH locus were associated with DKI and DTI markers. Of note, only one locus, chr9q31.3 (PVS-WM) was associated only with MD.

**Table 2:**
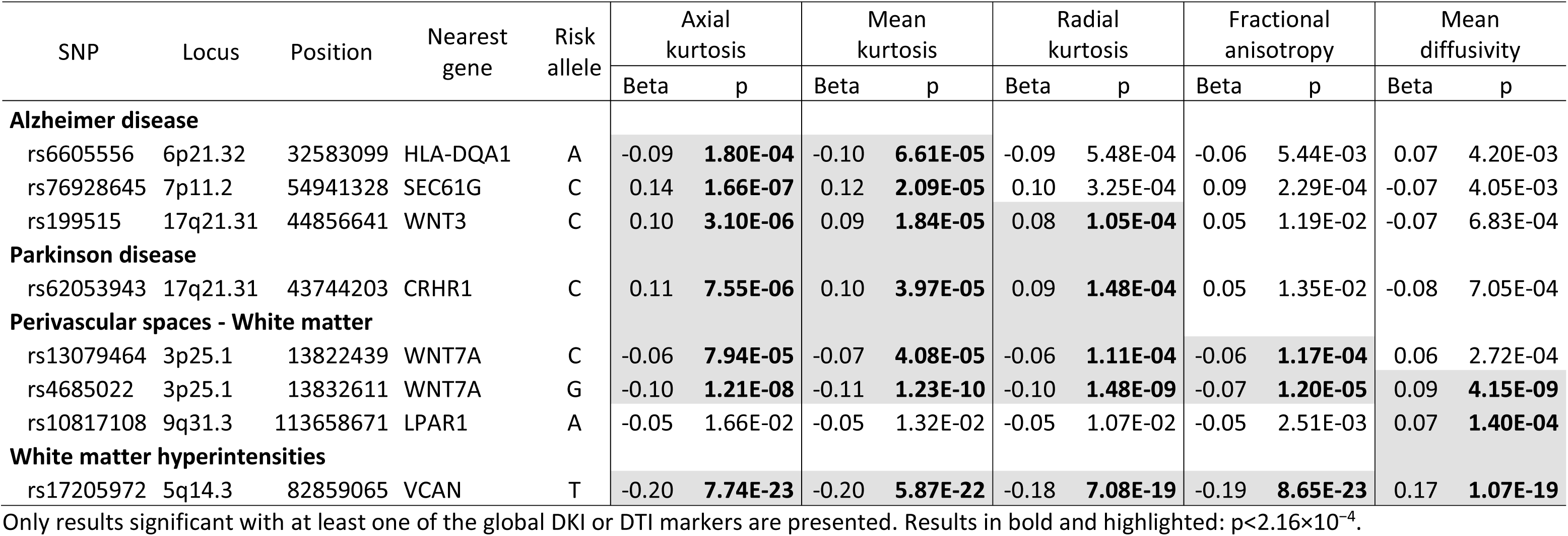
Significant association of loci associated with neurological traits with DKI and DTI markers.

| SNP | Locus | Position | Nearest gene | Risk allele | Axial kurtosis |  | Mean kurtosis |  | Radial kurtosis |  | Fractional anisotropy |  | Mean diffusivity |  |
| --- | --- | --- | --- | --- | --- | --- | --- | --- | --- | --- | --- | --- | --- | --- |
|  |  |  |  |  | Beta | p | Beta | p | Beta | p | Beta | p | Beta | p |
| Alzheimer disease |  |  |  |  |  |  |  |  |  |  |  |  |  |  |
| rs6605556 | 6p21.32 | 32583099 | HLA-DQA1 | A | -0.09 | 1.80E-04 | -0.10 | 6.61E-05 | -0.09 | 5.48E-04 | -0.06 | 5.44E-03 | 0.07 | 4.20E-03 |
| rs76928645 | 7p11.2 | 54941328 | SEC61G | C | 0.14 | 1.66E-07 | 0.12 | 2.09E-05 | 0.10 | 3.25E-04 | 0.09 | 2.29E-04 | -0.07 | 4.05E-03 |
| rs199515 | 17q21.31 | 44856641 | WNT3 | C | 0.10 | 3.10E-06 | 0.09 | 1.84E-05 | 0.08 | 1.05E-04 | 0.05 | 1.19E-02 | -0.07 | 6.83E-04 |
| Parkinson disease |  |  |  |  |  |  |  |  |  |  |  |  |  |  |
| rs62053943 | 17q21.31 | 43744203 | CRHR1 | C | 0.11 | 7.55E-06 | 0.10 | 3.97E-05 | 0.09 | 1.48E-04 | 0.05 | 1.35E-02 | -0.08 | 7.05E-04 |
| Perivascular spaces - White matter |  |  |  |  |  |  |  |  |  |  |  |  |  |  |
| rs13079464 | 3p25.1 | 13822439 | WNT7A | C | -0.06 | 7.94E-05 | -0.07 | 4.08E-05 | -0.06 | 1.11E-04 | -0.06 | 1.17E-04 | 0.06 | 2.72E-04 |
| rs4685022 | 3p25.1 | 13832611 | WNT7A | G | -0.10 | 1.21E-08 | -0.11 | 1.23E-10 | -0.10 | 1.48E-09 | -0.07 | 1.20E-05 | 0.09 | 4.15E-09 |
| rs10817108 | 9q31.3 | 113658671 | LPAR1 | A | -0.05 | 1.66E-02 | -0.05 | 1.32E-02 | -0.05 | 1.07E-02 | -0.05 | 2.51E-03 | 0.07 | 1.40E-04 |
| White matter hyperintensities |  |  |  |  |  |  |  |  |  |  |  |  |  |  |
| rs17205972 | 5q14.3 | 82859065 | VCAN | T | -0.20 | 7.74E-23 | -0.20 | 5.87E-22 | -0.18 | 7.08E-19 | -0.19 | 8.65E-23 | 0.17 | 1.07E-19 |

Only results significant with at least one of the global DKI or DTI markers are presented. Results in bold and highlighted: $p < 2.16 \times 10^{-4}$ .

### Clinical correlates of DKI-associated loci

To better characterize the identified DKI loci, we extracted results from suggestive (p<1×10^-6^) DKI loci with cSVD MRI-markers, stroke, dementia and vascular risk factors. Alleles associated with decreased DKI were associated with increased WMH volume at chr5q14.3 (p=1.13×10^-11^) and chr17q25.1 (p=8.35×10^-16^), with high PVS-WM burden (p=6.95×10^-8^) at chr3p25.1, and lower risk of Alzheimer disease at chr7p11.2 (p=1.51×10^-8^). Interestingly, the SNP at chr17q25.1 was also associated (p<1.75×10^-4^) with blood pressure, LDL-cholesterol and BMI (**Figure S4, Table S4**).

Five of the 15 suggestive DKI loci were significantly (p<0.05, none after correction for multiple testing) associated with cognitive performance in the Rhineland Study (**Figure S5, Table S6**). Of these, DKI-increasing allele at chr5q14.3 (*VCAN-AS1*) was associated with a negative evolution of executive function; chr7p11.2 with DKI-increasing allele associated with worst baseline performance for working memory and longer reaction time, and favorable evolution of global cognition, episodic verbal memory and executive function.

### Transcriptomic determinants of DKI markers

Next, to explore the possible causal genes underlying GWAS associations, we performed TWAS on DKI markers using TWAS-Fusion and eQTLs from external RNA sequencing resources in relevant tissues. We identified 21 genes (15 with AK, 14 with MK and 5 with RK) whose genetically predicted expression was associated with DKI markers (p<8.9x10^-6^ and p<0.05 in conditional analyses) in at least one tissue of which 17 presented colocalization of GWAS SNP and eQTL (COLOC-PP4>0.75), including 14 at the chr17q21.31 suggestive GWAS locus expressed in different tissues: *ARHGAP27*, *ARL17A*, *C17orf104*, *CRHR1-IT1*, *FAM215B*, *KANSL1-AS1*, *LRRC37A4P*, *PLEKHM1*, *RP11-259G18.1*, *RP11-259G18.2*, *RP11-259G18.3*, *RP11-707O23.5*, *RP11-798G7.8*, *WNT3* (**Figure 2**, **Figure S6, Table S7**). Of note, *VCAN* expression in arterial tissue, although eQTL was not in LD with GWAS lead SNPs, was significantly associated with the three DKI markers in TWAS analyses, but not colocalized (COLOC-PP4=0.5).

**Figure 2:**
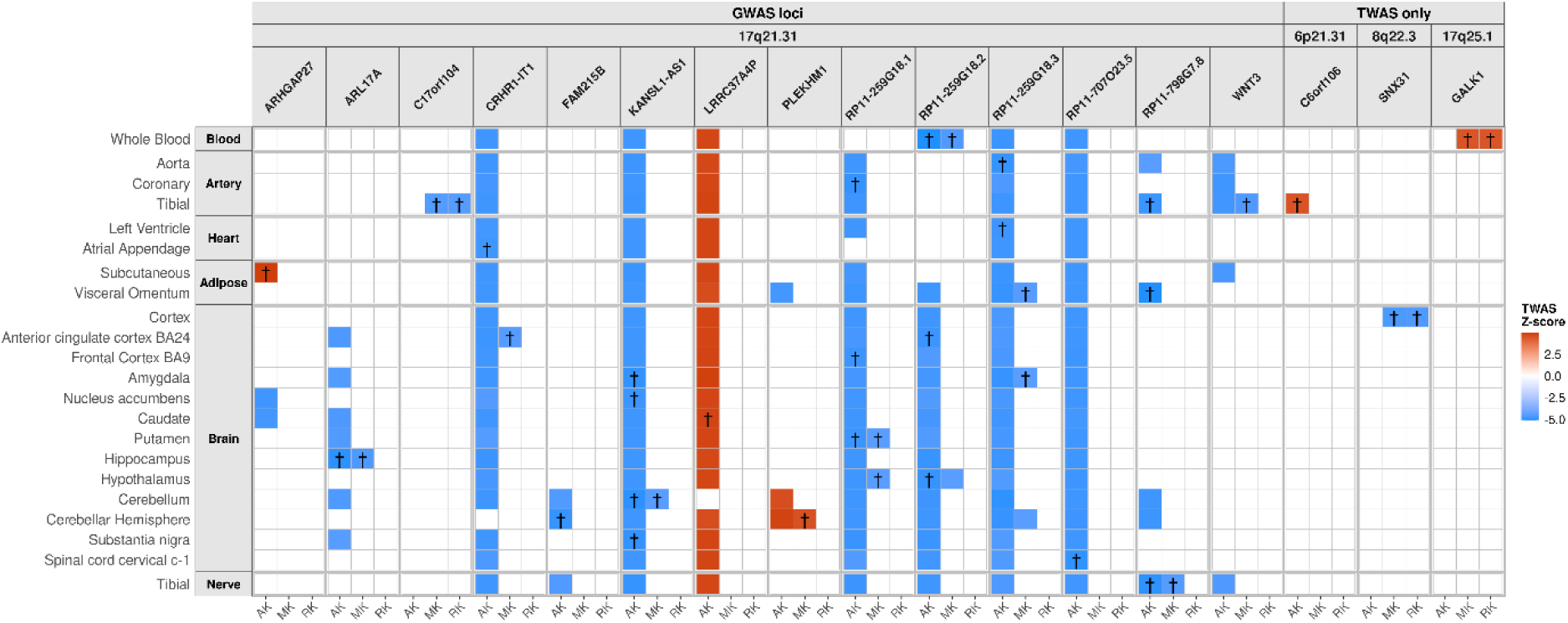
Transcriptome-wide association study (TWAS) of DKI markers in multiple tissues. Only genes with significant TWAS (p<8.9x10^-6^) and conditional (p<0.05) association, and colocalized (COLOC-PP4≥0.75) with at least one DKI marker are shown. Only results with TWAS p<8.9x10^-6^ are colored; *: TWAS p<8.9x10^-6^, p<0.05 in conditional analyses; †: TWAS p<8.9x10^-6^, p<0.05 in conditional analyses and COLOC-PP4≥0.75. AK: Axial kurtosis; MK: Mean kurtosis; RK: Radial kurtosis.

### Proteomic determinants of DKI markers

When studying proteins in loci identified in the GWAS of DKI markers, we identified eight unique proteins whose plasma levels were associated with DKI markers (8.06x10^-4^), chr5q14.2 (VCAN), chr7p11.2 (EGFR), chr7p22.3 (MAD1L1), chr16q12.2 (CES1), chr17q21.31 (GFAP), chr17q25.1 (CD300A, CD300C, CD300E) (**Figure 3**, **Table S8**). Of these, three were significantly (p<8.93x10^-4^) associated with WMH at baseline or at least one of the longitudinal cognitive scores in the Rhineland Study (**Figure 3**, **Table S9**). Higher MAD1L1 level was associated with lower DKI, higher WMH volume and decline in global cognition and episodic verbal memory. Lower EGFR was associated with lower DKI and cognitive decline (global cognition). Finally, higher GFAP was associated with lower DKI and decline in all cognitive domains (at p<0.05 for working memory).

**Figure 3:**
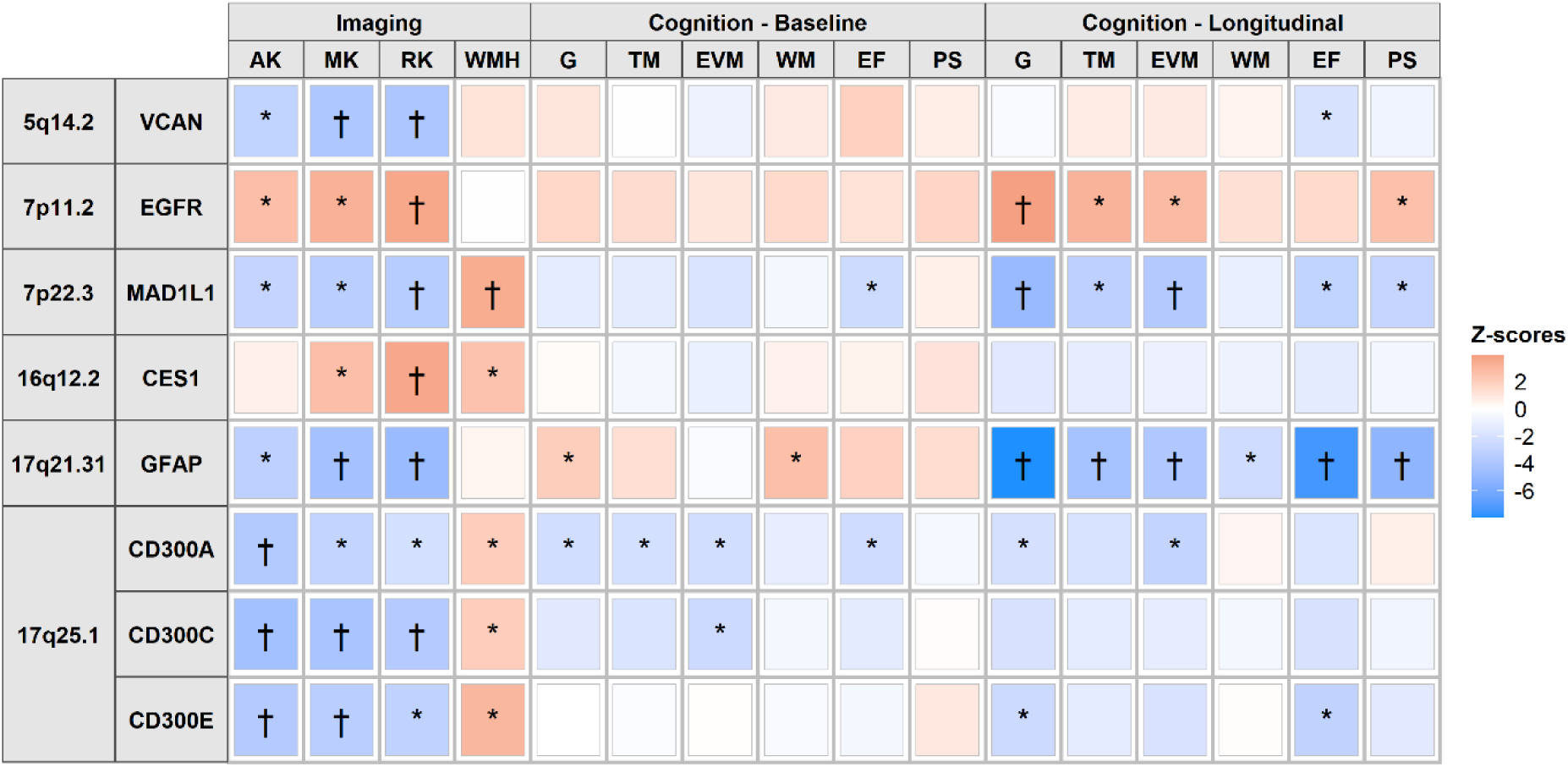
Association of proteins in DKI GWAS loci with DKI markers, cognition and WMH in the Rhineland Study. Only proteins associated with at least one DKI marker after correction for multiple testing (8.06x10^-4^) are shown. *: p<0.05; †: p<8.06x10^-4^ with DKI or p<8.93x10^-4^ with cognition-WMH. AK: Axial kurtosis; MK: Mean kurtosis; RK: Radial kurtosis; G: Global cognition; TM: Total memory; EVM: Episodic verbal memory; WM: Working memory; EF: Executive function; PS: Processing speed; WMH: White matter hyperintensities.

The DKI GWAS lead SNPs at chr5q14.3 exhibited a significant positive association (p<5x10^-8^) with VCAN levels in UK Biobank, while two additional DKI suggestive SNPs at chr5q14.2 were negatively associated (p<4.17x10^-4^) with VCAN. The DKI GWAS suggestive SNP at chr7p11.2 was negatively associated with GFAP level in UK Biobank (**Figure S7**).

## Discussion

In this first multi-omics study of DKI markers, we identified several novel genomic, transcriptomic and proteomic determinants of brain white matter microstructure in 5 930 participants from the Rhineland Study and explored their relation with cSVD and dementia. We identified four genome-wide significant loci associated with DKI markers at chr3p25.1 (*LINC00620*-*WNT7A*), chr5q14.2 (*VCAN*), chr5q14.3 (*VCAN-AS1*) and chr8q24.21 (*CCDC26*), and 11 additional suggestive DKI loci (p<1×10^-6^). Lead SNPs at chr5q14.3 and chr17q25.1 were associated with WMH volume, chr3p25.1 with PVS-WM, and chr7p11.2 with Alzheimer disease. Using TWAS, we identified 17 genes with genetically determined expression associated with DKI markers, including 14 at the chr17q21.31 suggestive GWAS locus. Finally, we identified eight proteins associated with DKI markers in GWAS suggestive loci. Of these, MAD1L1, EGFR and GFAP were also associated with cognitive decline, and MAD1L1 with WMH.

Individually, the two most significant genetic associations with DKI markers were observed for two independent loci at chr5q14.2 and, primarily, chr5q14.3, the known white matter microstructure (DTI and neurite orientation dispersion and density imaging markers) and WMH locus in *VCAN* (chr5q14.3) [4, 41, 42]. The lead SNP was also associated with versican plasma levels in UK Biobank [40], and VCAN protein levels were associated with DKI markers in the Rhineland Study. This locus is also the only one replicated from the recent DKI GWAS from a Chinese population [23]. *VCAN* encodes versican, a chondroitin sulfate proteoglycan and a major component of the extracellular matrix, playing a key role in tissue morphogenesis and in regulating immunity and inflammation [43]. *VCAN* is also one of the main hubs across the brain vascular matrisome,[44] which seem to play a central role in cSVD pathophysiology [9]. Versican was also found to be involved in remyelination processes in multiple sclerosis [45]. Our TWAS results showed significant association between genetically predicted *VCAN* expression in tibial artery and the three DKI markers, but did not colocalize with GWAS signals (COLOC-PP4=0.5). In the human brain, *VCAN* is mainly expressed in oligodendrocyte precursor cells and, to a lesser extent, astrocytes and ependymal cells [46]. The *VCAN* locus was the only one showing a significant interaction with age in the DKI GWAS, with association in older adults being non-significant. These results are in line with previous observations that *VCAN* expression is highest in the prenatal stages, and decreases over the life course [41, 47]. Further experimental follow-ups are needed to decipher the role of *VCAN* in cSVD.

The second most significant locus in GWAS analyses of DKI markers was an intergenic variant at chr3p25.1 (*LINC00620*-*WNT7A*), also associated with DTI-MD in both the Rhineland Study and a previous larger GWAS[16], and PVS-WM burden in the latest GWAS (p=6.95x10^-8^)[8]. This variant is in strong LD with the PVS-WM GWAS lead SNP rs4685022 (LD-r²=0.79)[8], which is an eQTL of *WNT7A* in brain tissues [38]. *WNT7A* is a member of the WNT gene family. These genes encode secreted signaling proteins regulating developmental central nervous system angiogenesis [48]. *WNT7A* encodes a protein directly targeting the vascular endothelium and involved in regulating brain angiogenesis and blood-brain barrier [48–50]. A study also showed that loss of Wnt7a/b function in mice resulted in severe white matter damage [51]. In the human brain, *WNT7A* is mainly expressed in astrocytes and, to a lesser extent, oligodendrocyte precursor cells, ependymal cells and neurons [46]. Interestingly, TWAS analyses showed that higher genetically predicted expression levels in arterial tissues of another member of the WNT family, *WNT3* in chr17q21.31, were associated with lower DKI values. *WNT3* expression in arterial tissues was also previously associated with increased PVS burden in basal ganglia [8]. Together, these results suggest an important role of this family of genes in cSVD pathophysiology. The canonical Wnt-β-catenin pathway, involved in regulating blood-brain barrier integrity, in response to vascular lesions, in oligodendrogenesis and myelination, has been suggested as a relevant pathway in cSVD and Alzheimer disease [9, 52–55]. More work is needed to fully understand the functional role of this pathway in the pathophysiology of these diseases.

An additional locus reached genome-wide significance with DKI markers, an intronic variant at chr8q24.21 in *CCDC26*. DKI-increasing allele of the lead variant at this locus was also associated (p<5x10^-8^) with increased DTI-FA in the Rhineland Study. This variant was previously reported as associated with specific regions of interest in UK Biobank (N = 33 000) using DTI and other dMRI metrics [42]. *CCDC26* is a long non-coding RNA gene. In the human brain, *CCDC26* is mainly expressed in microglia [46]. *CCDC26* has been shown to be involved in glioma, with the DKI-associated lead variant being in high LD with the probably causal low-grade glioma risk variant rs55705857 (LD-r²=0.82) [56, 57].

Although only reaching the suggestive threshold in GWAS analyses with AK and MK, but nearly significant with AK (p=5.71x10^-8^), the chr17q21.31 locus presented interesting results in clinical follow-ups and omics analyses. Indeed, Alzheimer disease and Parkinson disease GWAS lead variants, in moderate LD with the DKI lead SNP (respectively LD-r²=0.35 and 0.25), are located in this locus and showed significant associations with DKI markers (p<2.16×10^−4^). In addition, in TWAS analyses, we identified 14 genes at this locus with genetically determined expression in vascular or brain tissues associated with DKI markers with evidence of colocalization. Among these genes, *CRHR1-IT1* and *KANSL1-AS1* were previously associated with WMH [4], and *RP11-259G18.1*, *RP11-259G18.2*, *RP11-259G18.3* and the aforementioned *WNT3* gene with PVS in basal ganglia [8]. Finally, in proteomic analyses, higher GFAP (Glial Fibrillary Acidic Protein) levels were significantly associated with lower DKI, in line with previous findings with DTI [58], and with decline in all cognitive domains. GFAP is a major intermediate filament proteins of mature astrocytes, and is highly expressed in astrocytes [46]. Mutations in the *GFAP* gene are known to cause Alexander disease, a rare neurodegenerative disease affecting astrocytes and leading to psychomotor regression and death. Interestingly, a recent study showed that blood GFAP levels correlate with Alzheimer disease neuropathology, supporting its role as a diagnostic biomarker and providing insights about the role of astrocyte reactivity in Alzheimer disease [59]. Higher circulating GFAP levels have previously been shown to be associated with lower general cognition and higher risk of dementia [60]. In a previous study, plasma GFAP levels were associated with WMH in older adults [61]. However, we did not confirm this association in our sample. Finally, a recent study in a Japanese population showed an association of a missense variant in *GFAP* (common in Japanese people, rare in non-Asian populations) with WMH [62]. Thus, these results may suggest an important role of this locus in the pathophysiology of both cSVD and dementia.

Expression in the human brain of the three main genes in the most significant DKI loci (*VCAN*, *WNT7A, CCDC26*) was primarily in oligodendrocyte precursor cells, astrocytes and ependymal cells (*VCAN*, *WNT7A*), and microglia (*CCDC26*). Previous studies have shown an enrichment of genes within WMH risk loci in several cell types, including oligodendrocytes, oligodendrocyte precursors, ependymal cells and astrocytes [4, 9]. These and GFAP results suggest an important role of glial cells, and particularly astrocytes, in white matter microstructure variability, possibly related to cSVD.

Several studies have shown that DKI can overcome some limitations of DTI and allow to differentiate cognitively normal individuals and patients with Alzheimer disease [63, 64]. Suggestive DKI GWAS loci were associated with cSVD MRI-markers and Alzheimer disease. The lead SNP at chr17q25.1 was associated with increased DKI and smaller WMH volume[4] (in moderate LD [r^2^=0.36] with the known WMH *TRIM65* locus). The lead SNP at chr7p11.2 (*SEC61G-DT*; *EGFR*) was associated with increased DKI and higher risk for Alzheimer disease. This SNP was also associated with lower GFAP (chr17q21.31) protein level in the UK Biobank. A similar unexpected direction of effect with dementia was also observed for the Alzheimer and Parkinson risk locus at chr17q21.31 (in *CRHR1* and *WNT3*). Finally, we showed that lower EGFR protein levels were associated with lower DKI and cognitive decline in the Rhineland Study. Overall, these findings suggest a complex relationship between DKI markers, cognition and neurodegenerative diseases at these loci. Further studies are needed to confirm these associations.

Although most DKI loci were associated with DTI markers, an interesting finding of this study is that, at similar sample size, known cSVD loci were similarly associated with DKI and DTI markers, but neurodegenerative diseases loci were only associated with DKI markers. This could suggest a greater sensitivity of DKI markers to neurodegenerative pathways [18]. Thus, our results suggest that, combined with other omics data, DKI markers may capture cSVD- and neurodegenerative-related variations in white matter microstructure, or their complex interplay. However, additional studies in independent large population-based cohorts are needed to better understand these results.

To our knowledge, this study is the first exploring molecular determinants of the emerging DKI markers. We used a single large homogeneous population-based cohort with a wide age range and, to our knowledge, the only population-based cohort of European ancestry participants with DKI and omics data available. We combined extensive high-quality multi-b-value MRI measures, neuropsychological tests, individual-level omics data, as well as publicly available external resources to comprehensively characterize molecular determinants of these cutting-edge dMRI markers. We acknowledge limitations. Our cohort was of limited sample size for omics analyses. Due to unavailability of other cohorts with similar individual-level omics data, we were not able to replicate our findings in an independent sample. However, to strengthen our findings we conducted many look-ups on related traits (including DTI, and cSVD MRI-markers) from external publicly available data. Further omics studies on DKI will be crucial to confirm our findings. Finally, study participants were all of European ancestry, and from a privileged region of the world, thus limiting the generalizability of our results.

In summary, leveraging omics data, our study identified novel molecular determinants of white matter microstructure using DKI markers, providing important novel insights into life course determinants of cSVD, a leading cause of stroke and dementia. Our results showed interesting associations of *VCAN*, *WNT7A* and *CCDC26* loci, many genes at chr17q21.31 (including *WNT3*), and of GFAP plasma levels with DKI markers, as well as intriguing associations of several DKI-associated loci and proteins with cognitive decline and neurodegenerative diseases. Moreover, our results suggest a potentially important role of glial cells, particularly astrocytes, in white matter microstructure variability. Further research is warranted to decipher the molecular pathways and mechanisms underlying the relation of DKI markers, cSVD and dementia.

## Acknowledgements

We would like to thank all participants of the Rhineland Study and the study personnel involved in the extensive data collection.

## Data availability

The data from the Rhineland Study are not publicly available due to data protection regulations. Access to data can be provided to scientists in accordance with the Rhineland Study’s Data Use and Access Policy. Requests for additional information or access to the Rhineland Study’s datasets can be sent to https://RS-DUAC@dzne.de.

We used publicly available resources in this manuscript, including data from GTEx (https://gtexportal.org/home/), the Gusev laboratory (http://gusevlab.org/projects/fusion/), OMIM (https://www.omim.org/); and publicly available GWAS summary statistics for plasma protein levels in UK Biobank (https://www.synapse.org/Synapse:syn51364943/wiki/622119), brain MRI traits in UK Biobank (https://open.oxcin.ox.ac.uk/ukbiobank/big40/), WMH (https://www.ncbi.nlm.nih.gov/gap/, phs002227.v1.p1), PVS (https://www.ebi.ac.uk/gwas/, GCST90244151, GCST90244153, GCST90244155), ischemic stroke (https://www.ebi.ac.uk/gwas/, GCST90104540), Parkinson disease (https://drive.google.com/drive/folders/10bGj6HfAXgl-JslpI9ZJIL_JIgZyktxn), Alzheimer disease (https://www.ebi.ac.uk/gwas/, GCST90027158), blood pressure (https://www.ebi.ac.uk/gwas/, GCST006624,GCST006630,GCST006629), lipid traits (https://csg.sph.umich.edu/willer/public/glgc-lipids2021/), BMI and WHR (https://portals.broadinstitute.org/collaboration/giant/index.php/GIANT_consortium_data_ files).

## References

1. Alber J, Alladi S, Bae H-J, Barton DA, Beckett LA, Bell JM, et al. White matter hyperintensities in vascular contributions to cognitive impairment and dementia (VCID): Knowledge gaps and opportunities. Alzheimers Dement (N Y). 2019;5:107–117.

2. Iadecola C, Duering M, Hachinski V, Joutel A, Pendlebury ST, Schneider JA, et al. Vascular Cognitive Impairment and Dementia: JACC Scientific Expert Panel. J Am Coll Cardiol. 2019;73:3326–3344.

3. Debette S, Schilling S, Duperron M-G, Larsson SC, Markus HS. Clinical Significance of Magnetic Resonance Imaging Markers of Vascular Brain Injury: A Systematic Review and Meta-analysis. JAMA Neurol. 2019;76:81–94.

4. Sargurupremraj M, Suzuki H, Jian X, Sarnowski C, Evans TE, Bis JC, et al. Cerebral small vessel disease genomics and its implications across the lifespan. Nat Commun. 2020;11:6285.

5. Chauhan G, Adams HHH, Satizabal CL, Bis JC, Teumer A, Sargurupremraj M, et al. Genetic and lifestyle risk factors for MRI-defined brain infarcts in a population-based setting. Neurology. 2019;92(5):10.1212/WNL.0000000000006851.

6. Wardlaw JM, Benveniste H, Nedergaard M, Zlokovic BV, Mestre H, Lee H, et al. Perivascular spaces in the brain: anatomy, physiology and pathology. Nat Rev Neurol. 2020;16:137–153.

7. Wardlaw JM, Smith EE, Biessels GJ, Cordonnier C, Fazekas F, Frayne R, et al. Neuroimaging standards for research into small vessel disease and its contribution to ageing and neurodegeneration. Lancet Neurol. 2013;12:822–838.

8. Duperron M-G, Knol MJ, Le Grand Q, Evans TE, Mishra A, Tsuchida A, et al. Genomics of perivascular space burden unravels early mechanisms of cerebral small vessel disease. Nat Med. 2023;29:950–962.

9. Bordes C, Sargurupremraj M, Mishra A, Debette S. Genetics of common cerebral small vessel disease. Nat Rev Neurol. 2022;18:84–101.

10. Baykara E, Gesierich B, Adam R, Tuladhar AM, Biesbroek JM, Koek HL, et al. A Novel Imaging Marker for Small Vessel Disease Based on Skeletonization of White Matter Tracts and Diffusion Histograms. Ann Neurol. 2016;80:581–592.

11. Konieczny MJ, Dewenter A, Ter Telgte A, Gesierich B, Wiegertjes K, Finsterwalder S, et al. Multi-shell Diffusion MRI Models for White Matter Characterization in Cerebral Small Vessel Disease. Neurology. 2021;96:e698–e708.

12. Jian X, Fornage M. Imaging Endophenotypes of Stroke as a Target for Genetic Studies. Stroke. 2018;49:1557–1562.

13. Maillard P, Carmichael O, Harvey D, Fletcher E, Reed B, Mungas D, et al. FLAIR and diffusion MRI signals are independent predictors of white matter hyperintensities. AJNR Am J Neuroradiol. 2013;34:54–61.

14. de Groot M, Verhaaren BFJ, de Boer R, Klein S, Hofman A, van der Lugt A, et al. Changes in normal-appearing white matter precede development of white matter lesions. Stroke. 2013;44:1037–1042.

15. Persyn E, Hanscombe KB, Howson JMM, Lewis CM, Traylor M, Markus HS. Genome-wide association study of MRI markers of cerebral small vessel disease in 42,310 participants. Nat Commun. 2020;11:2175.

16. Zhao B, Li T, Yang Y, Wang X, Luo T, Shan Y, et al. Common genetic variation influencing human white matter microstructure. Science. 2021;372:eabf3736.

17. Rutten-Jacobs LCA, Tozer DJ, Duering M, Malik R, Dichgans M, Markus HS, et al. Genetic Study of White Matter Integrity in UK Biobank (N=8448) and the Overlap With Stroke, Depression, and Dementia. Stroke. 2018;49:1340–1347.

18. Yang K, Wu Z, Long J, Li W, Wang X, Hu N, et al. White matter changes in Parkinson’s disease. NPJ Parkinsons Dis. 2023;9:150.

19. Vemuri P, Decarli C, Duering M. Imaging Markers of Vascular Brain Health: Quantification, Clinical Implications, and Future Directions. Stroke. 2022;53:416–426.

20. Jensen JH, Helpern JA, Ramani A, Lu H, Kaczynski K. Diffusional kurtosis imaging: the quantification of non-gaussian water diffusion by means of magnetic resonance imaging. Magn Reson Med. 2005;53:1432–1440.

21. Li T, Zhang Y, Fu X, Zhang X, Luo Y, Ni H. Microstructural white matter alterations in Alzheimer’s disease and amnestic mild cognitive impairment and its diagnostic value based on diffusion kurtosis imaging: a tract-based spatial statistics study. Brain Imaging Behav. 2022;16:31–42.

22. Kamagata K, Andica C, Hatano T, Ogawa T, Takeshige-Amano H, Ogaki K, et al. Advanced diffusion magnetic resonance imaging in patients with Alzheimer’s and Parkinson’s diseases. Neural Regen Res. 2020;15:1590–1600.

23. Fu J, Wang J, Xue H, Wang M, Zhang B, Zhu W, et al. Genome-wide association studies of brain diffusion kurtosis imaging phenotypes. EBioMedicine. 2026;127:106261.

24. Lohner V, Enkirch SJ, Hattingen E, Stöcker T, Breteler MMB. Safety of Tattoos, Permanent Make-Up, and Medical Implants in Population-Based 3T Magnetic Resonance Brain Imaging: The Rhineland Study. Front Neurol. 2022;13:795573.

25. Tobisch A, Stirnberg R, Harms RL, Schultz T, Roebroeck A, Breteler MMB, et al. Compressed Sensing Diffusion Spectrum Imaging for Accelerated Diffusion Microstructure MRI in Long-Term Population Imaging. Front Neurosci. 2018;12:650.

26. Koch A, Stirnberg R, Estrada S, Zeng W, Lohner V, Shahid M, et al. Versatile MRI acquisition and processing protocol for population-based neuroimaging. Nat Protoc. 2024. 13 December 2024. 10.1038/s41596-024-01085-w.

27. Li J, Ji L. Adjusting multiple testing in multilocus analyses using the eigenvalues of a correlation matrix. Heredity. 2005;95:221–227.

28. Coors A, Breteler MMB, Ettinger U. Processing speed, but not working memory or global cognition, is associated with pupil diameter during fixation. Psychophysiology. 2022;59:e14089.

29. Coors A, Imtiaz M-A, Boenniger MM, Aziz NA, Ettinger U, Breteler MMB. Associations of genetic liability for Alzheimer’s disease with cognition and eye movements in a large, population-based cohort study. Transl Psychiatry. 2022;12:337.

30. Mbatchou J, Barnard L, Backman J, Marcketta A, Kosmicki JA, Ziyatdinov A, et al. Computationally efficient whole-genome regression for quantitative and binary traits. Nat Genet. 2021;53:1097–1103.

31. Mishra A, Malik R, Hachiya T, Jürgenson T, Namba S, Posner DC, et al. Stroke genetics informs drug discovery and risk prediction across ancestries. Nature. 2022:1–15.

32. Bellenguez C, Küçükali F, Jansen IE, Kleineidam L, Moreno-Grau S, Amin N, et al. New insights into the genetic etiology of Alzheimer’s disease and related dementias. Nat Genet. 2022;54:412–436.

33. Nalls MA, Blauwendraat C, Vallerga CL, Heilbron K, Bandres-Ciga S, Chang D, et al. Identification of novel risk loci, causal insights, and heritable risk for Parkinson’s disease: a meta-analysis of genome-wide association studies. Lancet Neurol. 2019;18:1091–1102.

34. Evangelou E, Warren HR, Mosen-Ansorena D, Mifsud B, Pazoki R, Gao H, et al. Genetic analysis of over one million people identifies 535 new loci associated with blood pressure traits. Nat Genet. 2018;50:1412–1425.

35. Graham SE, Clarke SL, Wu K-HH, Kanoni S, Zajac GJM, Ramdas S, et al. The power of genetic diversity in genome-wide association studies of lipids. Nature. 2021;600:675– 679.

36. Pulit SL, Stoneman C, Morris AP, Wood AR, Glastonbury CA, Tyrrell J, et al. Meta-analysis of genome-wide association studies for body fat distribution in 694 649 individuals of European ancestry. Hum Mol Genet. 2019;28:166–174.

37. Gusev A, Ko A, Shi H, Bhatia G, Chung W, Penninx BWJH, et al. Integrative approaches for large-scale transcriptome-wide association studies. Nat Genet. 2016;48:245–252.

38. GTEx Consortium. The GTEx Consortium atlas of genetic regulatory effects across human tissues. Science. 2020;369:1318–1330.

39. Giambartolomei C, Vukcevic D, Schadt EE, Franke L, Hingorani AD, Wallace C, et al. Bayesian Test for Colocalisation between Pairs of Genetic Association Studies Using Summary Statistics. PLoS Genetics. 2014;10:e1004383.

40. Sun BB, Chiou J, Traylor M, Benner C, Hsu Y-H, Richardson TG, et al. Plasma proteomic associations with genetics and health in the UK Biobank. Nature. 2023;622:329–338.

41. Le Grand Q, Tsuchida A, Koch A, Imtiaz M-A, Aziz NA, Vigneron C, et al. Diffusion imaging genomics provides novel insight into early mechanisms of cerebral small vessel disease. Mol Psychiatry. 2024. 29 May 2024. 10.1038/s41380-024-02604-7.

42. Smith SM, Douaud G, Chen W, Hanayik T, Alfaro-Almagro F, Sharp K, et al. An expanded set of genome-wide association studies of brain imaging phenotypes in UK Biobank. Nat Neurosci. 2021;24:737–745.

43. Wight TN, Kang I, Evanko SP, Harten IA, Chang MY, Pearce OMT, et al. Versican-A Critical Extracellular Matrix Regulator of Immunity and Inflammation. Front Immunol. 2020;11:512.

44. Pokhilko A, Brezzo G, Handunnetthi L, Heilig R, Lennon R, Smith C, et al. Global proteomic analysis of extracellular matrix in mouse and human brain highlights relevance to cerebrovascular disease. J Cereb Blood Flow Metab. 2021;41:2423–2438.

45. Ghorbani S, Jelinek E, Jain R, Buehner B, Li C, Lozinski BM, et al. Versican promotes T helper 17 cytotoxic inflammation and impedes oligodendrocyte precursor cell remyelination. Nat Commun. 2022;13:2445.

46. Yang AC, Vest RT, Kern F, Lee DP, Agam M, Maat CA, et al. A human brain vascular atlas reveals diverse mediators of Alzheimer’s risk. Nature. 2022;603:885–892.

47. Kang HJ, Kawasawa YI, Cheng F, Zhu Y, Xu X, Li M, et al. Spatio-temporal transcriptome of the human brain. Nature. 2011;478:483–489.

48. Mancuso MR, Kuhnert F, Kuo CJ. Developmental Angiogenesis of the Central Nervous System. Lymphatic Research and Biology. 2008;6:173.

49. Stenman JM, Rajagopal J, Carroll TJ, Ishibashi M, McMahon J, McMahon AP. Canonical Wnt signaling regulates organ-specific assembly and differentiation of CNS vasculature. Science. 2008;322:1247–1250.

50. Manukjan N, Chau S, Caiment F, van Herwijnen M, Smeets HJ, Fulton D, et al. Wnt7a Decreases Brain Endothelial Barrier Function Via β-Catenin Activation. Mol Neurobiol. 2024;61:4854–4867.

51. Chavali M, Ulloa-Navas MJ, Pérez-Borredá P, Garcia-Verdugo JM, McQuillen PS, Huang EJ, et al. Wnt-Dependent Oligodendroglial-Endothelial Interactions Regulate White Matter Vascularization and Attenuate Injury. Neuron. 2020;108:1130–1145.e5.

52. Jia L, Piña-Crespo J, Li Y. Restoring Wnt/β-catenin signaling is a promising therapeutic strategy for Alzheimer’s disease. Mol Brain. 2019;12:104.

53. Couffinhal T, Dufourcq P, Duplàa C. Beta-catenin nuclear activation: common pathway between Wnt and growth factor signaling in vascular smooth muscle cell proliferation? Circ Res. 2006;99:1287–1289.

54. Guo F, Lang J, Sohn J, Hammond E, Chang M, Pleasure D. Canonical Wnt signaling in the oligodendroglial lineage--puzzles remain. Glia. 2015;63:1671–1693.

55. Palomer E, Buechler J, Salinas PC. Wnt Signaling Deregulation in the Aging and Alzheimer’s Brain. Front Cell Neurosci. 2019;13:227.

56. Shete S, Hosking FJ, Robertson LB, Dobbins SE, Sanson M, Malmer B, et al. Genome-wide association study identifies five susceptibility loci for glioma. Nat Genet. 2009;41:899– 904.

57. Yanchus C, Drucker KL, Kollmeyer TM, Tsai R, Winick-Ng W, Liang M, et al. A noncoding single-nucleotide polymorphism at 8q24 drives IDH1-mutant glioma formation. Science. 2022;378:68–78.

58. Bettcher BM, Olson KE, Carlson NE, McConnell BV, Boyd T, Adame V, et al. Astrogliosis and episodic memory in late life: higher GFAP is related to worse memory and white matter microstructure in healthy aging and Alzheimer’s disease. Neurobiol Aging. 2021;103:68–77.

59. Sánchez-Juan P, Valeriano-Lorenzo E, Ruiz-González A, Pastor AB, Rodrigo Lara H, López-González F, et al. Serum GFAP levels correlate with astrocyte reactivity, post-mortem brain atrophy and neurofibrillary tangles. Brain. 2024;147:1667–1679.

60. Gonzales MM, Wiedner C, Wang C-P, Liu Q, Bis JC, Li Z, et al. A population-based meta-analysis of circulating GFAP for cognition and dementia risk. Ann Clin Transl Neurol. 2022;9:1574–1585.

61. van Gennip ACE, Satizabal CL, Tracy RP, Sigurdsson S, Gudnason V, Launer LJ, et al. Associations of plasma NfL, GFAP, and t-tau with cerebral small vessel disease and incident dementia: longitudinal data of the AGES-Reykjavik Study. Geroscience. 2024;46:505–516.

62. Furuta Y, Akiyama M, Hirabayashi N, Honda T, Shibata M, Ohara T, et al. Common protein-altering variant in GFAP is associated with white matter lesions in the older Japanese population. NPJ Genom Med. 2024;9:59.

63. Nelson MR, Keeling EG, Stokes AM, Bergamino M. Exploring white matter microstructural alterations in mild cognitive impairment: a multimodal diffusion MRI investigation utilizing diffusion kurtosis and free-water imaging. Front Neurosci. 2024;18:1440653.

64. Struyfs H, Van Hecke W, Veraart J, Sijbers J, Slaets S, De Belder M, et al. Diffusion Kurtosis Imaging: A Possible MRI Biomarker for AD Diagnosis? J Alzheimers Dis. 2015;48:937–948.

