## Supplementary Materials for "Diffusion kurtosis imaging (gen)omics unravels mechanisms of cerebral small vessel disease"

#### *Supplement*

### Supplementary Methods

#### Study population

The Rhineland Study (www.rheinland-studie.de) is an ongoing community-based prospective cohort study that invites inhabitants aged 30 years and above at baseline living in two geographically defined areas in the city of Bonn, Germany. Persons living in those areas were predominantly German with Caucasian ethnicity. The sole exclusion criterion is insufficient German language skills to provide informed consent. Approval to undertake the study was obtained from the ethics committee of the Medical Faculty of the University of Bonn. The study is carried out in accordance with the recommendations of the International Council for Harmonization (ICH) Good Clinical Practice (GCP) standards (ICH-GCP). Written informed consent was obtained from all participants in accordance with the Declaration of Helsinki. We used baseline data for 5930 participants with both genotype data and MRI scans available (mean age ± standard deviation (SD): 55.6 ± 13.4 [range: 30-95] years; 58% women) after excluding participants with neurological disorders (dementia, multiple sclerosis, stroke and intracranial hemorrhage), of which 2233 also had proteomics. In addition, we performed age-stratified analyses using age strata defined by age-tertiles: ≤50 (N = 1 995), [50-62[ (N = 2 008), and >62 (N = 1 927) years.

#### MRI Acquisition and Phenotyping

The Rhineland Study MRI data was acquired at two examination sites in Bonn on 3 Tesla MRI scanners (MAGNETOM Prisma, Siemens Healthineers) equipped with a whole-body gradient system of 80 mT/m amplitude and 200 T/m/s slew rate and a 64-channel head-neck coil. The one-hour Rhineland Study MRI protocol included a 3D T1-weighted multi-echo magnetization-prepared rapid gradient-echo (ME-MPRAGE) sequence at 0.8 mm isotropic resolution (TA = 6.5 min, 4 TE between 1.7 and 6.5 ms, TR = 2560 ms, TI = 1100 ms, flip angle = 7°, FOV = 256 × 256 mm, 224 slices), a 3D T2-weighted turbo-spin-echo (TSE) sequence at 0.8 mm isotropic resolution (TA = 4.6 min, TR = 2800 ms, TE = 405 ms, FOV = 256 × 256 mm, 224 slices) and a 3D T2 fluid-attenuated inversion recovery (FLAIR) sequence at 1.0 mm isotropic resolution (TA = 4.5 min, TR = 5000 ms, TE = 393 ms, TI = 1800 ms, FOV = 256 × 256 mm, 176 slices) [1–3]. A compressed sensing diffusion spectrum imaging (CS-DSI) protocol was used to collect dMRI scans at 1.5 mm isotropic resolution (TA = 12.1 min, TR = 5500 ms, TE = 105 ms, FOV = 210 x

210 mm, 93 slices, multiband factor = 3, $b_{max}$ = 6800 s/mm$^2$, $\Delta$ = 51.3 ms, $\delta$ = 20.1 ms) with 4 pairs of AP and PA phase-encoded b = 0 s/mm$^2$ [4–6].

All T1-weighted images were processed using FreeSurfer version 6.0 (http://surfer.nmr.mgh.harvard.edu/) to derive volumetric segmentation and quantitative volumetric measures [7, 8]. The estimated total intracranial volume (eTIV) generated by FreeSurfer was used as a proxy for head size. White matter hyperintensities were segmented based on T1-weighted, T2-weighted and FLAIR images using an in-house developed pipeline [1]. Preprocessing steps for dMRI included the correction of susceptibility-induced and eddy current-induced distortions and head motion using FSL (www.fmrib.ox.ac.uk/fsl) and subsequent CS-DSI reconstruction of nonacquired diffusion spectrum samples [4, 9, 10]. The microstructure diffusion toolbox (MDT: https://github.com/robbert-harms/MDT) was used to estimate mean, axial and radial kurtosis from the diffusion kurtosis model as well as fractional anisotropy and mean diffusivity from the diffusion tensor model [11–13]. Subsequently, the standard tract-based spatial statistics (TBSS) framework in FSL was used to project all estimated dMRI parameters to a template FA skeleton [14]. We calculated the mean across voxels within the WM skeleton.

#### Cognitive assessment

The cognitive test battery of the Rhineland Study has been described in detail elsewhere [15]. We computed cognitive domain performance scores for the following domains: episodic verbal memory, working memory, executive function and processing speed. These scores were created by averaging the z scores of the tests contributing to each domain. Working memory was assessed with the orally performed Digit Span forward and backward task, and the touchpad-based Corsi block-tapping test, based on the Psychology Experiment Building Language battery [16]. Episodic verbal memory was evaluated with the Auditory Verbal Learning and Memory test with a list length of 15 words [17]. Assessment of processing speed was based on a numbers-only Trail-Making Test (Trail-Making Test, Part A), and prosaccade latency (time needed to initiate a saccade), derived from an eye movement test battery [18]. We examined executive function using the antisaccade error rate (percentage of trials in which the participant made a direction error) from the same battery, combined with a categorical word fluency task (animals), and a number-and-letters Trail-Making Test (Trail-Making Test, Part B). All examinations were administered in German language, following a standardized procedure by certified study technicians. Global cognition score was computed by averaging the z scores for episodic verbal memory, working memory, executive function, processing speed. Working memory and episodic verbal memory scores were further averaged to produce a total memory composite score. To normalize distributions, we applied a rank-based inverse normal transformation to cognitive variables.

#### Genotyping, quality control, and imputation

In the Rhineland study, DNA extracted from buffy coat samples were genotyped using Infinium Omni2.5Exome-8 BeadChip containing 2 612 357 SNPs and processed using GenomeStudio (version 2.0.5). Quality control of genotypes was performed using PLINK (version 1.9). Single-nucleotide polymorphisms (SNPs) exclusion criteria were Hardy-Weinberg disequilibrium ($p<1\times10^{-6}$), minor allele frequency (<0.01) and poor genotyping rate (<98%). Participants with potentially problematic samples were excluded, comprising cases with poor call rate (<95%), abnormal heterozygosity and gender mismatch. Since variation in population structure can cause systematic differences in allele frequencies[19], we used EIGENSTRAT (version 16000), which uses principal components (PCs) to detect and correct for variation in population structure [19]. Based on the EIGENSTRAT estimation, we excluded participants of non-Caucasian descent, retaining only participants from Caucasian descent for further analysis. We used the 1000 Genomes phase 3 reference panel [20] version 5 for the imputation of missing genotypes using impute2 (version 2) [21].

#### Plasma proteomics data

Out of the first 5 500 participants enrolled before April 2020, 2 852 participants were selected using an age- and sex-stratified random sampling strategy for proteomics analysis. Plasma protein profiling was conducted using the Olink Explore platform, which includes a comprehensive inbuilt quality control (QC) system that integrates both internal and external controls to ensure the reliability of assay performance and sample quality. Two main types of QC annotations were used in the received data output: QC warnings at the sample level and assay warnings at the protein level. A sample receives a QC_warning status of WARN if it fails to meet one or more of the following criteria: the average matched counts fall below 500, or the Incubation and Amplification Control values deviate by more than ±0.3 NPX from the plate median across the abundance block. Additionally, samples flagged through manual inspection are annotated with MANUAL_WARN in the same column. At the assay level, an Assay_warning of WARN is assigned if the median value of the triplicate Negative Controls for a given assay deviates by more than 5 standard deviations from a predefined threshold established during assay validation. As an initial filtering step, each batch was scanned for "Excluded" protein flags. For any given sample-panel combination, if a single protein was flagged as "Excluded", all protein measurements within that panel for the sample were removed. In total, 56 proteins were removed due to exclusion flags. In addition, outlier detection was performed using two complementary methods applied within each protein panel. The first method involved principal component analysis (PCA), in which samples with standardized PC1 or PC2 scores exceeding ±6 standard deviations from the panel mean were flagged. The second method evaluated the NPX distribution within each panel, flagging samples whose NPX median or interquartile range (IQR) exceeded ±6 SD. One sample consistently showed as outlier across all panels, then was excluded. Further quality control statistics were evaluated at both the protein and sample levels using the internal Olink warning flags (Assay_warning and QC_warning). Among the 2 888 protein assays in the final dataset, 123 protein assays (4.3%) were marked with Assay_warning. The QC_warning percentage across all assays ranged from 0% to 8.60%, with higher percentages concentrated in the Oncology II panel. Most assays had a QC Warning percentage of 0%, suggesting generally high technical reliability across panels. Only a small number of assays exceeded 5% QC Warnings, and these were exclusively observed in the Oncology II panel. Likewise, Assay Warnings were absent (0%) for the majority of proteins, with only a limited subset showing elevated percentages. When restricting to proteins with Assay Warning Percentage ≥ 5%, warnings remained sparse but were more frequently observed in the Oncology II and Inflammation II panels.

#### Statistical analyses

##### *Genome-wide association study (GWAS)*

We performed GWAS using the genome-wide linear mixed model implemented in REGENIE [22]. REGENIE computation is composed of two steps. The step 1 uses a subset of genetic markers to fit a whole genome regression model that captures a good fraction of the phenotype variance attributable to genetic effects. We used a pruned subset of genotyped SNPs using a LD-$r^2$ threshold of 0.6 with a window size of 1 000 and a step size of 100 markers.

The step 2 uses a larger set of genetic markers and tests them for association with the phenotype conditional upon the prediction from the regression model in Step 1, using a leave one chromosome out (LOCO) scheme [22]. These analyses were restricted to SNPs with imputation score >0.5 and MAF>0.01 and adjusted for age at MRI, sex, intracranial volume and the first 10 PCs of population stratification. We finally annotated the nearest genes and SNPs functions using Annovar [23].

#### Supplementary References

1. Koch A, Stirnberg R, Estrada S, Zeng W, Lohner V, Shahid M, et al. Versatile MRI acquisition and processing protocol for population-based neuroimaging. Nat Protoc. 2024. 13 December 2024. https://doi.org/10.1038/s41596-024-01085-w.
2. Brenner D, Stirnberg R, Pracht ED, Stöcker T. Two-dimensional accelerated MP-RAGE imaging with flexible linear reordering. MAGMA. 2014;27:455–462.
3. van der Kouwe AJW, Benner T, Salat DH, Fischl B. Brain morphometry with multiecho MPRAGE. Neuroimage. 2008;40:559–569.
4. Tobisch A, Stirnberg R, Harms RL, Schultz T, Roebroeck A, Breteler MMB, et al. Compressed Sensing Diffusion Spectrum Imaging for Accelerated Diffusion Microstructure MRI in Long-Term Population Imaging. Front Neurosci. 2018;12:650.
5. Menzel MI, Tan ET, Khare K, Sperl JI, King KF, Tao X, et al. Accelerated diffusion spectrum imaging in the human brain using compressed sensing. Magn Reson Med. 2011;66:1226–1233.
6. Wedeen VJ, Hagmann P, Tseng W-YI, Reese TG, Weisskoff RM. Mapping complex tissue architecture with diffusion spectrum magnetic resonance imaging. Magn Reson Med. 2005;54:1377–1386.
7. Fischl B, Salat DH, Busa E, Albert M, Dieterich M, Haselgrove C, et al. Whole brain segmentation: automated labeling of neuroanatomical structures in the human brain. Neuron. 2002;33:341–355.
8. Fischl B. FreeSurfer. Neuroimage. 2012;62:774–781.
9. Tobisch A, Schultz T, Stirnberg R, Varela-Mattatall G, Knutsson H, Irarrázaval P, et al. Comparison of basis functions and q-space sampling schemes for robust compressed sensing reconstruction accelerating diffusion spectrum imaging. NMR Biomed. 2019;32:e4055.
10. Andersson JLR, Sotiropoulos SN. An integrated approach to correction for off-resonance effects and subject movement in diffusion MR imaging. Neuroimage. 2016;125:1063–1078.
11. Basser PJ, Mattiello J, LeBihan D. MR diffusion tensor spectroscopy and imaging. Biophys J. 1994;66:259–267.
12. Jensen JH, Helpern JA, Ramani A, Lu H, Kaczynski K. Diffusional kurtosis imaging: the quantification of non-gaussian water diffusion by means of magnetic resonance imaging. Magn Reson Med. 2005;53:1432–1440.
13. Harms RL, Fritz FJ, Tobisch A, Goebel R, Roebroeck A. Robust and fast nonlinear optimization of diffusion MRI microstructure models. Neuroimage. 2017;155:82–96.

14. Smith SM, Jenkinson M, Johansen-Berg H, Rueckert D, Nichols TE, Mackay CE, et al. Tract-based spatial statistics: voxelwise analysis of multi-subject diffusion data. Neuroimage. 2006;31:1487–1505.
15. Coors A, Breteler MMB, Ettinger U. Processing speed, but not working memory or global cognition, is associated with pupil diameter during fixation. Psychophysiology. 2022;59:e14089.
16. Mueller ST, Piper BJ. The Psychology Experiment Building Language (PEBL) and PEBL Test Battery. J Neurosci Methods. 2014;222:250–259.
17. Boenniger MM, Staerk C, Coors A, Huijbers W, Ettinger U, Breteler MMB. Ten German versions of Rey's auditory verbal learning test: Age and sex effects in 4,000 adults of the Rhineland Study. J Clin Exp Neuropsychol. 2021;43:637–653.
18. Coors A, Merten N, Ward DD, Schmid M, Breteler MMB, Ettinger U. Strong age but weak sex effects in eye movement performance in the general adult population: Evidence from the Rhineland Study. Vision Res. 2021;178:124–133.
19. Price AL, Patterson NJ, Plenge RM, Weinblatt ME, Shadick NA, Reich D. Principal components analysis corrects for stratification in genome-wide association studies. Nat Genet. 2006;38:904–909.
20. 1000 Genomes Project Consortium, Auton A, Brooks LD, Durbin RM, Garrison EP, Kang HM, et al. A global reference for human genetic variation. Nature. 2015;526:68–74.
21. Howie BN, Donnelly P, Marchini J. A flexible and accurate genotype imputation method for the next generation of genome-wide association studies. PLoS Genet. 2009;5:e1000529.
22. Mbatchou J, Barnard L, Backman J, Marcketta A, Kosmicki JA, Ziyatdinov A, et al. Computationally efficient whole-genome regression for quantitative and binary traits. Nat Genet. 2021;53:1097–1103.
23. Wang K, Li M, Hakonarson H. ANNOVAR: functional annotation of genetic variants from high-throughput sequencing data. Nucleic Acids Research. 2010;38:e164–e164.
24. Fu J, Wang J, Xue H, Wang M, Zhang B, Zhu W, et al. Genome-wide association studies of brain diffusion kurtosis imaging phenotypes. EBioMedicine. 2026;127:106261.

#### Supplementary Figures

Figure S1: Manhattan plots of DKI markers

**Axial kurtosis**

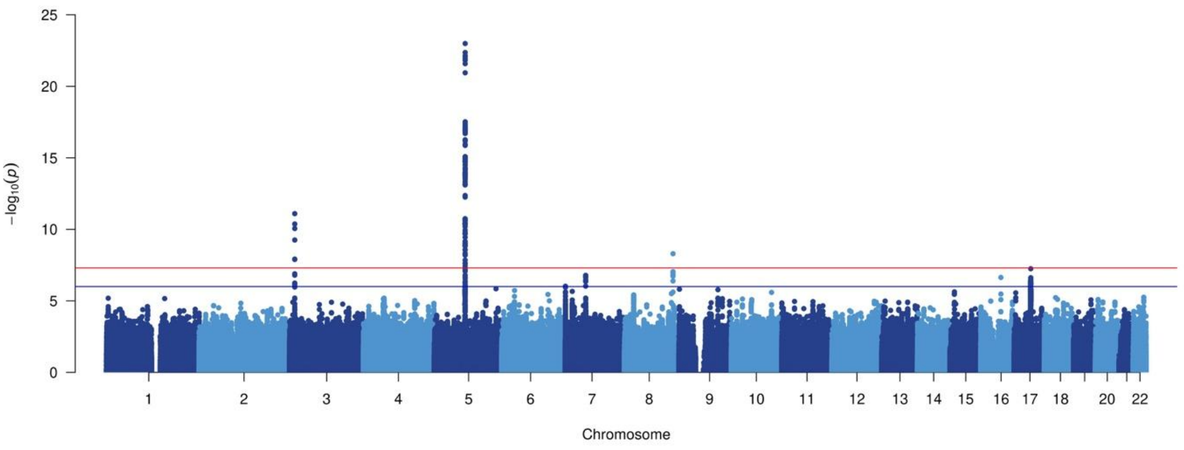

**Mean kurtosis**

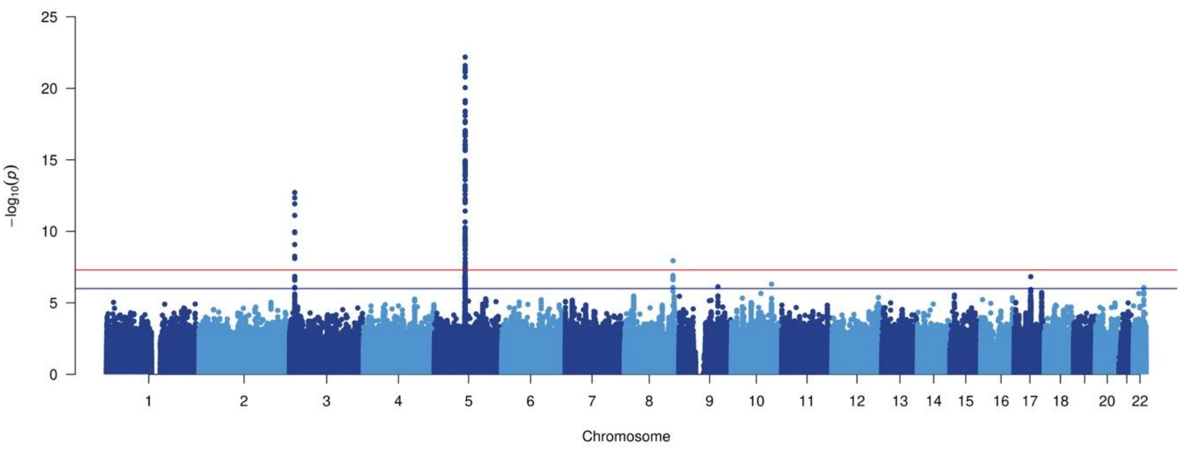

**Radial kurtosis**

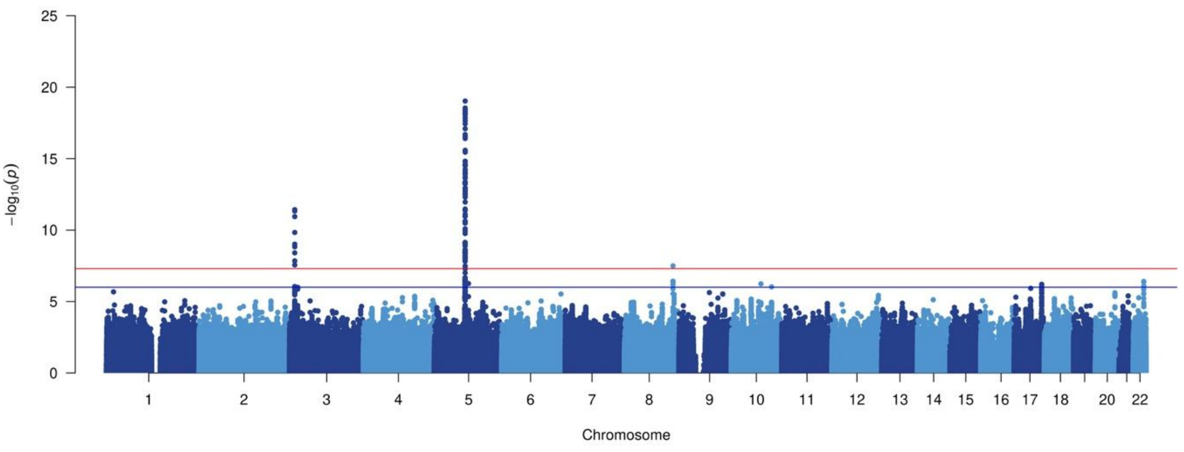

Figure S2: Regional plots of genome-wide significant DKI loci

**Axial kurtosis**

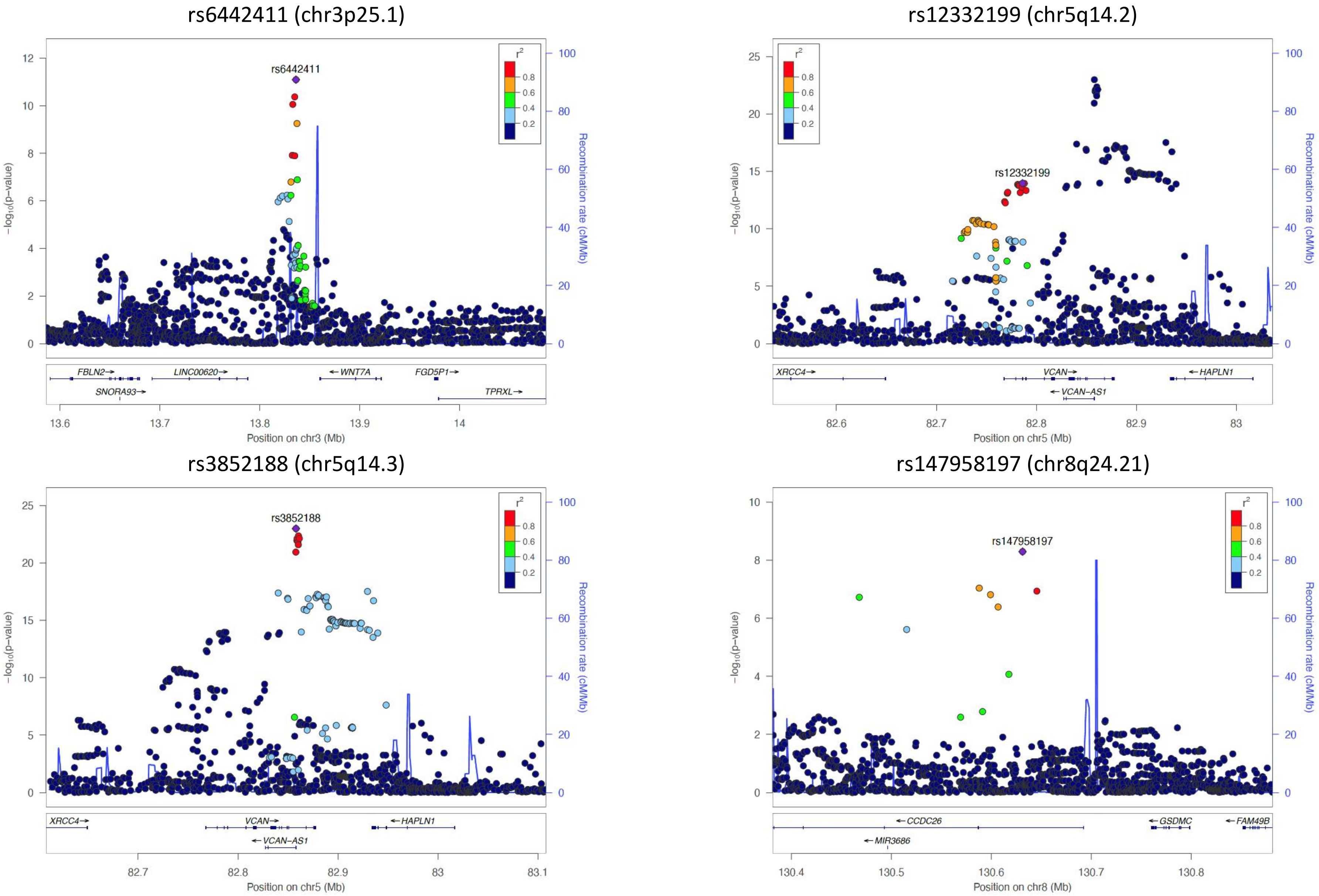

#### Mean kurtosis

rs6442411 (chr3p25.1)

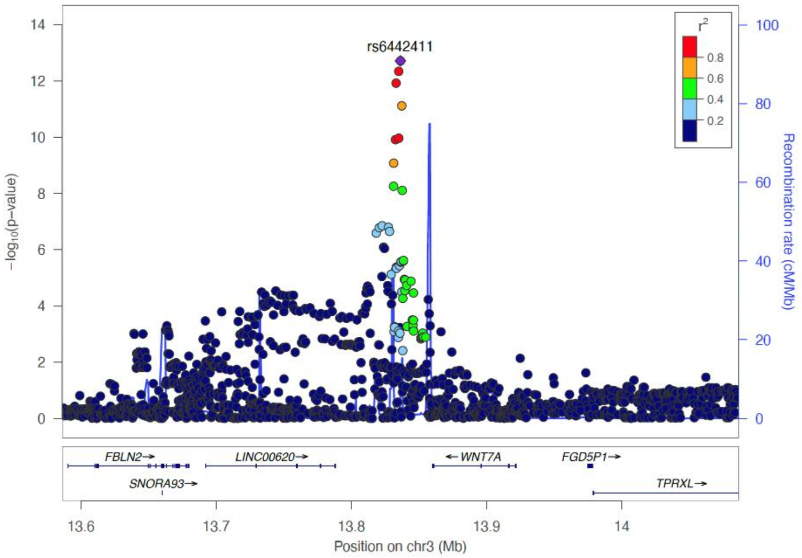

rs11749904 (chr5q14.2)

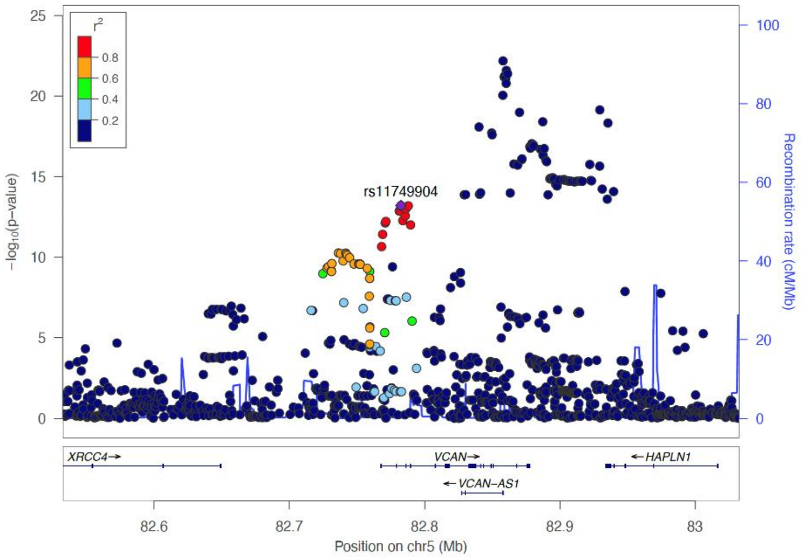

rs3852188 (chr5q14.3)

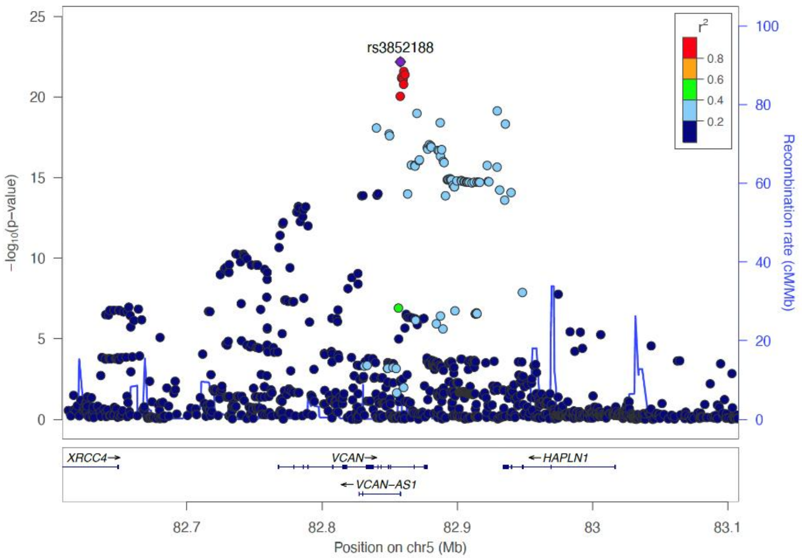

rs147958197 (chr8q24.21)

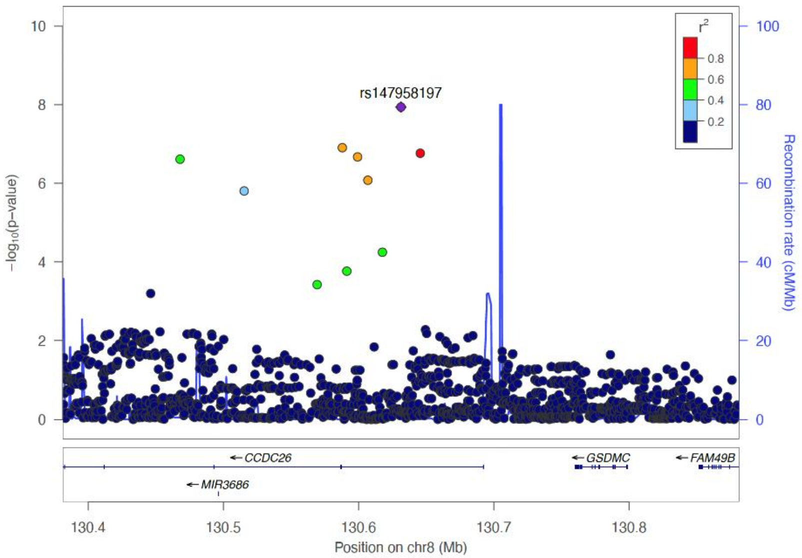

#### Radial kurtosis

##### rs6442411 (chr3p25.1)

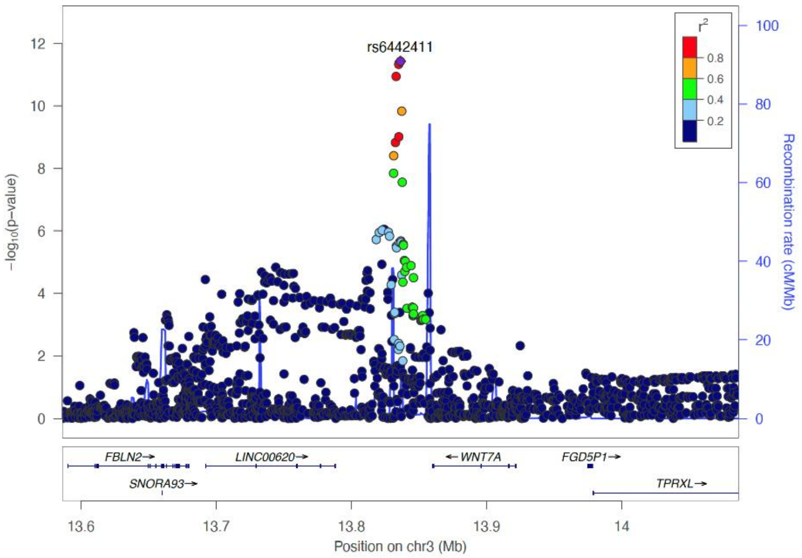

##### rs11749904 (chr5q14.2)

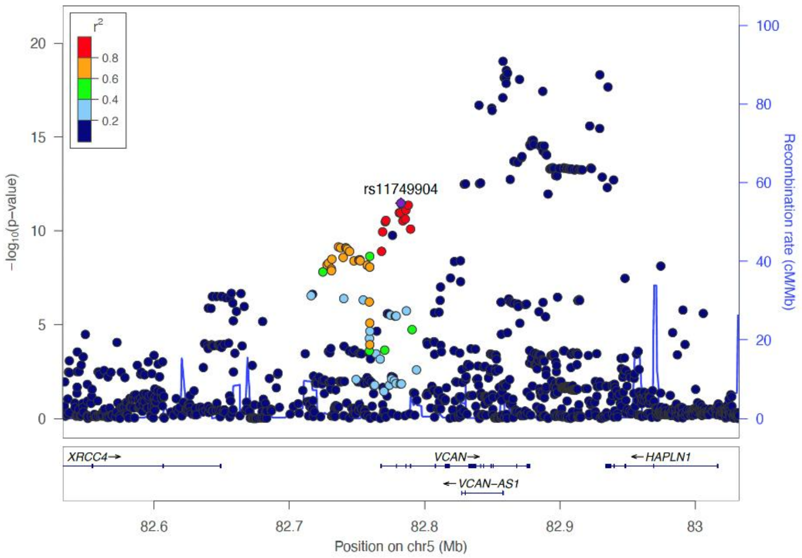

##### rs3852188 (chr5q14.3)

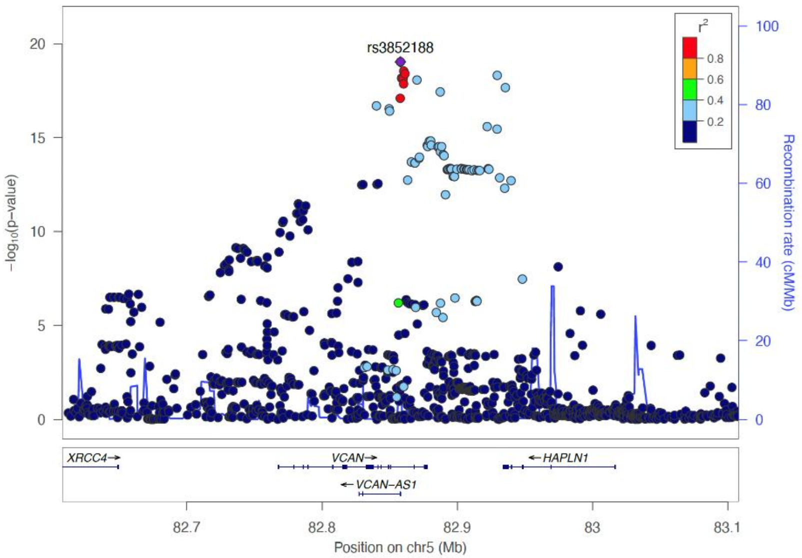

##### rs147958197 (chr8q24.21)

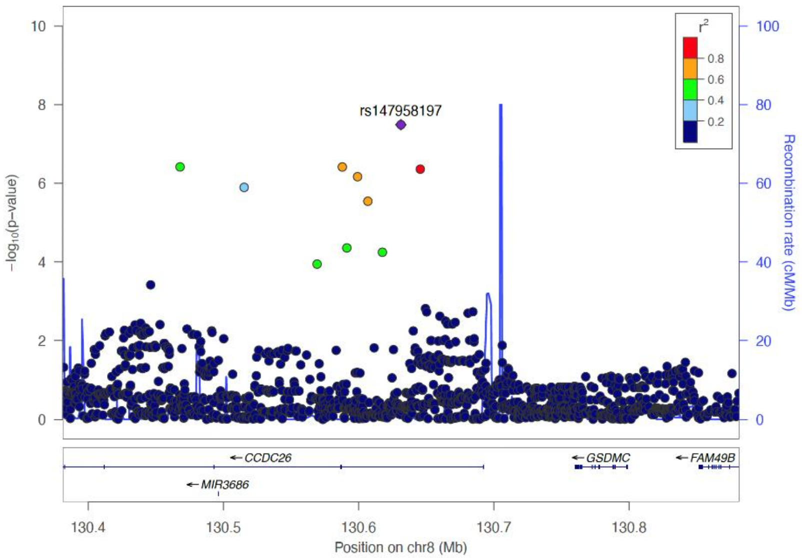

Figure S3: Sex-stratified results for genome-wide significant DKI loci

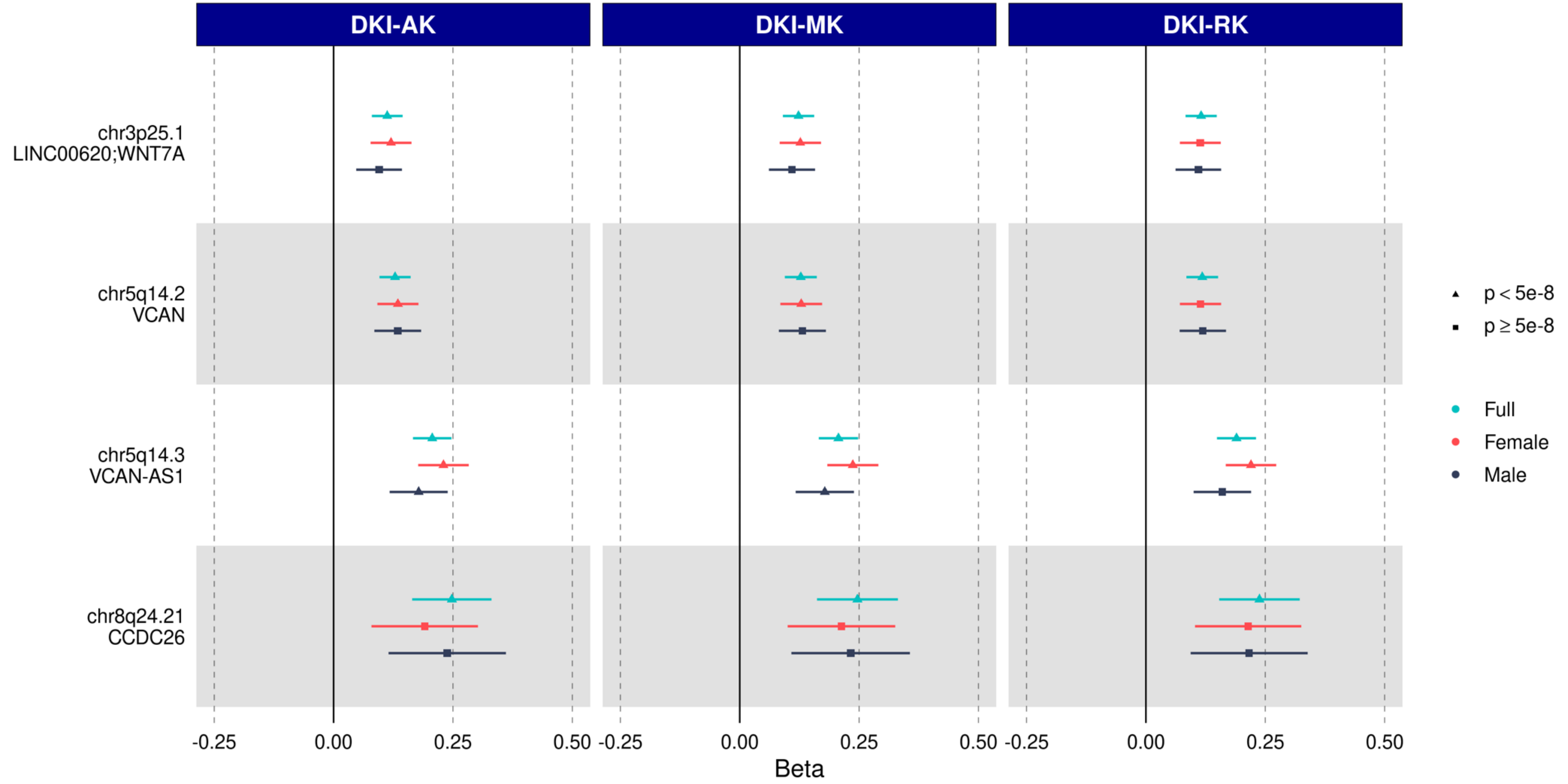

“Full”: Full sample (N=5 930); “Female”: Women only (N=3 427); “Male”: Men only (N=2 503). DKI: Diffusion kurtosis imaging; AK: Axial kurtosis; MK: Mean kurtosis; RK: Radial kurtosis.

Figure S4: Association of DKI loci with DTI, cSVD, stroke, dementia and vascular risk factors

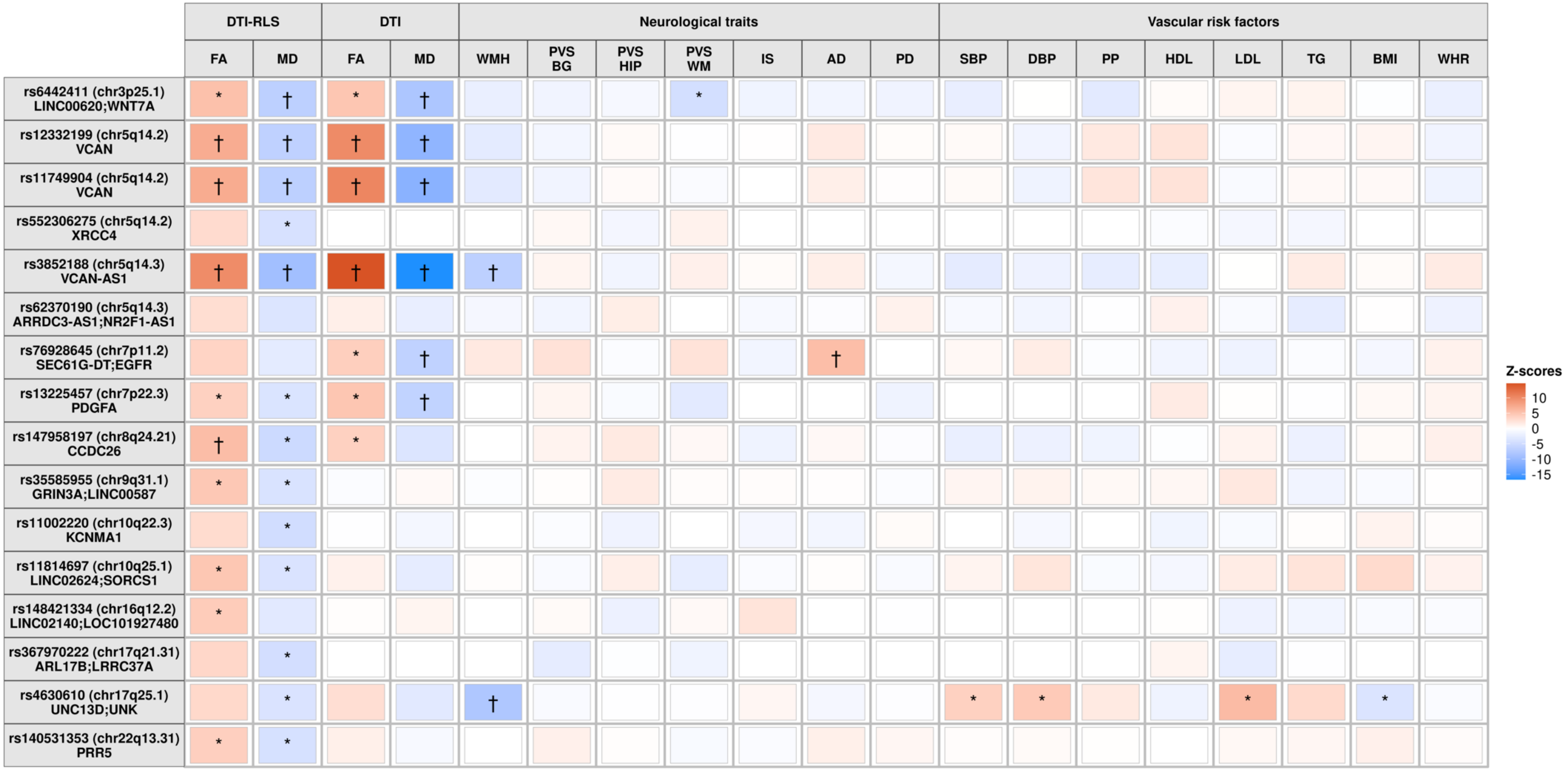

Only results with $p<0.05$ are colored. *: $p<1.75x10^{-4}$; †: $p<5x10^{-8}$. Z-score: Beta of effect allele (increase of DKI) / Standard error.
RLS: Rhineland Study; DTI: Diffusion tensor imaging; FA: Fractional anisotropy; MD: Mean diffusivity; WMH: White Matter Hyperintensities; PVS: Perivascular spaces; BG: Basal ganglia; HIP: Hippocampus; WM: White matter; IS: Ischemic stroke; AD: Alzheimer disease; PD: Parkinson disease; SBP: Systolic blood pressure; DBP: Diastolic blood pressure; PP: Pulse pressure; HDL: HDL-cholesterol; LDL: LDL-cholesterol; TG: Triglycerides; BMI: Body Mass Index; WHR: Waist-Hip Ratio.

Figure S5: Association of DKI loci with baseline and longitudinal cognitive scores in the Rhineland Study

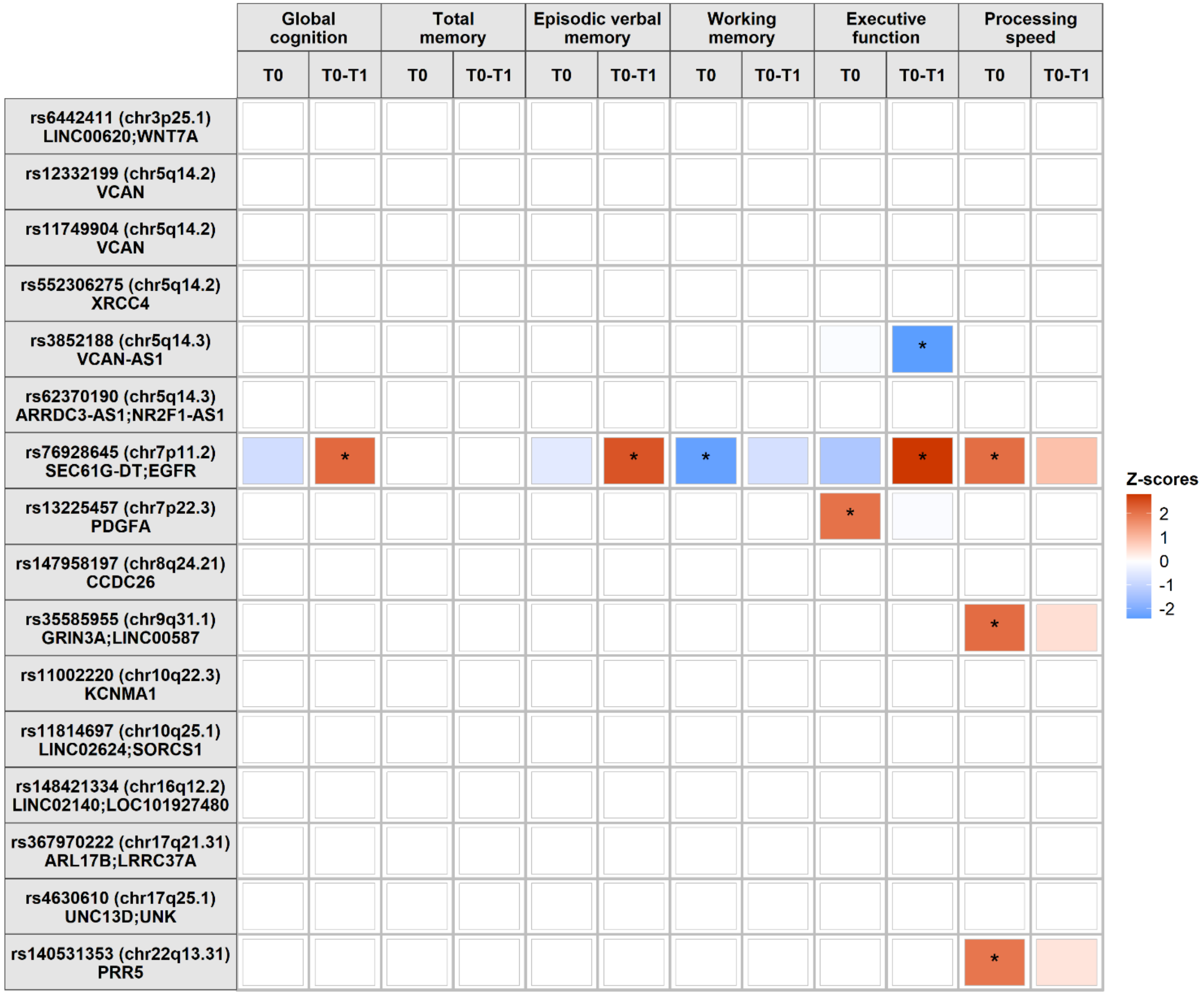

Only results with p<0.05 with T0 and/or T0-T1 are colored. *: p<0.05. Z-score: Beta of effect allele (increase of DKI) / Standard error.
T0: Baseline cognition; T0-T1: Cognitive trajectory between baseline and follow-up.

Figure S6: Transcriptome-wide association study (TWAS) of DKI markers in multiple tissues

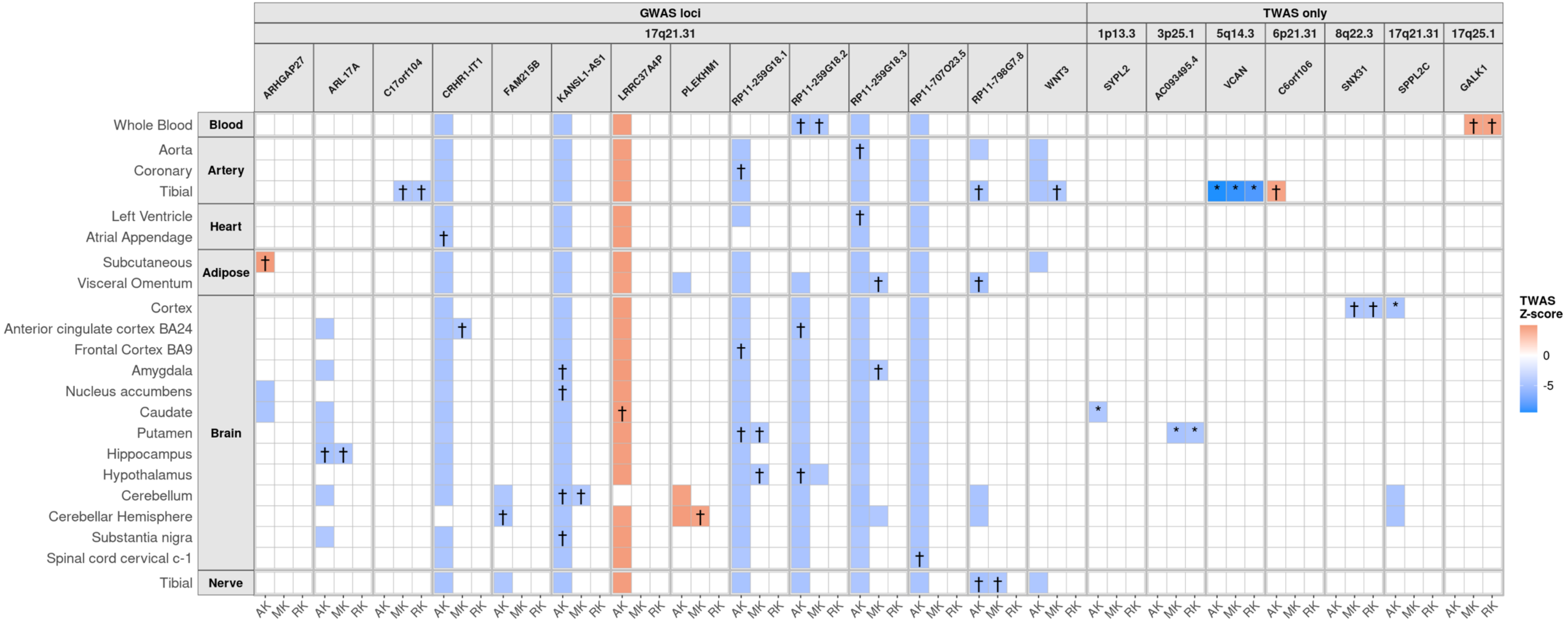

Only genes with significant TWAS ($p<8.9\times10^{-6}$) and conditional ($p<0.05$) association with at least one DKI marker are shown. Only results with TWAS $p<8.9\times10^{-6}$ are colored; *: TWAS $p<8.9\times10^{-6}$, $p<0.05$ in conditional analyses; †: TWAS $p<8.9\times10^{-6}$, $p<0.05$ in conditional analyses and COLOC-PP4≥0.75.
AK: Axial kurtosis; MK: Mean kurtosis; RK: Radial kurtosis.

Figure S7: Association of DKI loci with protein levels in UK Biobank

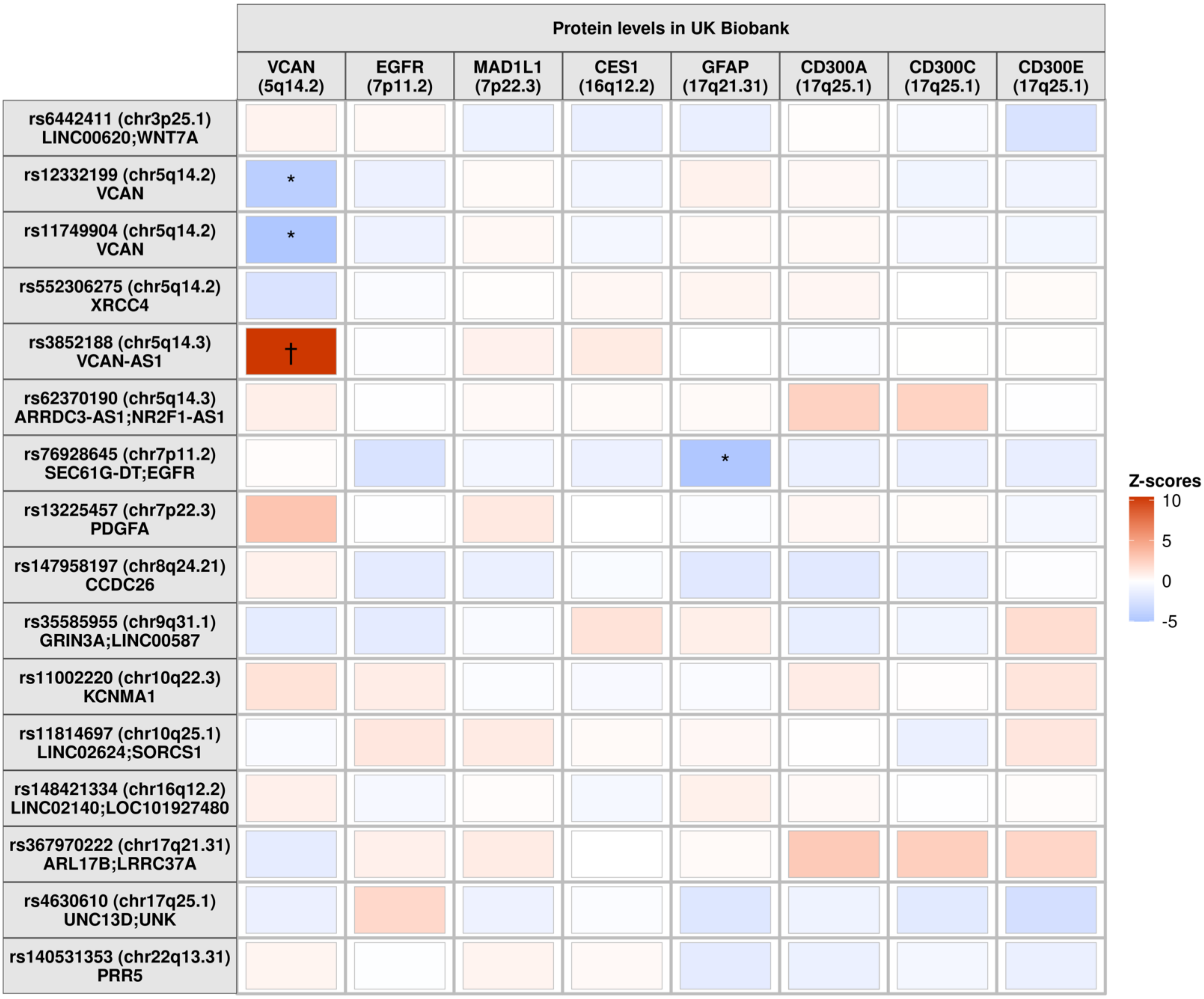

*: p p<4.17x10$^{-4}$; †: p<5x10$^{-8}$. Z-score: Beta of effect allele (increase of DKI) / Standard error.

#### Supplementary Tables

Table S1: GWAS results - DKI suggestive results at $p<1.10^{-6}$

| SNP | Locus | POS | Function | Nearest genes | EA | NEA | EAF | Beta | SE | p | p SNPxAge |
|---|---|---|---|---|---|---|---|---|---|---|---|
| **Axial kurtosis** | | | | | | | | | | | |
| rs6442411 | 3p25.1 | 13836296 | intergenic | LINC00620;WNT7A | T | C | 0.39 | 0.11 | 0.02 | **8.01E-12** | 0.0720 |
| rs12332199 | 5q14.2 | 82786194 | exonic | VCAN | T | C | 0.63 | 0.13 | 0.02 | **1.08E-14** | 0.6788 |
| rs3852188 | 5q14.3 | 82858014 | ncRNA_exonic | VCAN-AS1 | C | G | 0.80 | 0.21 | 0.02 | **1.02E-23** | **0.0257** |
| rs13225457 | 7p22.3 | 541830 | intronic | PDGFA | A | G | 0.36 | 0.08 | 0.02 | 9.50E-07 | 0.8252 |
| rs76928645 | 7p11.2 | 54941328 | intergenic | SEC61G-DT;EGFR | C | T | 0.90 | 0.14 | 0.03 | 1.66E-07 | 0.9113 |
| rs147958197 | 8q24.21 | 130631395 | ncRNA_intronic | CCDC26 | T | C | 0.96 | 0.25 | 0.04 | **5.05E-09** | 0.6545 |
| rs148421334 | 16q12.2 | 54485980 | intergenic | LINC02140;LOC101927480 | T | C | 0.02 | 0.36 | 0.07 | 2.31E-07 | 0.4248 |
| rs367970222 | 17q21.31 | 44388697 | intronic | ARL17B;LRRC37A | G | T | 0.80 | 0.13 | 0.02 | 5.71E-08 | 0.8588 |
| **Mean kurtosis** | | | | | | | | | | | |
| rs6442411 | 3p25.1 | 13836296 | intergenic | LINC00620;WNT7A | T | C | 0.39 | 0.12 | 0.02 | **1.94E-13** | 0.0960 |
| rs552306275 | 5q14.2 | 82613400 | intronic | XRCC4 | C | T | 0.89 | 0.15 | 0.03 | 3.91E-07 | 0.6426 |
| rs11749904 | 5q14.2 | 82782530 | intronic | VCAN | T | C | 0.64 | 0.13 | 0.02 | **6.21E-14** | 0.3711 |
| rs3852188 | 5q14.3 | 82858014 | ncRNA_exonic | VCAN-AS1 | C | G | 0.80 | 0.21 | 0.02 | **6.57E-23** | **0.0244** |
| rs147958197 | 8q24.21 | 130631395 | ncRNA_intronic | CCDC26 | T | C | 0.96 | 0.25 | 0.04 | **1.14E-08** | 0.7485 |
| rs35585955 | 9q31.1 | 104996411 | intergenic | GRIN3A;LINC00587 | T | C | 0.31 | 0.09 | 0.02 | 7.47E-07 | 0.2355 |
| rs11814697 | 10q25.1 | 108027940 | intergenic | LINC02624;SORCS1 | T | C | 0.06 | 0.17 | 0.03 | 4.95E-07 | 0.9204 |
| rs367970222 | 17q21.31 | 44388697 | intronic | ARL17B;LRRC37A | G | T | 0.80 | 0.13 | 0.02 | 1.48E-07 | 0.7062 |
| rs140531353 | 22q13.31 | 45088305 | intronic | PRR5 | A | C | 0.02 | 0.30 | 0.06 | 8.51E-07 | 0.8916 |
| **Radial kurtosis** | | | | | | | | | | | |
| rs6442411 | 3p25.1 | 13836296 | intergenic | LINC00620;WNT7A | T | C | 0.39 | 0.12 | 0.02 | **3.74E-12** | 0.0883 |
| rs552306275 | 5q14.2 | 82613400 | intronic | XRCC4 | C | T | 0.89 | 0.15 | 0.03 | 5.67E-07 | 0.5629 |
| rs11749904 | 5q14.2 | 82782530 | intronic | VCAN | T | C | 0.64 | 0.12 | 0.02 | **3.36E-12** | 0.1195 |
| rs3852188 | 5q14.3 | 82858014 | ncRNA_exonic | VCAN-AS1 | C | G | 0.80 | 0.19 | 0.02 | **9.33E-20** | **0.0186** |
| rs62370190 | 5q14.3 | 91746254 | intergenic | ARRDC3-AS1;NR2F1-AS1 | A | G | 0.92 | 0.15 | 0.03 | 5.54E-07 | 0.5589 |
| rs147958197 | 8q24.21 | 130631395 | ncRNA_intronic | CCDC26 | T | C | 0.96 | 0.24 | 0.04 | **3.21E-08** | 0.9280 |
| rs11002220 | 10q22.3 | 79376302 | intronic | KCNMA1 | A | G | 0.04 | 0.24 | 0.05 | 5.80E-07 | 0.8615 |
| rs11814697 | 10q25.1 | 108027940 | intergenic | LINC02624;SORCS1 | T | C | 0.06 | 0.16 | 0.03 | 9.42E-07 | 0.8183 |
| rs4630610 | 17q25.1 | 73822583 | downstream | UNC13D;UNK | C | A | 0.86 | 0.12 | 0.02 | 6.22E-07 | 0.2780 |
| rs140531353 | 22q13.31 | 45088305 | intronic | PRR5 | A | C | 0.02 | 0.31 | 0.06 | 3.91E-07 | 0.6985 |

POS: Position (build 37); EA: Effect allele; NEA: Non-effect allele; EAF: Effect allele frequency; Beta: Regression coefficient for EA; SE: Standard error for Beta; p: p-value. p SNPxAge: pvalue of SNPxAge interaction.
In bold: p-values significant at $p<5.10^{-8}$. p SNPxAge in bold: $p<0.05$.

Table S2: GWAS results - DKI loci with significant interaction with age

| SNP | Locus | POS | Function | Nearest genes | TYPE | Age | N | EA | NEA | Beta | SE | p | p SNPxAge |
|---|---|---|---|---|---|---|---|---|---|---|---|---|---|
| rs3852188 | 5q14.3 | 82858014 | ncRNA_exonic | *VCAN-AS1* | **Axial kurtosis** | Full Sample | 5930 | C | G | 0.21 | 0.02 | **1.02E-23** | **0.0257** |
| | | | | | | T1_30_50 | 1995 | | | 0.24 | 0.04 | **2.17E-10** | |
| | | | | | | T2_50_62 | 2008 | | | 0.23 | 0.04 | **2.33E-10** | |
| | | | | | | T3_62plus | 1927 | | | 0.14 | 0.03 | 2.46E-04 | |
| | | | | | **Mean kurtosis** | Full Sample | 5930 | C | G | 0.21 | 0.02 | **6.57E-23** | **0.0244** |
| | | | | | | T1_30_50 | 1995 | | | 0.24 | 0.04 | **4.30E-10** | |
| | | | | | | T2_50_62 | 2008 | | | 0.25 | 0.04 | **3.46E-11** | |
| | | | | | | T3_62plus | 1927 | | | 0.13 | 0.03 | 2.46E-04 | |
| | | | | | **Radial kurtosis** | Full Sample | 5930 | C | G | 0.19 | 0.02 | **9.33E-20** | **0.0186** |
| | | | | | | T1_30_50 | 1995 | | | 0.22 | 0.04 | **2.02E-09** | |
| | | | | | | T2_50_62 | 2008 | | | 0.23 | 0.04 | **3.28E-10** | |
| | | | | | | T3_62plus | 1927 | | | 0.12 | 0.03 | 2.46E-04 | |

POS: Position (build 37); EA: Effect allele; NEA: Non-effect allele; Beta: Regression coefficient for EA; SE: Standard error for Beta; p: p-value. p SNPxAge: pvalue of SNPxAge interaction. T1_30_50: Age tertile 1 (30-50 years); T2_50_62: Age tertile 2 (50-62 years); T3_62plus: Age tertile 3 (>63 years).
In bold: p-values significant at $p<5.10^{-8}$. p SNPxAge in bold: $p<0.05$.

Table S3: Associations of DKI loci identified in a Chinese population with DKI markers in the Rhineland Study

| | | | | CHIMGEN* | | | Rhineland study | |
|---|---|---|---|---|---|---|---|---|
| SNP | Locus | Position | Nearest genes | EA | Beta | p | Beta | p |
| **Axial kurtosis** | | | | | | | | |
| rs185356 | 5q14.2 | 82729928 | XRCC4;VCAN | C | 0.12 | 2.97E-08 | 0.08 | **3.47E-06** |
| rs16901197 | 5q14.3 | 83976407 | EDIL3;NBPF22P | G | 0.16 | 2.71E-08 | -0.06 | 3.62E-01 |
| rs1363164 | 5q23.2 | 121598606 | LOC100505841;SNCAIP | A | 0.12 | 3.17E-08 | 0.00 | 9.98E-01 |
| rs11247104 | 15q26.3 | 100046049 | LRRC28;MEF2A | A | 0.14 | 5.65E-09 | 0.01 | 6.13E-01 |
| rs2076404 | 20p13 | 2380396 | TGM6 | A | 0.13 | 3.02E-08 | 0.02 | 2.36E-01 |
| **Mean kurtosis** | | | | | | | | |
| rs34908550 | 1p31.3 | 68740499 | WLS;RPE65 | A | 0.21 | 3.77E-08 | -0.01 | 5.72E-01 |
| rs256460 | 5q14.2 | 82759588 | XRCC4;VCAN | C | 0.13 | 1.11E-09 | 0.08 | **2.12E-06** |
| rs28296 | 5q14.2 | 82760017 | XRCC4;VCAN | T | 0.16 | 1.31E-13 | 0.07 | **6.45E-05** |
| rs4558964 | 5q14.2 | 82786549 | VCAN | G | 0.15 | 5.77E-12 | 0.10 | **3.03E-08** |
| rs4470745 | 5q14.2 | 82789647 | VCAN | A | 0.23 | 3.65E-08 | 0.12 | **9.91E-13** |
| rs149529 | 5q14.3 | 82829385 | VCAN-AS1 | T | 0.16 | 3.80E-10 | 0.20 | **1.34E-14** |
| rs10052710 | 5q14.3 | 82860025 | VCAN | T | 0.16 | 6.37E-11 | 0.20 | **7.61E-22** |
| rs13176921 | 5q14.3 | 82860348 | VCAN | A | 0.18 | 3.32E-14 | 0.20 | **1.63E-21** |
| rs67827860 | 5q14.3 | 82860485 | VCAN | C | 0.14 | 2.29E-08 | 0.20 | **2.58E-22** |
| rs12206116 | 6q24.2 | 143949714 | PHACTR2 | A | 0.66 | 8.69E-09 | -0.03 | 5.29E-02 |
| rs17104965 | 14q13.3 | 37145916 | PAX9 | C | 0.15 | 2.00E-08 | 0.00 | 9.69E-01 |
| rs117668447 | 18q21.31 | 56122696 | MIR122;ALPK2 | G | 0.29 | 8.64E-09 | 0.02 | 8.23E-01 |
| **Radial kurtosis** | | | | | | | | |
| rs115854471 | 3q12.3 | 101636861 | NFKBIZ;LINC02085 | A | 0.24 | 2.61E-08 | 0.00 | 9.84E-01 |
| rs2346012 | 5q31.1 | 135314880 | LECT2;TGFBI | G | 0.14 | 7.98E-10 | 0.00 | 8.33E-01 |

EA: Effect allele (DKI-increasing in CHIMGEN); Beta: Regression coefficient for EA; p: p-value. In bold: p-values significant at $1.09\times10^{-3}$. In blue: p-values significant at $p<5\times10^{-8}$. * From [24].

Table S4: Associations of DKI loci with DTI, cSVD, stroke, dementia and vascular risk factors

| Trait | SNP | Locus | CHR | Position | Gene | EA | NEA | Beta | SE | p |
|---|---|---|---|---|---|---|---|---|---|---|
| DTI-MD | rs6442411 | 3p25.1 | 3 | 13836296 | LINC00620;WNT7A | T | C | -0.06 | 0.01 | **1.20E-16** |
| RLS-DTI-MD | rs6442411 | 3p25.1 | 3 | 13836296 | LINC00620;WNT7A | T | C | -0.10 | 0.02 | **1.03E-11** |
| RLS-DTI-FA | rs6442411 | 3p25.1 | 3 | 13836296 | LINC00620;WNT7A | T | C | 0.08 | 0.02 | 6.60E-08 |
| PVS-WM | rs6442411 | 3p25.1 | 3 | 13836296 | LINC00620;WNT7A | T | C | -0.02 | 0.00 | 6.95E-08 |
| DTI-FA | rs6442411 | 3p25.1 | 3 | 13836296 | LINC00620;WNT7A | T | C | 0.04 | 0.01 | 6.93E-07 |
| RLS-DTI-MD | rs552306275 | 5q14.2 | 5 | 82613400 | XRCC4 | C | T | -0.11 | 0.03 | 2.66E-05 |
| DTI-MD | rs11749904 | 5q14.2 | 5 | 82782530 | VCAN | T | C | -0.08 | 0.01 | **2.74E-29** |
| DTI-FA | rs11749904 | 5q14.2 | 5 | 82782530 | VCAN | T | C | 0.08 | 0.01 | **8.63E-26** |
| RLS-DTI-FA | rs11749904 | 5q14.2 | 5 | 82782530 | VCAN | T | C | 0.11 | 0.02 | **4.34E-13** |
| RLS-DTI-MD | rs11749904 | 5q14.2 | 5 | 82782530 | VCAN | T | C | -0.11 | 0.02 | **1.55E-11** |
| DTI-MD | rs12332199 | 5q14.2 | 5 | 82786194 | VCAN | T | C | -0.08 | 0.01 | **3.34E-27** |
| DTI-FA | rs12332199 | 5q14.2 | 5 | 82786194 | VCAN | T | C | 0.08 | 0.01 | **7.24E-24** |
| RLS-DTI-FA | rs12332199 | 5q14.2 | 5 | 82786194 | VCAN | T | C | 0.11 | 0.02 | **6.85E-13** |
| RLS-DTI-MD | rs12332199 | 5q14.2 | 5 | 82786194 | VCAN | T | C | -0.10 | 0.02 | **3.77E-11** |
| DTI-MD | rs3852188 | 5q14.3 | 5 | 82858014 | VCAN-AS1 | C | G | -0.15 | 0.01 | **1.32E-62** |
| DTI-FA | rs3852188 | 5q14.3 | 5 | 82858014 | VCAN-AS1 | C | G | 0.14 | 0.01 | **4.34E-49** |
| RLS-DTI-FA | rs3852188 | 5q14.3 | 5 | 82858014 | VCAN-AS1 | C | G | 0.19 | 0.02 | **1.58E-23** |
| RLS-DTI-MD | rs3852188 | 5q14.3 | 5 | 82858014 | VCAN-AS1 | C | G | -0.18 | 0.02 | **2.30E-20** |
| WMH | rs3852188 | 5q14.3 | 5 | 82858014 | VCAN-AS1 | C | G | -0.05 | 0.01 | **1.13E-11** |
| DTI-MD | rs13225457 | 7p22.3 | 7 | 541830 | PDGFA | A | G | -0.05 | 0.01 | **1.79E-10** |
| DTI-FA | rs13225457 | 7p22.3 | 7 | 541830 | PDGFA | A | G | 0.04 | 0.01 | 7.74E-07 |
| RLS-DTI-FA | rs13225457 | 7p22.3 | 7 | 541830 | PDGFA | A | G | 0.06 | 0.02 | 6.80E-05 |
| RLS-DTI-MD | rs13225457 | 7p22.3 | 7 | 541830 | PDGFA | A | G | -0.06 | 0.02 | 1.16E-04 |
| DTI-MD | rs76928645 | 7p11.2 | 7 | 54941328 | SEC61G-DT;EGFR | C | T | -0.08 | 0.01 | **7.95E-11** |
| AD | rs76928645 | 7p11.2 | 7 | 54941328 | SEC61G-DT;EGFR | C | T | 0.08 | 0.01 | **1.51E-08** |
| DTI-FA | rs76928645 | 7p11.2 | 7 | 54941328 | SEC61G-DT;EGFR | C | T | 0.05 | 0.01 | 3.39E-05 |
| RLS-DTI-FA | rs147958197 | 8q24.21 | 8 | 130631395 | CCDC26 | T | C | 0.22 | 0.04 | **1.07E-08** |
| RLS-DTI-MD | rs147958197 | 8q24.21 | 8 | 130631395 | CCDC26 | T | C | -0.21 | 0.04 | 1.47E-07 |
| DTI-FA | rs147958197 | 8q24.21 | 8 | 130631395 | CCDC26 | T | C | 0.08 | 0.02 | 7.96E-05 |
| RLS-DTI-FA | rs35585955 | 9q31.1 | 9 | 104996411 | GRIN3A;LINC00587 | T | C | 0.08 | 0.02 | 1.75E-06 |
| RLS-DTI-MD | rs35585955 | 9q31.1 | 9 | 104996411 | GRIN3A;LINC00587 | T | C | -0.06 | 0.02 | 6.65E-05 |
| RLS-DTI-MD | rs11002220 | 10q22.3 | 10 | 79376302 | KCNMA1 | A | G | -0.21 | 0.04 | 1.33E-06 |
| RLS-DTI-FA | rs11814697 | 10q25.1 | 10 | 108027940 | LINC02624;SORCS1 | T | C | 0.15 | 0.03 | 1.01E-06 |
| RLS-DTI-MD | rs11814697 | 10q25.1 | 10 | 108027940 | LINC02624;SORCS1 | T | C | -0.12 | 0.03 | 8.97E-05 |
| RLS-DTI-FA | rs148421334 | 16q12.2 | 16 | 54485980 | LINC02140;LOC101927480 | T | C | 0.29 | 0.06 | 9.30E-06 |
| RLS-DTI-MD | rs367970222 | 17q21.31 | 17 | 44388697 | ARL17B;LRRC37A | G | T | -0.10 | 0.02 | 3.60E-06 |
| WMH | rs4630610 | 17q25.1 | 17 | 73822583 | UNC13D;UNK | C | A | -0.06 | 0.01 | **8.35E-16** |
| LDL | rs4630610 | 17q25.1 | 17 | 73822583 | UNC13D;UNK | C | A | 0.01 | 0.00 | 4.37E-07 |
| DBP | rs4630610 | 17q25.1 | 17 | 73822583 | UNC13D;UNK | C | A | 0.11 | 0.02 | 5.11E-06 |
| SBP | rs4630610 | 17q25.1 | 17 | 73822583 | UNC13D;UNK | C | A | 0.17 | 0.04 | 7.40E-05 |
| RLS-DTI-MD | rs4630610 | 17q25.1 | 17 | 73822583 | UNC13D;UNK | C | A | -0.09 | 0.02 | 8.98E-05 |
| BMI | rs4630610 | 17q25.1 | 17 | 73822583 | UNC13D;UNK | C | A | -0.01 | 0.00 | 1.37E-04 |
| RLS-DTI-MD | rs140531353 | 22q13.31 | 22 | 45088305 | PRR5 | A | C | -0.24 | 0.06 | 2.03E-05 |
| RLS-DTI-FA | rs140531353 | 22q13.31 | 22 | 45088305 | PRR5 | A | C | 0.23 | 0.06 | 3.87E-05 |

Only results with $p<1.75 \times 10^{-4}$ are shown. Genome-wide significant results ($p<5 \times 10^{-8}$) are in bold.

EA: Effect allele; NEA: Non-effect allele; Beta: Effect size of effect allele (increase of DKI); SE: Standard error.

RLS: Rhineland Study; DTI: Diffusion tensor imaging; FA: Fractional anisotropy; MD: Mean diffusivity; WMH: White Matter Hyperintensities; PVS: Perivascular spaces; BG: Basal ganglia; HIP: Hippocampus; WM: White matter; IS: Ischemic stroke; AD: Alzheimer disease; PD: Parkinson disease; SBP: Systolic blood pressure; DBP: Diastolic blood pressure; PP: Pulse pressure; HDL: HDL-cholesterol; LDL: LDL-cholesterol; TG: Triglycerides; BMI: Body Mass Index; WHR: Waist-Hip Ratio.

Table S5: Association of loci associated with neurological traits with DKI and DTI markers

| SNP | Locus | Position | Nearest genes | Risk allele | Axial kurtosis | | Mean kurtosis | | Radial kurtosis | | Fractional anisotropy | | Mean diffusivity | |
|---|---|---|---|---|---|---|---|---|---|---|---|---|---|---|
| | | | | | Beta | p | Beta | p | Beta | p | Beta | p | Beta | p |
| **Alzheimer disease** | | | | | | | | | | | | | | |
| rs679515 | 1q32.2 | 207750568 | CR1 | T | 0.01 | 6.93E-01 | 0.00 | 8.55E-01 | 0.01 | 6.85E-01 | 0.00 | 8.92E-01 | 0.00 | 9.20E-01 |
| rs72777026 | 2p25.1 | 9699011 | ADAM17 | G | -0.02 | 4.71E-01 | -0.03 | 2.36E-01 | -0.03 | 2.03E-01 | 0.00 | 9.45E-01 | 0.01 | 5.72E-01 |
| rs17020490 | 2p22.2 | 37531939 | PRKD3 | C | 0.02 | 3.61E-01 | 0.03 | 2.36E-01 | 0.03 | 2.64E-01 | 0.01 | 6.73E-01 | -0.01 | 5.25E-01 |
| rs6733839 | 2q14.3 | 127892810 | BIN1 | T | -0.05 | **4.21E-03** | -0.04 | **7.25E-03** | -0.04 | **2.64E-02** | -0.05 | **3.89E-04** | 0.03 | **3.80E-02** |
| rs10933431 | 2q37.1 | 233981912 | INPP5D | C | 0.03 | 1.61E-01 | 0.03 | 1.80E-01 | 0.02 | 2.29E-01 | 0.02 | 3.58E-01 | -0.02 | 2.58E-01 |
| rs16824536 | 3q25.2 | 154787511 | MME | G | 0.02 | 5.03E-01 | 0.02 | 5.43E-01 | 0.03 | 4.29E-01 | 0.02 | 5.24E-01 | -0.05 | 1.37E-01 |
| rs61762319 | 3q25.2 | 154801978 | MME | G | 0.01 | 8.08E-01 | 0.00 | 9.82E-01 | 0.00 | 9.43E-01 | 0.03 | 6.22E-01 | -0.03 | 5.57E-01 |
| rs6846529 | 4p16.1 | 11025131 | CLNK | C | -0.03 | 1.53E-01 | -0.02 | 2.22E-01 | -0.02 | 3.10E-01 | -0.03 | 8.12E-02 | 0.01 | 3.86E-01 |
| rs2245466 | 4p14 | 40198846 | RHOH | G | 0.00 | 7.78E-01 | 0.00 | 9.90E-01 | 0.00 | 8.14E-01 | 0.00 | 9.00E-01 | 0.01 | 5.22E-01 |
| rs112403360 | 5p15.2 | 14724413 | ANKH | A | 0.00 | 9.00E-01 | 0.01 | 7.29E-01 | 0.01 | 6.80E-01 | 0.01 | 7.23E-01 | -0.01 | 7.88E-01 |
| rs62374257 | 5q14.3 | 86223195 | COX7C | C | -0.03 | 1.50E-01 | -0.02 | 3.14E-01 | -0.01 | 4.64E-01 | -0.01 | 6.68E-01 | 0.01 | 6.90E-01 |
| rs871269 | 5q33.1 | 150432388 | TNIP1 | C | 0.00 | 9.81E-01 | 0.01 | 6.95E-01 | 0.01 | 5.44E-01 | 0.00 | 9.01E-01 | -0.01 | 3.76E-01 |
| rs113706587 | 5q35.3 | 179628150 | RASGEF1C | A | -0.02 | 4.97E-01 | -0.01 | 6.46E-01 | -0.01 | 5.85E-01 | 0.00 | 9.72E-01 | 0.02 | 3.97E-01 |
| **rs6605556** | **6p21.32** | **32583099** | **HLA-DQA1** | A | -0.09 | **1.80E-04** | -0.10 | **6.61E-05** | -0.09 | **5.48E-04** | -0.06 | **5.44E-03** | 0.07 | **4.20E-03** |
| rs10947943 | 6p21.1 | 41004093 | UNC5CL | G | 0.01 | 5.53E-01 | 0.01 | 8.20E-01 | 0.00 | 8.63E-01 | 0.02 | 3.46E-01 | 0.00 | 9.45E-01 |
| rs143332484 | 6p21.1 | 41129207 | TREM2 | T | 0.04 | 6.40E-01 | 0.03 | 7.57E-01 | 0.04 | 6.41E-01 | 0.00 | 9.87E-01 | -0.02 | 8.45E-01 |
| rs7767350 | 6p12.3 | 47485126 | CD2AP | T | -0.04 | **3.45E-02** | -0.03 | 1.58E-01 | -0.01 | 6.21E-01 | -0.03 | 6.93E-02 | 0.01 | 6.19E-01 |
| rs785129 | 6q22.1 | 114612895 | HS3ST5 | T | -0.02 | 2.17E-01 | -0.02 | 2.71E-01 | -0.01 | 4.78E-01 | -0.03 | 7.49E-02 | 0.02 | 2.68E-01 |
| rs6943429 | 7p21.3 | 7856894 | UMAD1 | T | -0.01 | 4.79E-01 | -0.02 | 2.10E-01 | -0.02 | 2.27E-01 | 0.00 | 8.80E-01 | 0.01 | 6.69E-01 |
| rs10952097 | 7p21.3 | 8244012 | ICA1 | T | -0.02 | 4.61E-01 | -0.03 | 2.51E-01 | -0.04 | 2.09E-01 | -0.02 | 3.80E-01 | 0.03 | 2.51E-01 |
| rs13237518 | 7p21.3 | 12269593 | TMEM106B | C | -0.03 | 8.15E-02 | -0.03 | 8.60E-02 | -0.03 | 8.32E-02 | -0.02 | 3.09E-01 | 0.03 | 7.33E-02 |
| rs6966331 | 7p14.1 | 37883793 | EPDR1 | C | 0.00 | 8.68E-01 | 0.00 | 8.78E-01 | 0.00 | 9.26E-01 | 0.00 | 8.93E-01 | 0.01 | 7.13E-01 |
| **rs76928645** | **7p11.2** | **54941328** | **SEC61G** | C | 0.14 | **1.66E-07** | 0.12 | **2.09E-05** | 0.10 | **3.25E-04** | 0.09 | **2.29E-04** | -0.07 | **4.05E-03** |
| rs7384878 | 7q22.1 | 99932049 | SPDYE3 | T | 0.05 | **7.28E-03** | 0.04 | **1.24E-02** | 0.04 | **2.31E-02** | 0.04 | **9.77E-03** | -0.04 | **2.76E-02** |
| rs11771145 | 7q35 | 143110762 | EPHA1 | G | 0.02 | 2.41E-01 | 0.01 | 4.53E-01 | 0.01 | 7.26E-01 | 0.01 | 3.86E-01 | 0.01 | 5.21E-01 |
| rs1065712 | 8p23.1 | 11702122 | CTSB | C | 0.04 | 2.54E-01 | 0.05 | 1.35E-01 | 0.04 | 1.72E-01 | 0.02 | 5.84E-01 | 0.00 | 9.60E-01 |
| rs73223431 | 8p21.2 | 27219987 | PTK2B | T | -0.02 | 1.99E-01 | -0.01 | 4.86E-01 | -0.01 | 4.93E-01 | -0.01 | 3.66E-01 | 0.02 | 3.47E-01 |
| rs11787077 | 8p21.1 | 27465312 | CLU | C | 0.00 | 8.68E-01 | 0.00 | 8.24E-01 | 0.00 | 7.73E-01 | 0.00 | 7.61E-01 | -0.01 | 7.14E-01 |
| rs34173062 | 8q24.3 | 145158607 | SHARPIN | A | -0.06 | 5.28E-02 | -0.03 | 2.94E-01 | -0.01 | 7.55E-01 | -0.07 | **1.50E-02** | 0.05 | 1.24E-01 |
| rs1800978 | 9q31.1 | 107665978 | ABCA1 | G | 0.05 | 5.15E-02 | 0.04 | 1.01E-01 | 0.03 | 2.04E-01 | 0.04 | 6.93E-02 | -0.02 | 3.47E-01 |
| rs7912495 | 10p14 | 11718713 | USP6NL | G | -0.01 | 6.85E-01 | 0.00 | 7.73E-01 | 0.01 | 3.72E-01 | -0.01 | 5.03E-01 | -0.01 | 6.13E-01 |
| rs7068231 | 10q21.2 | 61784928 | ANK3 | G | 0.01 | 6.81E-01 | 0.01 | 7.46E-01 | 0.00 | 9.67E-01 | 0.01 | 7.28E-01 | 0.00 | 8.04E-01 |
| rs6586028 | 10q23.1 | 82253984 | TSPAN14 | T | -0.01 | 7.29E-01 | -0.01 | 6.59E-01 | 0.00 | 8.13E-01 | -0.01 | 6.55E-01 | -0.01 | 4.63E-01 |
| rs6584063 | 10q24.1 | 98026407 | BLNK | A | 0.00 | 9.91E-01 | 0.01 | 8.31E-01 | 0.00 | 9.06E-01 | 0.00 | 9.25E-01 | -0.02 | 6.15E-01 |
| rs7908662 | 10q26.13 | 124172912 | PLEKHA1 | A | 0.03 | 1.03E-01 | 0.03 | 6.33E-02 | 0.04 | **3.09E-02** | 0.03 | 6.09E-02 | -0.03 | **2.76E-02** |

| SNP | Locus | Position | Nearest genes | Risk allele | Axial kurtosis | | Mean kurtosis | | Radial kurtosis | | Fractional anisotropy | | Mean diffusivity | |
|---|---|---|---|---|---|---|---|---|---|---|---|---|---|---|
| | | | | | Beta | p | Beta | p | Beta | p | Beta | p | Beta | p |
| rs10437655 | 11p11.2 | 47391948 | SPI1 | A | -0.03 | 9.48E-02 | -0.04 | **2.87E-02** | -0.04 | **1.95E-02** | -0.02 | 2.93E-01 | 0.03 | 8.55E-02 |
| rs1582763 | 11q12.2 | 60021948 | MS4A4A | G | -0.01 | 4.33E-01 | -0.01 | 4.14E-01 | -0.02 | 2.92E-01 | -0.01 | 4.50E-01 | 0.02 | 1.91E-01 |
| rs3851179 | 11q14.2 | 85868640 | EED | C | 0.02 | 3.20E-01 | 0.02 | 2.10E-01 | 0.03 | 1.11E-01 | 0.02 | 1.15E-01 | -0.03 | **2.82E-02** |
| rs74685827 | 11q24.1 | 121353077 | SORL1 | G | 0.04 | 5.74E-01 | 0.09 | 1.82E-01 | 0.12 | 8.20E-02 | 0.05 | 4.04E-01 | -0.06 | 3.29E-01 |
| rs11218343 | 11q24.1 | 121435587 | SORL1 | T | 0.04 | 2.94E-01 | 0.04 | 2.93E-01 | 0.05 | 2.34E-01 | 0.07 | 6.64E-02 | -0.08 | **3.36E-02** |
| rs6489896 | 12q24.13 | 113719788 | TPCN1 | C | 0.03 | 4.50E-01 | 0.05 | 1.33E-01 | 0.06 | 9.09E-02 | 0.02 | 5.99E-01 | -0.05 | 1.64E-01 |
| rs17125924 | 14q22.1 | 53391680 | FERMT2 | G | 0.00 | 9.31E-01 | 0.01 | 7.87E-01 | 0.00 | 8.96E-01 | 0.01 | 8.16E-01 | -0.01 | 5.79E-01 |
| rs7401792 | 14q32.12 | 92931261 | SLC24A4 | G | -0.01 | 5.30E-01 | -0.01 | 5.41E-01 | -0.01 | 4.41E-01 | -0.01 | 5.27E-01 | 0.02 | 3.14E-01 |
| rs12590654 | 14q32.12 | 92938855 | SLC24A4 | G | 0.01 | 3.81E-01 | 0.01 | 6.39E-01 | 0.00 | 9.10E-01 | 0.00 | 8.67E-01 | 0.02 | 3.43E-01 |
| rs7157106 | 14q32.33 | 106228095 | IGHgenecluster | A | 0.04 | 5.13E-02 | 0.04 | 8.09E-02 | 0.04 | 9.81E-02 | 0.04 | 5.18E-02 | -0.03 | 1.03E-01 |
| rs10131280 | 14q32.33 | 107121607 | IGHgenecluster | G | 0.03 | 2.01E-01 | 0.04 | 1.13E-01 | 0.04 | 9.46E-02 | 0.04 | **4.92E-02** | -0.05 | **4.24E-02** |
| rs8025980 | 15q21.2 | 50994011 | SPPL2A | A | -0.02 | 3.65E-01 | -0.01 | 4.49E-01 | -0.01 | 3.94E-01 | -0.02 | 2.47E-01 | 0.02 | 3.24E-01 |
| rs602602 | 15q21.3 | 59057023 | MINDY2 | T | -0.02 | 3.43E-01 | -0.03 | 1.60E-01 | -0.02 | 2.25E-01 | -0.01 | 4.08E-01 | 0.00 | 9.12E-01 |
| rs117618017 | 15q22.2 | 63569902 | APH1B | T | -0.02 | 4.89E-01 | -0.02 | 4.45E-01 | -0.02 | 5.02E-01 | -0.02 | 3.66E-01 | 0.02 | 3.74E-01 |
| rs3848143 | 15q22.31 | 64423506 | SNX1 | G | -0.03 | 1.18E-01 | -0.02 | 2.89E-01 | -0.02 | 3.01E-01 | -0.03 | 1.05E-01 | 0.02 | 2.67E-01 |
| rs12592898 | 15q25.1 | 79229199 | CTSH | G | -0.02 | 4.47E-01 | -0.01 | 7.89E-01 | 0.01 | 7.68E-01 | -0.02 | 3.37E-01 | -0.01 | 5.15E-01 |
| rs1140239 | 16p11.2 | 30021402 | DOC2A | C | 0.00 | 7.72E-01 | 0.00 | 8.53E-01 | 0.00 | 8.19E-01 | 0.01 | 6.54E-01 | 0.00 | 8.50E-01 |
| rs889555 | 16p11.2 | 31122571 | BCKDK | C | -0.02 | 1.71E-01 | -0.02 | 3.27E-01 | -0.02 | 3.60E-01 | -0.02 | 1.50E-01 | 0.02 | 3.15E-01 |
| rs4985556 | 16q22.1 | 70694000 | IL34 | A | 0.02 | 3.31E-01 | 0.03 | 2.29E-01 | 0.03 | 1.86E-01 | 0.04 | 1.06E-01 | -0.03 | 1.57E-01 |
| rs450674 | 16q23.2 | 79608408 | MAF | T | 0.00 | 7.66E-01 | -0.01 | 4.07E-01 | -0.02 | 2.94E-01 | 0.01 | 5.54E-01 | 0.00 | 9.20E-01 |
| rs12446759 | 16q23.3 | 81773003 | PLCG2 | A | 0.02 | 3.65E-01 | 0.00 | 7.76E-01 | 0.00 | 9.80E-01 | 0.02 | 2.89E-01 | 0.00 | 7.81E-01 |
| rs72824905 | 16q23.3 | 81942028 | PLCG2 | C | 0.06 | 4.51E-01 | 0.07 | 3.96E-01 | 0.09 | 2.70E-01 | 0.11 | 1.30E-01 | -0.10 | 1.90E-01 |
| rs16941239 | 16q24.1 | 86454210 | FOXF1 | A | -0.10 | 7.70E-02 | -0.10 | 6.43E-02 | -0.08 | 1.38E-01 | -0.09 | 6.45E-02 | 0.07 | 1.57E-01 |
| rs56407236 | 16q24.3 | 90170095 | PRDM7 | A | 0.02 | 5.39E-01 | 0.01 | 8.53E-01 | 0.00 | 9.72E-01 | 0.04 | 2.27E-01 | 0.02 | 5.87E-01 |
| rs7225151 | 17p13.2 | 5137047 | SCIMP | A | -0.01 | 6.76E-01 | -0.02 | 3.89E-01 | -0.02 | 4.06E-01 | -0.01 | 7.90E-01 | 0.00 | 8.43E-01 |
| rs2242595 | 17p11.2 | 18059454 | MYO15A | G | 0.00 | 8.63E-01 | 0.00 | 9.05E-01 | 0.00 | 9.15E-01 | 0.01 | 7.22E-01 | -0.01 | 5.44E-01 |
| rs5848 | 17q21.31 | 42430244 | GRN | T | 0.02 | 3.72E-01 | 0.02 | 3.83E-01 | 0.01 | 5.34E-01 | 0.01 | 5.02E-01 | -0.01 | 7.16E-01 |
| **rs199515** | **17q21.31** | **44856641** | **WNT3** | C | 0.10 | **3.10E-06** | 0.09 | **1.84E-05** | 0.08 | **1.05E-04** | 0.05 | **1.19E-02** | -0.07 | **6.83E-04** |
| rs2526377 | 17q22 | 56410041 | TSPOAP1 | A | 0.01 | 6.17E-01 | 0.01 | 4.62E-01 | 0.01 | 4.41E-01 | 0.00 | 8.52E-01 | -0.01 | 7.28E-01 |
| rs4277405 | 17q23.3 | 61548918 | ACE | T | 0.01 | 6.57E-01 | 0.01 | 5.22E-01 | 0.01 | 4.58E-01 | 0.02 | 3.00E-01 | 0.00 | 8.07E-01 |
| rs12151021 | 19p13.3 | 1050874 | ABCA7 | A | -0.01 | 6.18E-01 | 0.00 | 8.72E-01 | 0.00 | 8.24E-01 | -0.01 | 7.01E-01 | 0.01 | 5.10E-01 |
| rs9304690 | 19q13.33 | 50453317 | SIGLEC11 | T | 0.02 | 2.95E-01 | 0.02 | 3.10E-01 | 0.03 | 1.69E-01 | 0.03 | 6.54E-02 | -0.03 | 6.34E-02 |
| rs587709 | 19q13.42 | 54771451 | LILRB2 | C | 0.03 | 8.04E-02 | 0.03 | 1.03E-01 | 0.02 | 1.80E-01 | 0.02 | 3.41E-01 | -0.01 | 7.02E-01 |
| rs1358782 | 20p13 | 393978 | RBCK1 | G | 0.01 | 6.65E-01 | 0.02 | 4.12E-01 | 0.01 | 5.50E-01 | 0.01 | 5.83E-01 | 0.00 | 9.79E-01 |
| rs6014724 | 20q13.2 | 54998544 | CASS4 | A | 0.02 | 5.65E-01 | 0.02 | 4.01E-01 | 0.03 | 3.69E-01 | 0.00 | 8.54E-01 | -0.02 | 5.40E-01 |
| rs6742 | 20q13.33 | 62374441 | SLC2A4RG | C | -0.01 | 6.78E-01 | 0.00 | 8.46E-01 | -0.01 | 7.52E-01 | 0.00 | 9.93E-01 | 0.02 | 4.00E-01 |
| rs2154481 | 21q21.3 | 27473875 | APP | T | 0.00 | 8.65E-01 | 0.00 | 8.60E-01 | 0.00 | 8.11E-01 | 0.00 | 8.38E-01 | -0.01 | 5.40E-01 |
| rs2830489 | 21q21.3 | 28148191 | ADAMTS1 | C | 0.03 | 1.15E-01 | 0.03 | 1.21E-01 | 0.02 | 2.62E-01 | 0.02 | 2.51E-01 | -0.01 | 5.85E-01 |

| SNP | Locus | Position | Nearest genes | Risk allele | Axial kurtosis | | Mean kurtosis | | Radial kurtosis | | Fractional anisotropy | | Mean diffusivity | |
|---|---|---|---|---|---|---|---|---|---|---|---|---|---|---|
| | | | | | Beta | p | Beta | p | Beta | p | Beta | p | Beta | p |
| **Parkinson disease** | | | | | | | | | | | | | | |
| rs114138760 | 1q21.3 | 154898185 | PMVK | C | 0.05 | 5.26E-01 | 0.09 | 2.77E-01 | 0.08 | 3.38E-01 | 0.07 | 3.84E-01 | -0.05 | 5.03E-01 |
| rs35749011 | 1q22 | 155135036 | KRTCAP2 | A | 0.14 | **4.81E-02** | 0.12 | 8.83E-02 | 0.11 | 1.30E-01 | 0.13 | **4.58E-02** | -0.13 | 5.89E-02 |
| rs6658353 | 1q23.3 | 161469054 | FCGR2A | C | 0.00 | 8.21E-01 | 0.00 | 9.17E-01 | -0.01 | 7.39E-01 | -0.01 | 6.94E-01 | 0.00 | 7.64E-01 |
| rs11578699 | 1q24.3 | 171719769 | VAMP4 | C | 0.01 | 6.74E-01 | 0.01 | 7.98E-01 | 0.00 | 8.37E-01 | 0.02 | 3.19E-01 | 0.00 | 8.60E-01 |
| rs823118 | 1q32.1 | 205723572 | NUCKS1 | T | 0.01 | 7.36E-01 | 0.01 | 6.11E-01 | 0.01 | 7.39E-01 | 0.01 | 7.33E-01 | -0.01 | 6.60E-01 |
| rs11557080 | 1q32.1 | 205737739 | RAB29 | A | -0.03 | 1.94E-01 | -0.04 | 8.09E-02 | -0.04 | 6.14E-02 | -0.03 | 1.34E-01 | 0.05 | **2.02E-02** |
| rs4653767 | 1q42.12 | 226916078 | ITPKB | T | 0.02 | 2.32E-01 | 0.02 | 2.36E-01 | 0.02 | 2.42E-01 | 0.02 | 2.21E-01 | -0.02 | 2.61E-01 |
| rs10797576 | 1q42.2 | 232664611 | SIPA1L2 | T | 0.04 | 1.15E-01 | 0.03 | 2.98E-01 | 0.02 | 4.36E-01 | 0.04 | 1.09E-01 | -0.02 | 5.01E-01 |
| rs76116224 | 2p24.2 | 18147848 | KCNS3 | A | 0.01 | 8.43E-01 | 0.00 | 9.07E-01 | -0.01 | 7.20E-01 | -0.01 | 7.32E-01 | 0.03 | 3.10E-01 |
| rs2042477 | 2q11.1 | 96000943 | KCNIP3 | T | -0.02 | 2.13E-01 | -0.03 | 1.58E-01 | -0.03 | 1.75E-01 | -0.02 | 1.81E-01 | 0.02 | 1.78E-01 |
| rs11683001 | 2q11.2 | 102396963 | MAP4K4 | A | -0.01 | 5.25E-01 | -0.02 | 3.93E-01 | -0.01 | 4.92E-01 | 0.00 | 7.84E-01 | 0.00 | 9.46E-01 |
| rs57891859 | 2q21.3 | 135464616 | TMEM163 | A | -0.04 | **3.11E-02** | -0.03 | 9.25E-02 | -0.02 | 2.46E-01 | -0.03 | **4.77E-02** | 0.02 | 1.42E-01 |
| rs1474055 | 2q24.3 | 169110394 | STK39 | T | 0.03 | 2.20E-01 | 0.05 | 7.22E-02 | 0.05 | 6.09E-02 | 0.01 | 5.21E-01 | -0.02 | 3.49E-01 |
| rs73038319 | 3p24.3 | 18361759 | SATB1 | C | 0.03 | 4.65E-01 | 0.02 | 5.30E-01 | 0.03 | 4.05E-01 | 0.03 | 3.58E-01 | -0.04 | 2.81E-01 |
| rs6808178 | 3p24.1 | 28705690 | LINC00693 | T | 0.02 | 3.10E-01 | 0.02 | 3.65E-01 | 0.01 | 5.49E-01 | 0.01 | 4.04E-01 | 0.00 | 7.61E-01 |
| rs12497850 | 3p21.31 | 48748989 | IP6K2 | T | 0.02 | 1.94E-01 | 0.03 | 6.85E-02 | 0.03 | 7.51E-02 | 0.03 | 5.12E-02 | -0.02 | 1.14E-01 |
| rs55961674 | 3q21.1 | 122196892 | KPNA1 | T | 0.03 | 2.06E-01 | 0.04 | 9.83E-02 | 0.04 | 5.86E-02 | 0.01 | 4.87E-01 | -0.03 | 1.61E-01 |
| rs11707416 | 3q25.1 | 151108965 | MED12L | T | 0.01 | 7.44E-01 | 0.00 | 9.38E-01 | -0.01 | 6.96E-01 | 0.00 | 9.56E-01 | 0.01 | 4.50E-01 |
| rs1450522 | 3q26.1 | 161077630 | SPTSSB | G | 0.04 | **3.57E-02** | 0.03 | 5.61E-02 | 0.02 | 1.84E-01 | 0.02 | 2.34E-01 | -0.02 | 3.03E-01 |
| rs10513789 | 3q27.1 | 182760073 | MCCC1 | T | -0.01 | 5.70E-01 | 0.00 | 9.17E-01 | 0.01 | 8.00E-01 | 0.00 | 8.24E-01 | 0.01 | 7.13E-01 |
| rs873786 | 4p16.3 | 925376 | GAK | C | 0.05 | 5.06E-02 | 0.04 | 1.26E-01 | 0.03 | 2.74E-01 | 0.04 | 8.40E-02 | -0.01 | 6.36E-01 |
| rs34311866 | 4p16.3 | 951947 | TMEM175 | C | 0.02 | 3.18E-01 | 0.01 | 4.87E-01 | 0.01 | 7.43E-01 | 0.01 | 5.09E-01 | 0.00 | 9.01E-01 |
| rs4698412 | 4p15.32 | 15737348 | BST1 | A | 0.01 | 6.68E-01 | 0.00 | 8.08E-01 | 0.00 | 8.42E-01 | 0.01 | 7.28E-01 | -0.01 | 7.02E-01 |
| rs34025766 | 4p15.31 | 17968811 | LCORL | T | 0.00 | 9.26E-01 | -0.01 | 5.57E-01 | -0.02 | 4.63E-01 | -0.02 | 4.53E-01 | 0.01 | 7.55E-01 |
| rs6825004 | 4q21.1 | 77110365 | SCARB2 | C | -0.01 | 7.37E-01 | -0.01 | 4.16E-01 | -0.02 | 3.05E-01 | -0.01 | 6.01E-01 | 0.00 | 9.18E-01 |
| rs4101061 | 4q21.1 | 77147969 | FAM47E | G | 0.00 | 9.80E-01 | 0.01 | 5.58E-01 | 0.02 | 3.98E-01 | -0.01 | 7.24E-01 | -0.02 | 1.45E-01 |
| rs6854006 | 4q21.1 | 77198054 | FAM47E-STBD1 | C | 0.01 | 6.44E-01 | 0.01 | 5.86E-01 | 0.01 | 4.91E-01 | 0.01 | 5.39E-01 | -0.01 | 6.11E-01 |
| rs356182 | 4q22.1 | 90626111 | SNCA | G | 0.00 | 9.33E-01 | 0.00 | 9.57E-01 | 0.00 | 9.04E-01 | 0.00 | 7.99E-01 | -0.01 | 5.20E-01 |
| rs5019538 | 4q22.1 | 90636630 | SNCA | G | -0.02 | 3.22E-01 | -0.01 | 5.37E-01 | -0.01 | 6.53E-01 | -0.02 | 2.54E-01 | 0.00 | 9.21E-01 |
| rs13117519 | 4q26 | 114369065 | CAMK2D | T | 0.01 | 7.31E-01 | 0.01 | 6.68E-01 | 0.01 | 7.96E-01 | 0.01 | 7.86E-01 | 0.01 | 4.72E-01 |
| rs62333164 | 4q33 | 170583157 | CLCN3 | G | -0.02 | 2.71E-01 | -0.01 | 6.04E-01 | 0.00 | 7.84E-01 | -0.01 | 5.46E-01 | 0.00 | 9.62E-01 |
| rs1867598 | 5q12.1 | 60137959 | ELOVL7 | G | 0.01 | 7.54E-01 | 0.00 | 8.66E-01 | 0.00 | 9.99E-01 | 0.01 | 8.12E-01 | 0.00 | 9.27E-01 |
| rs26431 | 5q21.1 | 102365794 | PAM | C | -0.01 | 5.66E-01 | 0.00 | 8.28E-01 | 0.02 | 3.73E-01 | -0.01 | 6.41E-01 | -0.02 | 3.52E-01 |
| rs11950533 | 5q31.1 | 134199105 | C5orf24 | C | -0.03 | 2.12E-01 | -0.03 | 2.61E-01 | -0.03 | 2.67E-01 | -0.03 | 2.50E-01 | 0.02 | 4.05E-01 |
| rs4140646 | 6p22.1 | 27738801 | LOC100131289 | A | 0.05 | **1.13E-02** | 0.05 | **1.15E-02** | 0.05 | **1.71E-02** | 0.05 | **7.26E-03** | -0.03 | 8.14E-02 |
| rs9261484 | 6p22.1 | 30108683 | TRIM40 | C | -0.01 | 4.44E-01 | -0.01 | 7.70E-01 | 0.00 | 9.11E-01 | 0.00 | 9.72E-01 | -0.03 | 7.35E-02 |
| rs112485576 | 6p21.32 | 32578772 | HLA-DRB5 | C | -0.08 | **6.04E-04** | -0.09 | **2.68E-04** | -0.07 | **1.93E-03** | -0.05 | **1.51E-02** | 0.05 | **1.46E-02** |

| SNP | Locus | Position | Nearest genes | Risk allele | Axial kurtosis Beta | p | Mean kurtosis Beta | p | Radial kurtosis Beta | p | Fractional anisotropy Beta | p | Mean diffusivity Beta | p |
|---|---|---|---|---|---|---|---|---|---|---|---|---|---|---|
| rs12528068 | 6q13 | 72487762 | RIMS1 | T | -0.02 | 3.71E-01 | -0.02 | 3.44E-01 | -0.02 | 3.94E-01 | -0.03 | 1.22E-01 | 0.01 | 4.37E-01 |
| rs997368 | 6q21 | 112243291 | FYN | A | -0.04 | 5.74E-02 | -0.04 | 5.46E-02 | -0.04 | **4.88E-02** | -0.04 | **3.68E-02** | 0.01 | 4.58E-01 |
| rs75859381 | 6q23.2 | 133210361 | RPS12 | C | -0.05 | 2.98E-01 | -0.03 | 4.69E-01 | -0.01 | 7.75E-01 | -0.02 | 5.83E-01 | 0.02 | 6.07E-01 |
| rs199351 | 7p15.3 | 23300049 | GPNMB | A | -0.02 | 1.83E-01 | -0.01 | 3.83E-01 | -0.01 | 5.35E-01 | -0.02 | 1.29E-01 | 0.01 | 5.33E-01 |
| rs76949143 | 7q11.21 | 66009851 | GS1-124K5.11 | T | -0.02 | 6.17E-01 | -0.02 | 6.68E-01 | -0.02 | 5.91E-01 | -0.04 | 3.40E-01 | 0.02 | 5.31E-01 |
| rs1293298 | 8p23.1 | 11712443 | CTSB | A | 0.03 | 1.04E-01 | 0.03 | 1.39E-01 | 0.02 | 2.25E-01 | 0.01 | 5.93E-01 | -0.01 | 6.10E-01 |
| rs620513 | 8p22 | 16697593 | FGF20 | G | -0.02 | 3.13E-01 | -0.02 | 3.84E-01 | -0.02 | 2.39E-01 | -0.01 | 5.28E-01 | 0.01 | 4.37E-01 |
| rs2280104 | 8p21.3 | 22525980 | BIN3 | T | -0.01 | 4.77E-01 | -0.01 | 4.44E-01 | -0.01 | 7.01E-01 | -0.01 | 4.14E-01 | 0.00 | 8.06E-01 |
| rs2086641 | 8q24.21 | 130901909 | FAM49B | C | 0.01 | 5.59E-01 | 0.01 | 4.84E-01 | 0.02 | 3.84E-01 | 0.01 | 5.44E-01 | -0.02 | 2.99E-01 |
| rs13294100 | 9p22.2 | 17579690 | SH3GL2 | G | -0.01 | 5.60E-01 | -0.01 | 4.82E-01 | -0.01 | 5.74E-01 | -0.01 | 3.43E-01 | 0.01 | 4.96E-01 |
| rs10756907 | 9p22.2 | 17727065 | SH3GL2 | G | 0.00 | 9.21E-01 | 0.00 | 8.66E-01 | 0.00 | 7.97E-01 | -0.02 | 1.95E-01 | 0.00 | 8.08E-01 |
| rs6476434 | 9p13.3 | 34046391 | UBAP2 | C | -0.03 | 1.27E-01 | -0.03 | 1.20E-01 | -0.02 | 2.18E-01 | -0.01 | 4.45E-01 | 0.00 | 7.73E-01 |
| rs896435 | 10p13 | 15557406 | ITGA8 | T | -0.04 | **2.72E-02** | -0.04 | **3.57E-02** | -0.04 | **4.13E-02** | -0.02 | 2.15E-01 | 0.02 | 1.31E-01 |
| rs10748818 | 10q24.32 | 104015279 | GBF1 | G | 0.00 | 8.83E-01 | 0.01 | 5.29E-01 | 0.02 | 4.05E-01 | 0.00 | 9.37E-01 | -0.01 | 5.36E-01 |
| rs72840788 | 10q26.11 | 121415685 | BAG3 | A | -0.03 | 1.66E-01 | -0.02 | 3.25E-01 | -0.02 | 3.60E-01 | -0.03 | 1.37E-01 | 0.02 | 2.18E-01 |
| rs117896735 | 10q26.11 | 121536327 | INPP5F | A | 0.02 | 7.97E-01 | 0.02 | 7.45E-01 | 0.04 | 5.82E-01 | 0.05 | 4.05E-01 | -0.05 | 3.89E-01 |
| rs7938782 | 11p15.4 | 10558777 | RNF141 | A | 0.00 | 8.42E-01 | -0.01 | 7.88E-01 | -0.01 | 7.82E-01 | 0.00 | 8.82E-01 | 0.01 | 7.92E-01 |
| rs12283611 | 11q14.1 | 83487277 | DLG2 | C | 0.00 | 9.15E-01 | -0.02 | 2.72E-01 | -0.02 | 1.76E-01 | 0.00 | 8.17E-01 | 0.01 | 3.65E-01 |
| rs3802920 | 11q25 | 133787001 | IGSF9B | T | 0.01 | 7.34E-01 | 0.00 | 9.05E-01 | -0.01 | 7.21E-01 | 0.01 | 5.72E-01 | -0.01 | 7.70E-01 |
| rs76904798 | 12q12 | 40614434 | LRRK2 | T | -0.02 | 3.87E-01 | -0.03 | 1.81E-01 | -0.03 | 2.20E-01 | -0.02 | 2.43E-01 | 0.03 | 1.39E-01 |
| rs7134559 | 12q13.11 | 46419086 | SCAF11 | C | 0.00 | 8.16E-01 | 0.00 | 8.81E-01 | 0.00 | 9.00E-01 | 0.00 | 9.91E-01 | 0.00 | 8.70E-01 |
| rs10847864 | 12q24.31 | 123326598 | HIP1R | T | 0.01 | 3.98E-01 | 0.02 | 3.67E-01 | 0.01 | 4.97E-01 | 0.02 | 1.14E-01 | -0.02 | 1.84E-01 |
| rs11610045 | 12q24.33 | 133063768 | FBRSL1 | A | 0.00 | 9.95E-01 | -0.01 | 5.56E-01 | -0.01 | 3.93E-01 | 0.00 | 7.70E-01 | 0.01 | 4.09E-01 |
| rs9568188 | 13q14.2 | 49927732 | CAB39L | T | 0.02 | 2.94E-01 | 0.02 | 3.88E-01 | 0.01 | 5.59E-01 | 0.01 | 5.33E-01 | -0.02 | 3.14E-01 |
| rs4771268 | 13q32.1 | 97865021 | MBNL2 | T | -0.06 | **3.16E-03** | -0.06 | **2.38E-03** | -0.05 | **4.79E-03** | -0.06 | **1.20E-03** | 0.05 | **3.60E-03** |
| rs12147950 | 14q21.1 | 37989270 | MIPOL1 | C | -0.01 | 7.16E-01 | 0.00 | 8.18E-01 | 0.00 | 8.85E-01 | 0.00 | 9.14E-01 | 0.00 | 8.29E-01 |
| rs11158026 | 14q22.2 | 55348869 | GCH1 | C | 0.00 | 8.31E-01 | 0.00 | 8.18E-01 | -0.01 | 5.28E-01 | 0.00 | 9.83E-01 | 0.01 | 6.60E-01 |
| rs3742785 | 14q24.3 | 75373034 | RPS6KL1 | A | -0.03 | 1.73E-01 | -0.02 | 2.69E-01 | -0.01 | 4.77E-01 | -0.02 | 1.97E-01 | 0.02 | 3.76E-01 |
| rs979812 | 14q31.3 | 88464264 | GALC | T | -0.01 | 5.68E-01 | 0.00 | 7.86E-01 | 0.00 | 8.59E-01 | 0.01 | 6.79E-01 | 0.00 | 7.58E-01 |
| rs2251086 | 15q22.2 | 61997385 | VPS13C | C | 0.02 | 3.92E-01 | 0.01 | 7.11E-01 | 0.00 | 9.38E-01 | 0.02 | 4.60E-01 | 0.00 | 9.53E-01 |
| rs6497339 | 16p12.3 | 19277493 | SYT17 | A | -0.01 | 3.65E-01 | -0.01 | 4.13E-01 | -0.01 | 6.09E-01 | 0.00 | 7.82E-01 | -0.01 | 7.09E-01 |
| rs2904880 | 16p11.2 | 28944396 | CD19 | G | -0.01 | 5.45E-01 | -0.01 | 7.13E-01 | 0.00 | 7.79E-01 | -0.01 | 3.78E-01 | 0.00 | 8.63E-01 |
| rs11150601 | 16p11.2 | 30977799 | SETD1A | A | -0.02 | 2.99E-01 | -0.01 | 5.54E-01 | -0.01 | 7.60E-01 | -0.02 | 1.22E-01 | 0.01 | 4.74E-01 |
| rs6500328 | 16q12.1 | 50736656 | NOD2 | A | 0.02 | 2.80E-01 | 0.01 | 4.18E-01 | 0.01 | 5.93E-01 | 0.01 | 3.58E-01 | -0.01 | 3.40E-01 |
| rs3104783 | 16q12.2 | 52636242 | CASC16 | A | -0.01 | 6.44E-01 | -0.01 | 4.64E-01 | -0.01 | 3.71E-01 | -0.01 | 5.17E-01 | 0.03 | 8.25E-02 |
| rs10221156 | 16q12.2 | 52969426 | CHD9 | G | -0.03 | 3.11E-01 | -0.03 | 2.42E-01 | -0.03 | 3.19E-01 | -0.02 | 5.66E-01 | 0.04 | 1.22E-01 |
| rs12600861 | 17p13.1 | 7355621 | CHRNB1 | C | 0.03 | **4.31E-02** | 0.03 | 5.04E-02 | 0.03 | 1.09E-01 | 0.03 | **2.61E-02** | -0.03 | 8.78E-02 |
| rs12951632 | 17q21.2 | 40741013 | RETREG3 | T | 0.00 | 8.14E-01 | 0.00 | 9.10E-01 | -0.01 | 5.74E-01 | -0.01 | 4.44E-01 | 0.03 | 8.30E-02 |

| | | | | | Axial kurtosis | | Mean kurtosis | | Radial kurtosis | | Fractional anisotropy | | Mean diffusivity | |
|---|---|---|---|---|---|---|---|---|---|---|---|---|---|---|
| SNP | Locus | Position | Nearest genes | Risk allele | Beta | p | Beta | p | Beta | p | Beta | p | Beta | p |
| rs2269906 | 17q21.31 | 42294337 | UBTF | A | -0.03 | 1.04E-01 | -0.03 | 1.35E-01 | -0.02 | 1.50E-01 | -0.02 | 2.75E-01 | 0.02 | 1.79E-01 |
| rs850738 | 17q21.31 | 42434630 | FAM171A2 | G | 0.01 | 3.78E-01 | 0.02 | 3.17E-01 | 0.01 | 5.00E-01 | 0.01 | 5.00E-01 | 0.00 | 7.78E-01 |
| **rs62053943** | **17q21.31** | **43744203** | **CRHR1** | C | 0.11 | **7.55E-06** | 0.10 | **3.97E-05** | 0.09 | **1.48E-04** | 0.05 | **1.35E-02** | -0.08 | **7.05E-04** |
| rs117615688 | 17q21.31 | 43798308 | CRHR1 | G | 0.08 | **3.62E-02** | 0.07 | 1.07E-01 | 0.05 | 2.12E-01 | 0.03 | 3.77E-01 | -0.06 | 1.03E-01 |
| rs11658976 | 17q21.31 | 44866805 | WNT3 | G | 0.04 | **8.62E-03** | 0.04 | **3.35E-02** | 0.03 | 5.28E-02 | 0.03 | 8.39E-02 | -0.03 | 5.87E-02 |
| rs61169879 | 17q23.2 | 59917366 | BRIP1 | T | 0.00 | 9.71E-01 | 0.01 | 7.98E-01 | 0.01 | 7.55E-01 | 0.02 | 3.92E-01 | -0.02 | 2.86E-01 |
| rs666463 | 17q25.3 | 76425480 | DNAH17 | A | 0.01 | 8.06E-01 | 0.00 | 9.46E-01 | -0.01 | 7.96E-01 | 0.02 | 4.39E-01 | 0.01 | 6.91E-01 |
| rs1941685 | 18q12.1 | 31304318 | ASXL3 | T | -0.02 | 1.31E-01 | -0.01 | 3.71E-01 | -0.01 | 7.04E-01 | -0.02 | 1.78E-01 | 0.00 | 8.53E-01 |
| rs12456492 | 18q12.3 | 40673380 | RIT2 | G | 0.02 | 1.79E-01 | 0.03 | 8.25E-02 | 0.03 | 9.67E-02 | 0.02 | 1.67E-01 | -0.02 | 3.21E-01 |
| rs8087969 | 18q21.2 | 48683589 | MEX3C | G | -0.01 | 5.40E-01 | -0.01 | 4.50E-01 | -0.02 | 3.00E-01 | -0.01 | 3.91E-01 | 0.01 | 3.75E-01 |
| rs55818311 | 19p13.3 | 2341047 | SPPL2B | C | 0.03 | 5.93E-02 | 0.04 | 5.12E-02 | 0.03 | 7.71E-02 | 0.02 | 3.03E-01 | -0.02 | 2.02E-01 |
| rs77351827 | 20p12.3 | 6006041 | CRLS1 | T | -0.02 | 4.04E-01 | -0.01 | 6.04E-01 | -0.02 | 4.98E-01 | -0.03 | 1.81E-01 | 0.01 | 5.45E-01 |
| rs2248244 | 21q22.13 | 38852361 | DYRK1A | A | -0.01 | 6.18E-01 | -0.01 | 5.13E-01 | -0.01 | 4.30E-01 | -0.02 | 3.03E-01 | 0.02 | 1.89E-01 |
| **PVS-Basal ganglia** | | | | | | | | | | | | | | |
| rs4675310 | 2q33.2 | 203880834 | NBEAL1,ICA1L | A | 0.04 | 8.54E-02 | 0.04 | 1.53E-01 | 0.03 | 3.03E-01 | 0.02 | 4.60E-01 | -0.01 | 6.66E-01 |
| rs6769442 | 3q26.31 | 171565463 | TMEM212 | G | -0.03 | 1.57E-01 | -0.03 | 1.56E-01 | -0.02 | 2.32E-01 | -0.03 | 6.16E-02 | 0.03 | 1.34E-01 |
| **PVS-Hippocampus** | | | | | | | | | | | | | | |
| rs10797812 | 1q25.3 | 182984597 | SHCBP1L,LAMC1 | A | 0.02 | 1.40E-01 | 0.02 | 2.93E-01 | 0.01 | 6.71E-01 | 0.02 | 9.46E-02 | 0.00 | 8.17E-01 |
| rs6540873 | 1q41 | 215137222 | CENPF,KCNK2 | A | -0.01 | 6.97E-01 | -0.01 | 4.05E-01 | -0.02 | 3.23E-01 | -0.01 | 3.90E-01 | 0.04 | **1.81E-02** |
| rs78857879 | 2p16.1 | 56135099 | EFEMP1 | G | -0.03 | 4.08E-01 | -0.03 | 4.06E-01 | -0.03 | 3.17E-01 | -0.01 | 6.56E-01 | 0.04 | 1.85E-01 |
| **PVS-White matter** | | | | | | | | | | | | | | |
| rs10494988 | 1q41 | 215141570 | CENPF,KCNK2 | C | -0.01 | 7.59E-01 | -0.01 | 4.61E-01 | -0.02 | 3.70E-01 | -0.01 | 4.61E-01 | 0.04 | **2.12E-02** |
| rs7596872 | 2p16.1 | 56128091 | EFEMP1 | C | -0.01 | 7.00E-01 | -0.01 | 6.77E-01 | -0.02 | 4.99E-01 | 0.00 | 9.94E-01 | 0.03 | 3.11E-01 |
| **rs13079464** | **3p25.1** | **13822439** | **WNT7A** | C | -0.06 | **7.94E-05** | -0.07 | **4.08E-05** | -0.06 | **1.11E-04** | -0.06 | **1.17E-04** | 0.06 | **2.72E-04** |
| **rs4685022** | **3p25.1** | **13832611** | **WNT7A** | G | -0.10 | **1.21E-08** | -0.11 | **1.23E-10** | -0.10 | **1.48E-09** | -0.07 | **1.20E-05** | 0.09 | **4.15E-09** |
| rs3772833 | 3q21.2 | 124518362 | ITGB5,UMPS | G | -0.03 | 1.58E-01 | -0.03 | 1.32E-01 | -0.04 | 1.02E-01 | -0.03 | 1.10E-01 | 0.03 | 1.10E-01 |
| rs1922930 | 6p25.3 | 1364691 | FOXQ1,FOXF2 | C | 0.02 | 4.74E-01 | 0.02 | 5.28E-01 | 0.01 | 6.13E-01 | -0.01 | 7.73E-01 | 0.00 | 9.34E-01 |
| rs4959689 | 6p25.2 | 2617122 | C6orf195 | C | -0.02 | 2.93E-01 | -0.02 | 2.11E-01 | -0.02 | 1.43E-01 | -0.02 | 1.18E-01 | 0.03 | **2.67E-02** |
| rs10954468 | 7q33 | 134434661 | BPGM,CALD1 | C | -0.04 | **1.57E-02** | -0.04 | **2.81E-02** | -0.03 | 1.16E-01 | -0.04 | **4.84E-03** | 0.03 | 9.01E-02 |
| rs2923437 | 8p11.21 | 42425399 | SMIM19,CHRNB3,SLC20A2 | A | -0.01 | 4.70E-01 | -0.02 | 2.34E-01 | -0.02 | 2.05E-01 | -0.02 | 3.09E-01 | 0.02 | 2.11E-01 |
| **rs10817108** | **9q31.3** | **113658671** | **LPAR1** | A | -0.05 | **1.66E-02** | -0.05 | **1.32E-02** | -0.05 | **1.07E-02** | -0.05 | **2.51E-03** | 0.07 | **1.40E-04** |
| rs12417836 | 11q13.3 | 70089700 | FADD,PPFIA1 | T | 0.02 | 5.31E-01 | 0.03 | 3.00E-01 | 0.03 | 2.86E-01 | 0.01 | 6.51E-01 | -0.04 | 1.59E-01 |
| rs8041189 | 15q25.3 | 85686327 | PDE8A | G | 0.02 | 3.67E-01 | 0.01 | 6.07E-01 | 0.00 | 9.50E-01 | 0.01 | 4.49E-01 | 0.00 | 8.48E-01 |
| rs1126642 | 17q21.31 | 42989063 | GFAP | C | 0.02 | 6.96E-01 | 0.03 | 4.34E-01 | 0.05 | 2.67E-01 | 0.00 | 9.29E-01 | -0.03 | 4.75E-01 |
| rs2425881 | 20q13.12 | 45255618 | SLC13A3 | A | -0.01 | 7.20E-01 | -0.01 | 7.61E-01 | 0.00 | 8.51E-01 | 0.00 | 9.16E-01 | 0.02 | 4.03E-01 |
| rs2425884 | 20q13.12 | 45258292 | SLC13A3 | C | -0.01 | 4.92E-01 | -0.01 | 4.20E-01 | -0.01 | 5.61E-01 | -0.01 | 5.50E-01 | 0.01 | 4.23E-01 |
| rs6011998 | 20q13.12 | 45269867 | SLC13A3 | C | -0.04 | 2.59E-01 | -0.05 | 1.82E-01 | -0.07 | 7.24E-02 | -0.04 | 2.83E-01 | 0.08 | **1.82E-02** |
| rs112407396 | 20q13.12 | 45276381 | SLC13A3 | T | -0.01 | 7.86E-01 | 0.02 | 6.32E-01 | 0.04 | 3.96E-01 | -0.01 | 7.64E-01 | -0.02 | 5.93E-01 |

| SNP | Locus | Position | Nearest genes | Risk allele | Axial kurtosis | | Mean kurtosis | | Radial kurtosis | | Fractional anisotropy | | Mean diffusivity | |
|---|---|---|---|---|---|---|---|---|---|---|---|---|---|---|
| | | | | | Beta | p | Beta | p | Beta | p | Beta | p | Beta | p |
| rs56104388 | 20q13.12 | 45302135 | SLC13A3 | T | -0.01 | 8.73E-01 | -0.02 | 7.08E-01 | -0.03 | 6.19E-01 | -0.02 | 7.59E-01 | -0.01 | 8.44E-01 |
| rs72485816 | 20q13.12 | 45314435 | TP53RK,SLC13A3 | T | -0.07 | 1.46E-01 | -0.08 | 1.00E-01 | -0.08 | 9.77E-02 | -0.06 | 1.87E-01 | 0.08 | 6.98E-02 |
| **Stroke** | | | | | | | | | | | | | | |
| rs2455132 | 1p36.32 | 3221083 | PRDM16 | C | -0.02 | 3.52E-01 | -0.03 | 1.82E-01 | -0.03 | 1.37E-01 | -0.02 | 1.61E-01 | 0.03 | 1.45E-01 |
| rs880315 | 1p36.22 | 10796866 | CASZ1 | C | 0.00 | 8.73E-01 | 0.00 | 8.51E-01 | 0.00 | 9.61E-01 | 0.00 | 8.19E-01 | 0.00 | 8.36E-01 |
| rs3790607 | 1p13.2 | 113053023 | WNT2B | C | 0.02 | 5.24E-01 | 0.02 | 4.88E-01 | 0.02 | 5.62E-01 | 0.03 | 3.33E-01 | -0.04 | 2.08E-01 |
| rs2251636 | 1q22 | 156202809 | PMF1 | G | -0.01 | 6.65E-01 | -0.01 | 6.89E-01 | 0.00 | 7.90E-01 | 0.00 | 9.00E-01 | 0.00 | 9.59E-01 |
| rs680084 | 1q24.2 | 170628255 | PRRX1 | G | 0.01 | 6.95E-01 | 0.00 | 8.65E-01 | -0.01 | 6.86E-01 | 0.00 | 9.70E-01 | 0.01 | 7.41E-01 |
| rs2877984 | 1q25.3 | 183090497 | LAMC1 | G | 0.02 | 2.72E-01 | 0.01 | 3.82E-01 | 0.01 | 6.82E-01 | 0.02 | 1.94E-01 | 0.00 | 8.49E-01 |
| rs11694327 | 2p23.3 | 26919429 | KCNK3 | C | 0.01 | 5.56E-01 | 0.02 | 3.24E-01 | 0.02 | 3.45E-01 | 0.00 | 9.66E-01 | -0.02 | 3.65E-01 |
| rs6722806 | 2p21 | 43627715 | THADA | A | 0.02 | 3.56E-01 | 0.01 | 5.04E-01 | 0.00 | 9.36E-01 | 0.01 | 4.55E-01 | 0.00 | 9.40E-01 |
| rs11691032 | 2q24.3 | 164788513 | FIGN | C | 0.01 | 6.69E-01 | 0.00 | 9.77E-01 | -0.01 | 6.64E-01 | 0.01 | 5.83E-01 | 0.00 | 9.31E-01 |
| rs2351524 | 2q33.2 | 203880992 | NBEAL1 | C | 0.04 | 7.69E-02 | 0.04 | 1.41E-01 | 0.03 | 2.88E-01 | 0.02 | 4.29E-01 | -0.01 | 6.34E-01 |
| rs4681330 | 3q24 | 146378341 | PLSCR5 | C | -0.02 | 2.84E-01 | -0.02 | 2.25E-01 | -0.02 | 2.52E-01 | -0.01 | 3.63E-01 | 0.03 | 7.96E-02 |
| rs16998073 | 4q21.21 | 81184341 | FGF5 | T | 0.01 | 5.88E-01 | 0.00 | 8.04E-01 | 0.00 | 8.73E-01 | 0.01 | 6.20E-01 | 0.00 | 9.56E-01 |
| rs6847935 | 4q25 | 111696651 | PITX2 | T | -0.02 | 4.17E-01 | -0.02 | 3.97E-01 | -0.02 | 3.13E-01 | -0.01 | 4.58E-01 | 0.01 | 6.80E-01 |
| rs6536024 | 4q31.3 | 155543369 | FGG | C | 0.04 | **1.73E-02** | 0.03 | **3.54E-02** | 0.03 | 6.10E-02 | 0.04 | **4.04E-03** | -0.03 | 8.23E-02 |
| rs4444878 | 4q35.2 | 187213883 | F11 | A | -0.03 | 1.08E-01 | -0.02 | 1.46E-01 | -0.02 | 1.90E-01 | -0.02 | 1.26E-01 | 0.02 | 2.73E-01 |
| rs17148926 | 5q23.2 | 121510586 | LOC100505841 | A | -0.01 | 6.83E-01 | -0.01 | 7.93E-01 | -0.01 | 7.71E-01 | 0.00 | 9.55E-01 | 0.02 | 4.10E-01 |
| rs79318212 | 6p25.3 | 1365244 | FOXF2 | G | 0.01 | 7.41E-01 | 0.01 | 8.11E-01 | 0.00 | 9.30E-01 | -0.02 | 3.95E-01 | 0.01 | 5.91E-01 |
| rs36229526 | 6p21.32 | 32820656 | TAP1 | T | 0.04 | 1.70E-01 | 0.05 | 9.75E-02 | 0.05 | 1.09E-01 | 0.03 | 3.39E-01 | -0.03 | 2.83E-01 |
| rs1574430 | 6p21.1 | 43269029 | SLC22A7 | A | -0.02 | 2.09E-01 | -0.02 | 3.02E-01 | -0.01 | 4.33E-01 | -0.01 | 3.60E-01 | 0.02 | 2.95E-01 |
| rs2501966 | 6p12.3 | 49457835 | CENPQ | A | -0.04 | **2.50E-02** | -0.03 | **4.76E-02** | -0.03 | 1.09E-01 | -0.03 | **4.12E-02** | 0.03 | 8.44E-02 |
| rs56393506 | 6q26 | 161089307 | LPA | T | 0.00 | 9.74E-01 | 0.00 | 8.57E-01 | 0.01 | 7.22E-01 | 0.00 | 8.48E-01 | 0.00 | 9.97E-01 |
| rs2107595 | 7p21.1 | 19049388 | HDAC9 | A | -0.01 | 6.47E-01 | -0.01 | 7.37E-01 | -0.01 | 7.20E-01 | -0.01 | 4.93E-01 | 0.01 | 5.24E-01 |
| rs7800053 | 7p15.2 | 27316640 | EVX1 | G | -0.01 | 6.46E-01 | -0.02 | 4.94E-01 | -0.02 | 3.94E-01 | 0.00 | 8.46E-01 | 0.00 | 8.47E-01 |
| rs113916171 | 7p12.1 | 51190564 | COBL | A | -0.04 | 2.23E-01 | -0.03 | 3.73E-01 | -0.03 | 3.77E-01 | -0.05 | 7.19E-02 | 0.02 | 5.03E-01 |
| rs42035 | 7q21.2 | 92239531 | CDK6 | A | -0.01 | 6.22E-01 | -0.01 | 7.04E-01 | -0.01 | 6.81E-01 | 0.01 | 5.41E-01 | 0.00 | 9.35E-01 |
| rs12539561 | 7q22.3 | 106437611 | PIK3CG | C | 0.01 | 6.63E-01 | 0.01 | 7.29E-01 | 0.01 | 7.92E-01 | 0.01 | 7.89E-01 | -0.01 | 7.19E-01 |
| rs114798023 | 7q31.1 | 108198301 | THAP5 | G | 0.01 | 8.60E-01 | -0.05 | 4.79E-01 | -0.10 | 1.57E-01 | -0.04 | 5.53E-01 | 0.09 | 1.54E-01 |
| rs1549758 | 7q36.1 | 150695726 | NOS3 | T | 0.02 | 2.77E-01 | 0.02 | 3.20E-01 | 0.02 | 3.13E-01 | 0.01 | 3.56E-01 | -0.03 | 1.13E-01 |
| rs2738158 | 8p23.1 | 6749669 | DEFB1 | G | 0.01 | 7.86E-01 | 0.00 | 9.88E-01 | 0.00 | 9.72E-01 | 0.04 | 1.36E-01 | -0.03 | 1.87E-01 |
| rs4298492 | 8p21.2 | 26024889 | EBF2 | A | 0.00 | 9.19E-01 | 0.00 | 8.32E-01 | 0.01 | 7.66E-01 | 0.03 | 8.87E-02 | -0.01 | 4.62E-01 |
| rs1487504 | 9p22.2 | 17000855 | BNC2 | A | 0.02 | 5.23E-01 | 0.02 | 5.62E-01 | 0.02 | 3.96E-01 | 0.00 | 8.58E-01 | -0.01 | 7.78E-01 |
| rs7859362 | 9p21.3 | 22105927 | CDKN2B-AS1 | C | 0.04 | **1.14E-02** | 0.04 | **1.64E-02** | 0.03 | **4.23E-02** | 0.04 | **1.71E-02** | -0.03 | 6.91E-02 |
| rs2405068 | 9q22.32 | 98386311 | PTCH1 | A | 0.02 | 2.29E-01 | 0.02 | 2.81E-01 | 0.02 | 3.37E-01 | 0.03 | 1.39E-01 | -0.03 | 1.79E-01 |
| rs649129 | 9q34.2 | 136154304 | ABO | T | 0.01 | 5.75E-01 | 0.02 | 2.79E-01 | 0.02 | 2.34E-01 | -0.01 | 7.66E-01 | -0.01 | 4.70E-01 |
| rs55983834 | 10q24.33 | 105601210 | SH3PXD2A | C | 0.01 | 6.55E-01 | 0.00 | 8.35E-01 | 0.00 | 9.00E-01 | 0.01 | 7.31E-01 | 0.01 | 5.63E-01 |

| | | | | | Axial kurtosis | | Mean kurtosis | | Radial kurtosis | | Fractional anisotropy | | Mean diffusivity | |
|---|---|---|---|---|---|---|---|---|---|---|---|---|---|---|
| SNP | Locus | Position | Nearest genes | Risk allele | Beta | p | Beta | p | Beta | p | Beta | p | Beta | p |
| rs10886430 | 10q26.11 | 121010256 | GRK5 | G | -0.05 | **3.74E-02** | -0.05 | 6.09E-02 | -0.04 | 9.47E-02 | -0.03 | 2.11E-01 | 0.04 | 9.12E-02 |
| rs60401382 | 10q26.13 | 124227624 | HTRA1 | C | 0.02 | 3.52E-01 | 0.02 | 2.42E-01 | 0.03 | 1.87E-01 | 0.02 | 2.74E-01 | -0.03 | 1.03E-01 |
| rs1973765 | 11p15.5 | 1898664 | LSP1 | C | 0.00 | 7.67E-01 | 0.00 | 8.77E-01 | 0.00 | 9.00E-01 | 0.01 | 3.86E-01 | 0.00 | 9.87E-01 |
| rs415895 | 11p15.4 | 9769562 | SWAP70 | G | -0.02 | 2.02E-01 | -0.03 | 1.22E-01 | -0.03 | 1.32E-01 | -0.02 | 1.68E-01 | 0.01 | 4.52E-01 |
| rs72985562 | 11q22.2 | 102800278 | MMP12 | G | 0.03 | 3.78E-01 | 0.04 | 1.63E-01 | 0.05 | 9.54E-02 | 0.04 | 1.80E-01 | -0.05 | 1.09E-01 |
| rs7304841 | 12p12.2 | 20577593 | PDE3A | A | -0.01 | 6.27E-01 | -0.01 | 3.85E-01 | -0.02 | 2.82E-01 | 0.00 | 9.45E-01 | 0.01 | 4.92E-01 |
| rs7973143 | 12q13.13 | 52263869 | ANKRD33 | T | -0.01 | 5.05E-01 | -0.01 | 5.94E-01 | -0.01 | 6.03E-01 | -0.02 | 3.33E-01 | 0.01 | 3.87E-01 |
| rs12426667 | 12q13.13 | 54433483 | HOXC4 | A | -0.02 | 1.75E-01 | -0.03 | 1.28E-01 | -0.03 | 1.48E-01 | -0.03 | 1.17E-01 | 0.01 | 4.03E-01 |
| rs12579302 | 12q21.33 | 90050503 | ATP2B1 | A | -0.02 | 2.64E-01 | -0.03 | 1.36E-01 | -0.03 | 2.15E-01 | -0.01 | 7.42E-01 | 0.01 | 7.94E-01 |
| rs10774625 | 12q24.12 | 111910219 | SH2B3 | A | 0.03 | 1.07E-01 | 0.03 | 1.12E-01 | 0.03 | 8.94E-02 | 0.02 | 1.01E-01 | -0.03 | 6.96E-02 |
| rs7974266 | 12q24.13 | 113007602 | PTPN11 | T | 0.02 | 3.18E-01 | 0.02 | 2.60E-01 | 0.02 | 2.84E-01 | 0.01 | 4.13E-01 | -0.03 | 6.57E-02 |
| rs35429 | 12q24.21 | 115555867 | 12q24 | A | 0.00 | 8.59E-01 | -0.01 | 6.99E-01 | -0.01 | 4.95E-01 | -0.01 | 5.67E-01 | 0.01 | 3.77E-01 |
| rs842365 | 13q14.13 | 47252379 | LRCH1 | A | 0.04 | **1.60E-02** | 0.04 | **2.82E-02** | 0.04 | **4.69E-02** | 0.04 | **8.86E-03** | -0.04 | **1.36E-02** |
| rs9515201 | 13q34 | 111040798 | COL4A2 | A | 0.00 | 8.04E-01 | 0.00 | 8.39E-01 | 0.00 | 9.39E-01 | -0.01 | 6.99E-01 | 0.01 | 4.55E-01 |
| rs2663905 | 15q25.1 | 81359137 | MESDC1 | G | 0.00 | 9.60E-01 | -0.01 | 7.31E-01 | -0.01 | 5.58E-01 | 0.00 | 8.40E-01 | -0.01 | 6.39E-01 |
| rs1573644 | 15q26.1 | 91421283 | FURIN | C | 0.01 | 5.41E-01 | 0.00 | 7.88E-01 | 0.00 | 8.30E-01 | 0.02 | 3.11E-01 | -0.01 | 6.19E-01 |
| rs2397816 | 15q26.2 | 96119880 | LINC00924 | A | 0.04 | **3.61E-02** | 0.03 | 1.01E-01 | 0.02 | 3.69E-01 | 0.03 | 7.10E-02 | -0.01 | 3.67E-01 |
| rs2359171 | 16q22.3 | 73053022 | ZFHX3 | A | -0.03 | 1.26E-01 | -0.02 | 2.64E-01 | -0.02 | 3.45E-01 | -0.03 | 1.60E-01 | 0.02 | 4.09E-01 |
| rs12445022 | 16q24.2 | 87575332 | ZCCHC14 | A | 0.00 | 8.08E-01 | 0.01 | 7.51E-01 | 0.01 | 4.69E-01 | 0.00 | 8.60E-01 | -0.02 | 2.31E-01 |
| rs2316757 | 17q21.32 | 45066225 | RPRML | A | -0.01 | 7.43E-01 | -0.01 | 6.53E-01 | -0.01 | 7.09E-01 | -0.01 | 7.52E-01 | 0.02 | 3.39E-01 |
| rs28860769 | 19p13.2 | 10737581 | SLC44A2 | G | 0.02 | 4.39E-01 | 0.01 | 4.78E-01 | 0.02 | 3.51E-01 | 0.01 | 5.63E-01 | -0.01 | 4.53E-01 |
| rs8106503 | 19p13.2 | 11196886 | LDLR | T | -0.01 | 7.08E-01 | -0.01 | 5.92E-01 | -0.01 | 6.76E-01 | -0.02 | 4.92E-01 | 0.00 | 8.81E-01 |
| rs11907011 | 20q11.22 | 33767770 | PROCR | C | -0.04 | 1.07E-01 | -0.03 | 1.92E-01 | -0.03 | 2.85E-01 | -0.03 | 1.59E-01 | 0.03 | 2.76E-01 |
| **WMH** | | | | | | | | | | | | | | |
| rs786921 | 1p22.2 | 89286673 | PKN2 | A | 0.01 | 5.73E-01 | 0.01 | 4.11E-01 | 0.01 | 4.67E-01 | 0.01 | 4.93E-01 | -0.02 | 3.04E-01 |
| rs73923006 | 2p21 | 43132224 | HAAO | G | -0.01 | 7.84E-01 | -0.01 | 7.68E-01 | -0.01 | 6.38E-01 | 0.01 | 6.49E-01 | 0.00 | 8.16E-01 |
| rs7596872 | 2p16.1 | 56128091 | EFEMP1 | A | 0.01 | 7.00E-01 | 0.01 | 6.77E-01 | 0.02 | 4.99E-01 | 0.00 | 9.94E-01 | -0.03 | 3.11E-01 |
| rs62172472 | 2q32.1 | 188028317 | CALCRL | G | 0.00 | 8.69E-01 | 0.01 | 6.74E-01 | 0.01 | 5.10E-01 | 0.02 | 3.47E-01 | -0.02 | 3.86E-01 |
| rs7603972 | 2q33.2 | 203780515 | CARF1 | A | 0.04 | 7.09E-02 | 0.04 | 1.18E-01 | 0.03 | 2.43E-01 | 0.02 | 3.88E-01 | -0.01 | 5.97E-01 |
| rs6797002 | 3q27.1 | 183363263 | KLHL24 | C | -0.02 | 3.68E-01 | -0.02 | 2.32E-01 | -0.02 | 1.93E-01 | -0.02 | 2.42E-01 | 0.03 | 8.93E-02 |
| **rs17205972** | **5q14.3** | **82859065** | **VCAN** | T | -0.20 | **7.74E-23** | -0.20 | **5.87E-22** | -0.18 | **7.08E-19** | -0.19 | **8.65E-23** | 0.17 | **1.07E-19** |
| rs2303655 | 5q23.2 | 121518378 | LOC100505841 | T | -0.02 | 3.47E-01 | -0.02 | 4.57E-01 | -0.02 | 4.70E-01 | -0.01 | 5.76E-01 | 0.03 | 1.61E-01 |
| rs6940540 | 6q25.1 | 151018909 | PLEKHG1 | G | 0.02 | 1.77E-01 | 0.03 | **4.62E-02** | 0.03 | **3.82E-02** | 0.00 | 8.82E-01 | -0.02 | 1.50E-01 |
| rs73184312 | 8p23.1 | 8179639 | SGK223 | G | 0.01 | 4.92E-01 | 0.00 | 8.84E-01 | 0.00 | 8.61E-01 | 0.00 | 7.92E-01 | 0.00 | 7.73E-01 |
| rs11249945 | 8p23.1 | 9628753 | TNKS | A | 0.02 | 3.60E-01 | 0.01 | 5.56E-01 | 0.01 | 7.40E-01 | 0.01 | 4.93E-01 | -0.01 | 6.36E-01 |
| rs7004825 | 8p23.1 | 11031472 | XKR6 | T | 0.01 | 7.30E-01 | 0.01 | 7.37E-01 | 0.00 | 8.49E-01 | 0.00 | 8.03E-01 | -0.01 | 5.96E-01 |
| rs4630220 | 10q24.33 | 105459116 | SH3PXD2A | G | -0.01 | 7.49E-01 | 0.00 | 8.74E-01 | 0.00 | 8.50E-01 | -0.02 | 3.34E-01 | 0.00 | 8.24E-01 |
| rs71471298 | 10q24.33 | 105507145 | SH3PXD2A-AS1 | T | 0.01 | 7.14E-01 | -0.01 | 6.09E-01 | -0.02 | 3.30E-01 | 0.01 | 8.09E-01 | 0.01 | 8.12E-01 |

| SNP | Locus | Position | Nearest genes | Risk allele | Axial kurtosis | | Mean kurtosis | | Radial kurtosis | | Fractional anisotropy | | Mean diffusivity | |
|---|---|---|---|---|---|---|---|---|---|---|---|---|---|---|
| | | | | | Beta | p | Beta | p | Beta | p | Beta | p | Beta | p |
| rs10786772 | 10q24.33 | 105610326 | SH3PXD2A | G | 0.01 | 7.55E-01 | 0.00 | 9.50E-01 | 0.00 | 7.95E-01 | 0.00 | 8.82E-01 | 0.01 | 4.58E-01 |
| rs55940034 | 13q34 | 111043309 | COL4A2 | G | 0.00 | 7.89E-01 | -0.01 | 7.75E-01 | 0.00 | 8.75E-01 | -0.01 | 6.98E-01 | 0.02 | 3.38E-01 |
| rs72680374 | 14q22.1 | 52604843 | NID2 | T | 0.02 | 3.19E-01 | 0.02 | 3.80E-01 | 0.02 | 3.55E-01 | 0.01 | 4.78E-01 | -0.01 | 4.07E-01 |
| rs1285847 | 14q32.11 | 91884655 | CCDC88C | T | 0.01 | 4.26E-01 | 0.01 | 4.35E-01 | 0.01 | 5.36E-01 | 0.01 | 4.42E-01 | -0.01 | 7.04E-01 |
| rs7157599 | 14q32.2 | 100625902 | DEGS2 | C | 0.01 | 6.09E-01 | 0.01 | 7.44E-01 | 0.00 | 8.41E-01 | 0.00 | 8.55E-01 | 0.00 | 9.64E-01 |
| rs12443113 | 15q22.31 | 65355468 | RASL12 | G | -0.02 | 2.85E-01 | -0.02 | 2.51E-01 | -0.02 | 2.55E-01 | -0.03 | 8.21E-02 | 0.02 | 2.34E-01 |
| rs1948948 | 16q12.1 | 51442679 | SALL1 | C | -0.02 | 1.83E-01 | -0.02 | 3.40E-01 | 0.00 | 8.69E-01 | 0.01 | 6.96E-01 | -0.03 | **4.52E-02** |
| rs12921170 | 16q24.2 | 87227397 | C16orf95 | A | 0.01 | 4.43E-01 | 0.01 | 5.28E-01 | 0.01 | 3.87E-01 | 0.02 | 1.89E-01 | -0.03 | 7.26E-02 |
| rs6503417 | 17q21.31 | 43144218 | NMT1 | C | -0.03 | 1.25E-01 | -0.03 | 5.66E-02 | -0.04 | **3.53E-02** | -0.01 | 3.94E-01 | 0.01 | 3.46E-01 |
| rs34974290 | 17q25.1 | 73888354 | TRIM65 | A | -0.05 | **1.53E-02** | -0.06 | **1.81E-03** | -0.07 | **8.49E-04** | -0.05 | **1.51E-02** | 0.05 | **1.40E-02** |
| rs5762197 | 22q12.1 | 27887471 | MN1 | C | -0.02 | 2.08E-01 | -0.03 | 1.32E-01 | -0.03 | 1.37E-01 | -0.02 | 2.48E-01 | 0.01 | 4.29E-01 |

In bold and highlighted: $p<2.16.10^{-4}$. In bold only: $p<0.05$

Table S6: Associations of DKI loci with baseline and longitudinal cognitive scores in the Rhineland Study

| Trait | Time | SNP | Locus | Position | Gene | EA | NEA | Beta | SE | p |
|---|---|---|---|---|---|---|---|---|---|---|
| Executive function | T0-T1 | rs3852188 | 5q14.3 | 82858014 | VCAN-AS1 | C | G | -0.01 | 0.00 | 0.0159 |
| Executive function | T0-T1 | rs76928645 | 7p11.2 | 54941328 | SEC61G-DT;EGFR | C | T | 0.01 | 0.00 | 0.0053 |
| Episodic verbal memory | T0-T1 | rs76928645 | 7p11.2 | 54941328 | SEC61G-DT;EGFR | C | T | 0.01 | 0.00 | 0.0145 |
| Working memory | T0 | rs76928645 | 7p11.2 | 54941328 | SEC61G-DT;EGFR | C | T | -0.06 | 0.02 | 0.0205 |
| Global cognition | T0-T1 | rs76928645 | 7p11.2 | 54941328 | SEC61G-DT;EGFR | C | T | 0.01 | 0.00 | 0.0307 |
| Processing speed | T0 | rs76928645 | 7p11.2 | 54941328 | SEC61G-DT;EGFR | C | T | 0.04 | 0.02 | 0.0358 |
| Executive function | T0 | rs13225457 | 7p22.3 | 541830 | PDGFA | A | G | 0.03 | 0.01 | 0.0410 |
| Processing speed | T0 | rs35585955 | 9q31.1 | 104996411 | GRIN3A;LINC00587 | T | C | 0.03 | 0.01 | 0.0343 |
| Processing speed | T0 | rs140531353 | 22q13.31 | 45088305 | PRR5 | A | C | 0.09 | 0.05 | 0.0479 |

Only results with $p<0.05$ are shown.
EA: Effect allele; NEA: Non-effect allele; Beta: Effect size of effect allele (increase of DKI); SE: Standard error.
T0: Baseline cognition; T0-T1: Cognitive trajectory between baseline and follow-up.

Table S7: Results of transcriptome-wide association study of DKI markers

| Tissue | Gene | CHR | Start | Stop | NWGT | MODEL | TWAS.Z | TWAS.P | JOINT.Z | JOINT.P | COLOC.PP4 |
|---|---|---|---|---|---|---|---|---|---|---|---|
| **Axial kurtosis** | | | | | | | | | | | |
| Brain Caudate basal ganglia | SYPL2 | 1 | 110009180 | 110024759 | 12 | enet | -4.58 | 4.63E-06 | -4.60 | 4.60E-06 | 0.32 |
| Artery Tibial | VCAN | 5 | 82767284 | 82878122 | 19 | enet | -9.41 | 4.83E-21 | -9.40 | 4.80E-21 | 0.53 |
| Artery Tibial | C6orf106 | 6 | 34555065 | 34664636 | 8 | lasso | 4.48 | 7.32E-06 | 4.50 | 7.30E-06 | **0.83** |
| Adipose Subcutaneous | ARHGAP27 | 17 | 43471275 | 43511787 | 183 | susie | 4.95 | 7.55E-07 | 4.90 | 7.50E-07 | **0.92** |
| Brain Caudate basal ganglia | LRRC37A4P | 17 | 43578685 | 43627701 | 135 | susie | 4.83 | 1.37E-06 | 4.80 | 1.40E-06 | **0.99** |
| Adipose Visceral Omentum | RP11-798G7.8 | 17 | 43608943 | 43611204 | 20 | enet | -5.08 | 3.70E-07 | -5.10 | 3.70E-07 | **0.99** |
| Artery Tibial | RP11-798G7.8 | 17 | 43608943 | 43611204 | 10 | lasso | -4.85 | 1.22E-06 | -4.90 | 1.20E-06 | **0.99** |
| Nerve Tibial | RP11-798G7.8 | 17 | 43608943 | 43611204 | 12 | lasso | -4.99 | 6.19E-07 | -5.00 | 6.20E-07 | **0.99** |
| Brain Spinal cord cervical c-1 | RP11-707O23.5 | 17 | 43678235 | 43679706 | 13 | lasso | -4.96 | 7.17E-07 | -5.00 | 7.20E-07 | **0.99** |
| Heart Atrial Appendage | CRHR1-IT1 | 17 | 43697694 | 43725582 | 14 | lasso | -4.82 | 1.47E-06 | -4.80 | 1.50E-06 | **0.99** |
| Brain Cortex | SPPL2C | 17 | 43922256 | 43924438 | 31 | susie | -4.84 | 1.30E-06 | -4.80 | 1.30E-06 | 0.61 |
| Brain Amygdala | KANSL1-AS1 | 17 | 44270942 | 44274089 | 13 | lasso | -4.95 | 7.56E-07 | -4.90 | 7.60E-07 | **0.99** |
| Brain Cerebellum | KANSL1-AS1 | 17 | 44270942 | 44274089 | 14 | lasso | -5.01 | 5.45E-07 | -5.00 | 5.40E-07 | **0.99** |
| Brain Nucleus accumbens basal ganglia | KANSL1-AS1 | 17 | 44270942 | 44274089 | 16 | enet | -4.83 | 1.34E-06 | -4.80 | 1.30E-06 | **0.99** |
| Brain Substantia nigra | KANSL1-AS1 | 17 | 44270942 | 44274089 | 12 | lasso | -4.94 | 7.90E-07 | -4.90 | 7.90E-07 | **0.99** |
| Brain Anterior cingulate cortex BA24 | RP11-259G18.2 | 17 | 44320972 | 44322410 | 21 | enet | -4.85 | 1.26E-06 | -4.80 | 1.30E-06 | **0.97** |
| Brain Hypothalamus | RP11-259G18.2 | 17 | 44320972 | 44322410 | 24 | enet | -4.91 | 9.13E-07 | -4.90 | 9.10E-07 | **0.97** |
| Whole Blood | RP11-259G18.2 | 17 | 44320972 | 44322410 | 28 | enet | -5.10 | 3.44E-07 | -5.10 | 3.40E-07 | **0.96** |
| Artery Aorta | RP11-259G18.3 | 17 | 44336917 | 44337972 | 24 | enet | -4.91 | 9.05E-07 | -4.90 | 9.10E-07 | **0.97** |
| Heart Left Ventricle | RP11-259G18.3 | 17 | 44336917 | 44337972 | 16 | lasso | -4.82 | 1.42E-06 | -4.80 | 1.40E-06 | **0.98** |
| Artery Coronary | RP11-259G18.1 | 17 | 44344403 | 44346060 | 65 | susie | -4.92 | 8.58E-07 | -4.90 | 8.60E-07 | **0.94** |
| Brain Frontal Cortex BA9 | RP11-259G18.1 | 17 | 44344403 | 44346060 | 66 | susie | -4.85 | 1.23E-06 | -4.90 | 1.20E-06 | **0.97** |
| Brain Putamen basal ganglia | RP11-259G18.1 | 17 | 44344403 | 44346060 | 64 | susie | -4.86 | 1.18E-06 | -4.90 | 1.20E-06 | **0.97** |
| Brain Hippocampus | ARL17A | 17 | 44594068 | 44657088 | 8 | lasso | -5.00 | 5.64E-07 | -5.00 | 5.60E-07 | **0.97** |
| Brain Cerebellar Hemisphere | FAM215B | 17 | 44636196 | 44640161 | 6 | lasso | -4.91 | 9.03E-07 | -4.90 | 9.00E-07 | **0.98** |
| **Mean kurtosis** | | | | | | | | | | | |
| Brain Putamen basal ganglia | AC093495.4 | 3 | 14186223 | 14189784 | 32 | enet | -4.93 | 8.38E-07 | -4.90 | 8.40E-07 | 0.13 |
| Artery Tibial | VCAN | 5 | 82767284 | 82878122 | 19 | enet | -9.06 | 1.27E-19 | -9.10 | 1.30E-19 | 0.53 |
| Brain Cortex | SNX31 | 8 | 101585116 | 101675643 | 28 | enet | -4.62 | 3.81E-06 | -4.60 | 3.80E-06 | **0.91** |
| Artery Tibial | C17orf104 | 17 | 42733762 | 42767676 | 1 | top1 | -4.52 | 6.24E-06 | -4.50 | 6.20E-06 | **0.96** |
| Brain Cerebellar Hemisphere | PLEKHM1 | 17 | 43513266 | 43568115 | 30 | enet | 4.55 | 5.43E-06 | 4.50 | 5.40E-06 | **0.97** |
| Nerve Tibial | RP11-798G7.8 | 17 | 43608943 | 43611204 | 12 | lasso | -4.52 | 6.30E-06 | -4.50 | 6.30E-06 | **0.97** |

| Tissue | Gene | CHR | Start | Stop | NWGT | MODEL | TWAS.Z | TWAS.P | JOINT.Z | JOINT.P | COLOC.PP4 |
|---|---|---|---|---|---|---|---|---|---|---|---|
| Brain Anterior cingulate cortex BA24 | CRHR1-IT1 | 17 | 43697694 | 43725582 | 89 | susie | -4.49 | 7.21E-06 | -4.50 | 7.20E-06 | **0.97** |
| Brain Cerebellum | KANSL1-AS1 | 17 | 44270942 | 44274089 | 14 | lasso | -4.45 | 8.59E-06 | -4.50 | 8.60E-06 | **0.97** |
| Whole Blood | RP11-259G18.2 | 17 | 44320972 | 44322410 | 28 | enet | -4.59 | 4.43E-06 | -4.60 | 4.40E-06 | **0.96** |
| Adipose Visceral Omentum | RP11-259G18.3 | 17 | 44336917 | 44337972 | 29 | enet | -4.48 | 7.33E-06 | -4.50 | 7.30E-06 | **0.95** |
| Brain Amygdala | RP11-259G18.3 | 17 | 44336917 | 44337972 | 27 | enet | -4.45 | 8.49E-06 | -4.50 | 8.50E-06 | **0.96** |
| Brain Hypothalamus | RP11-259G18.1 | 17 | 44344403 | 44346060 | 66 | susie | -4.46 | 8.09E-06 | -4.50 | 8.10E-06 | **0.96** |
| Brain Putamen basal ganglia | RP11-259G18.1 | 17 | 44344403 | 44346060 | 64 | susie | -4.46 | 8.39E-06 | -4.50 | 8.40E-06 | **0.96** |
| Brain Hippocampus | ARL17A | 17 | 44594068 | 44657088 | 8 | lasso | -4.63 | 3.66E-06 | -4.60 | 3.70E-06 | **0.96** |
| Artery Tibial | WNT3 | 17 | 44839872 | 44910520 | 255 | susie | -4.59 | 4.51E-06 | -4.60 | 4.50E-06 | **0.97** |
| Whole Blood | GALK1 | 17 | 73747675 | 73761792 | 6 | lasso | 4.47 | 7.87E-06 | 4.50 | 7.90E-06 | **0.96** |
| **Radial kurtosis** | | | | | | | | | | | |
| Brain Putamen basal ganglia | AC093495.4 | 3 | 14186223 | 14189784 | 32 | enet | -4.59 | 4.34E-06 | -4.60 | 4.30E-06 | 0.14 |
| Artery Tibial | VCAN | 5 | 82767284 | 82878122 | 19 | enet | -8.47 | 2.37E-17 | -8.50 | 2.40E-17 | 0.51 |
| Brain Cortex | SNX31 | 8 | 101585116 | 101675643 | 28 | enet | -4.58 | 4.59E-06 | -4.60 | 4.60E-06 | **0.90** |
| Artery Tibial | C17orf104 | 17 | 42733762 | 42767676 | 1 | top1 | -4.50 | 6.70E-06 | -4.50 | 6.70E-06 | **0.96** |
| Whole Blood | GALK1 | 17 | 73747675 | 73761792 | 6 | lasso | 4.47 | 7.97E-06 | 4.50 | 8.00E-06 | **0.96** |

Only significant genes after correction for multiple testing ($8.9x10^{-6}$) and conditionally significant ($p<0.05$) are shown. In bold: COLOC.PP4≥0.75.
TWAS.Z: Z-score from TWAS analyses; TWAS.P: pvalue from TWAS analyses; JOINT.Z: Z-score from conditional analyses; Joint.P: pvalue from conditional analyses; COLOC.PP4: Posterior probability 4 from colocalization analyses, corresponding to the probability to colocalized functional/GWAS associations; MODEL: Best performing model; NWGT: Number of SNPs with non-zero weights in the model.
AK: Axial kurtosis; MK: Mean kurtosis; RK: Radial kurtosis.

Table S8: Association of proteins in DKI GWAS loci with DKI markers in the Rhineland Study

| Protein | OlinkID | UniProt | Locus | Beta | SE | p |
|---|---|---|---|---|---|---|
| **Axial kurtosis** | | | | | | |
| CD300A | OID30576 | Q9UGN4 | 17q25.1 | -0.19 | 0.05 | 2.60E-04 |
| CD300C | OID21010 | Q08708 | 17q25.1 | -0.20 | 0.04 | 1.25E-05 |
| CD300E | OID21418 | Q496F6 | 17q25.1 | -0.14 | 0.04 | 3.58E-04 |
| **Mean kurtosis** | | | | | | |
| VCAN | OID21026 | P13611 | 5q14.2 | -0.19 | 0.05 | 1.26E-04 |
| GFAP | OID21247 | P14136 | 17q21.31 | -0.14 | 0.03 | 1.83E-05 |
| CD300C | OID21010 | Q08708 | 17q25.1 | -0.19 | 0.05 | 4.37E-05 |
| CD300E | OID21418 | Q496F6 | 17q25.1 | -0.13 | 0.04 | 7.62E-04 |
| **Radial kurtosis** | | | | | | |
| VCAN | OID21026 | P13611 | 5q14.2 | -0.19 | 0.05 | 7.92E-05 |
| MAD1L1 | OID20904 | Q9Y6D9 | 7p22.3 | -0.16 | 0.05 | 4.46E-04 |
| EGFR | OID20319 | P00533 | 7p11.2 | 0.28 | 0.08 | 2.69E-04 |
| CES1 | OID20244 | P23141 | 16q12.2 | 0.08 | 0.02 | 2.87E-04 |
| GFAP | OID21247 | P14136 | 17q21.31 | -0.16 | 0.03 | 2.20E-06 |
| CD300C | OID21010 | Q08708 | 17q25.1 | -0.16 | 0.04 | 3.21E-04 |

Only significant proteins after correction for multiple testing ($8.06x10^{-4}$) are shown

Table S9: Association of proteins in DKI GWAS loci with cognition and WMH in the Rhineland Study

| Trait | Protein | OlinkID | UniProt | Locus | Beta | SE | p |
|---|---|---|---|---|---|---|---|
| **Imaging - Baseline** | | | | | | | |
| White matter hyperintensities | CD300A | OID30576 | Q9UGN4 | 17q25.1 | 0.08 | 0.04 | 4.05E-02 |
| White matter hyperintensities | CD300C | OID21010 | Q08708 | 17q25.1 | 0.07 | 0.03 | 3.86E-02 |
| White matter hyperintensities | CD300E | OID21418 | Q496F6 | 17q25.1 | 0.09 | 0.03 | 3.13E-03 |
| White matter hyperintensities | CES1 | OID20244 | P23141 | 16q12.2 | 0.04 | 0.02 | 1.23E-02 |
| White matter hyperintensities | MAD1L1 | OID20904 | Q9Y6D9 | 7p22.3 | 0.12 | 0.04 | **6.62E-04** |
| **Cognition - Baseline** | | | | | | | |
| Episodic verbal memory | CD300A | OID30576 | Q9UGN4 | 17q25.1 | -0.10 | 0.04 | 2.48E-02 |
| Executive function | CD300A | OID30576 | Q9UGN4 | 17q25.1 | -0.11 | 0.05 | 1.82E-02 |
| Global cognition | CD300A | OID30576 | Q9UGN4 | 17q25.1 | -0.08 | 0.04 | 2.63E-02 |
| Total memory | CD300A | OID30576 | Q9UGN4 | 17q25.1 | -0.09 | 0.04 | 3.27E-02 |
| Episodic verbal memory | CD300C | OID21010 | Q08708 | 17q25.1 | -0.10 | 0.04 | 1.16E-02 |
| Global cognition | GFAP | OID21247 | P14136 | 17q21.31 | 0.05 | 0.02 | 4.33E-02 |
| Working memory | GFAP | OID21247 | P14136 | 17q21.31 | 0.08 | 0.03 | 8.09E-03 |
| Executive function | MAD1L1 | OID20904 | Q9Y6D9 | 7p22.3 | -0.11 | 0.04 | 1.39E-02 |
| **Cognition - Longitudinal** | | | | | | | |
| Episodic verbal memory | CD300A | OID30576 | Q9UGN4 | 17q25.1 | -0.01 | 0.00 | 3.36E-03 |
| Global cognition | CD300A | OID30576 | Q9UGN4 | 17q25.1 | 0.00 | 0.00 | 3.45E-02 |
| Executive function | CD300E | OID21418 | Q496F6 | 17q25.1 | -0.01 | 0.00 | 2.35E-03 |
| Global cognition | CD300E | OID21418 | Q496F6 | 17q25.1 | -0.01 | 0.00 | 1.10E-02 |
| Episodic verbal memory | EGFR | OID20319 | P00533 | 7p11.2 | 0.01 | 0.00 | 4.52E-03 |
| Global cognition | EGFR | OID20319 | P00533 | 7p11.2 | 0.01 | 0.00 | **9.31E-05** |
| Processing speed | EGFR | OID20319 | P00533 | 7p11.2 | 0.01 | 0.00 | 9.51E-03 |
| Total memory | EGFR | OID20319 | P00533 | 7p11.2 | 0.01 | 0.00 | 1.73E-03 |
| Episodic verbal memory | GFAP | OID21247 | P14136 | 17q21.31 | -0.01 | 0.00 | **1.33E-04** |
| Executive function | GFAP | OID21247 | P14136 | 17q21.31 | -0.02 | 0.00 | **1.79E-13** |
| Global cognition | GFAP | OID21247 | P14136 | 17q21.31 | -0.02 | 0.00 | **2.85E-15** |
| Processing speed | GFAP | OID21247 | P14136 | 17q21.31 | -0.02 | 0.00 | **2.34E-07** |
| Total memory | GFAP | OID21247 | P14136 | 17q21.31 | -0.01 | 0.00 | **2.46E-05** |
| Working memory | GFAP | OID21247 | P14136 | 17q21.31 | -0.01 | 0.00 | 1.69E-02 |
| Episodic verbal memory | MAD1L1 | OID20904 | Q9Y6D9 | 7p22.3 | -0.01 | 0.00 | **2.89E-04** |
| Executive function | MAD1L1 | OID20904 | Q9Y6D9 | 7p22.3 | -0.01 | 0.00 | 2.49E-03 |
| Global cognition | MAD1L1 | OID20904 | Q9Y6D9 | 7p22.3 | -0.01 | 0.00 | **1.71E-06** |
| Processing speed | MAD1L1 | OID20904 | Q9Y6D9 | 7p22.3 | -0.01 | 0.00 | 4.10E-03 |
| Total memory | MAD1L1 | OID20904 | Q9Y6D9 | 7p22.3 | -0.01 | 0.00 | 1.17E-03 |
| Executive function | VCAN | OID21026 | P13611 | 5q14.2 | -0.01 | 0.00 | 4.59E-02 |

Only results with $p<0.05$ are shown. Significant results ($p<8.93x10^{-4}$) are in bold.
